# Vaccination strategies to achieve outbreak control for MPXV Clade I with a one-time mass campaign in sub-Saharan Africa: a scenario-based modelling study

**DOI:** 10.1101/2025.01.17.25320753

**Authors:** Shihui Jin, Toshiaki R Asakura, Hiroaki Murayama, David Niyukuri, Diafuka Saila-Ngita, Jue Tao Lim, Akira Endo, Borame L Dickens

## Abstract

**Background:** Limited mpox vaccination coverage, declining cross-protection from historical smallpox vaccination campaigns, and persistent zoonotic reservoirs leave many sub-Saharan countries susceptible to mpox outbreaks. With millions of vaccine doses made available to the region since late 2024 and the absence of country-specific guidelines for allocation, estimating the country-specific impact of one-time mass vaccination strategies is necessary for ongoing outbreaks and other countries at future risk.

**Methods and findings:** We adapted a next generation matrix model to project disease transmission potential for 47 sub-Saharan countries from 2025 to 2050 under four transmission scenarios with different contributions of community vs sexual contacts. The model was informed by mpox datasets from Clade Ia and Ib outbreaks in the Democratic Republic of Congo (DRC), and incorporated country-specific demographics, contact patterns, and historical smallpox vaccination coverage. We then assessed strategies to reduce disease transmissibility by calculating country-specific minimum vaccine coverages and evaluating one-time, age-specific mass vaccination campaigns. At least 20 of the 47 countries are estimated to require vaccination in 2025, and 36 in 2050, to guard against potentially forthcoming national outbreaks. For 11 Clade I-affected countries, the minimum required coverage is estimated to range from 0%–4.8% to 14.5%–19.5% in 2025 with increasing sexual transmission, rising to 0%–10.8% and 22.6%–26.0% in 2050, respectively. The prioritised age group for vaccination shifts from 0–4 years to 20–29 years with additional sexual transmission. Note that all the projections were based on the best available evidence of Clade I transmission in the DRC as of December 2024.

**Interpretation:** With diminishing smallpox-vaccination-induced immunity, increasing disease transmissibility and potential for persistent outbreaks are expected for mpox, necessitating growing vaccine demand for outbreak containment. Given probable supply constraints, our findings could guide the ongoing and future mass vaccination efforts in sub-Saharan Africa by emphasising the prioritisation of high-risk groups, where allocation strategies are tailored to the evolving epidemiological landscape of each country.

## 1. Introduction

The monkeypox virus (MPVX), causing mpox disease, has now been responsible for multiple international outbreaks with sustained human-to-human transmission across its two clades: Clade I and Clade II. Historically, mpox has been reported in rural, forested areas within central and western Africa where infection is suspected to primarily occur through zoonotic exposure, with limited human spread resulting from close physical contact [1]. In 2023 however, the epidemiology of Clade I shifted with the emergence of the subclade Ib, an offshoot of Clade I. For this subclade, evidence exists of more sustained and rapid spread through physical and sexual contact in the general population [2], with the greatest proportion of reported cases being among children under five and young adults aged 20–29 years [3], as well as high-risk populations such as sex workers and their clients [4]. Overall, since January 2024, over 50,000 cases and 1000 deaths have been reported in the Democratic Republic of Congo (DRC) and surrounding affected countries for both subclades combined [3].

With Clade I currently estimated to have higher case fatality ratios (up to 12% versus 3.6% for Clade II) [5], public health systems within sub-Saharan Africa are concerned about the potential impacts of Clade I importations and outbreaks. This is exacerbated by the lack of rapid vaccine distribution [6], current low mpox vaccination coverage in affected countries [7], and waning cross-protection from historical smallpox vaccination campaigns [8], which had coverage of more than 90% across sub-Saharan Africa at its peak [9] and ended around the time when the World Health Organisation (WHO) declared smallpox to be eradicated in 1980.

Overall, the persistence of mpox, increasing transmissibility between humans, and loss of cross-protection have supported immunisation strategies using third-generation smallpox vaccines that show improved safety profiles [10]. The effective coordination and planning for the distribution of these vaccines and stockpiling will be of priority, which should be guided by evidence and scenario-based demand assessments and projections.

As of January 2025, approximately 0.5 million vaccines have been delivered to six sub-Saharan countries, with four (Central African Republic, the DRC, Nigeria, and Rwanda) having launched mpox vaccination campaigns [11]. An additional 4.83 million vaccine doses are expected to be available this year through the Vaccines Access and Allocation Mechanism for mpox, including 1.73 million doses of the MVA-BN smallpox vaccine, 50,000 doses of the ACAM smallpox vaccine, and over three million doses of the LC16m8 smallpox vaccine [11]. While MVA-BN has demonstrated 70–90% effectiveness against mpox infection (predominantly Clade IIb) in fully vaccinated individuals (two doses), the other two single-dose vaccines, despite not being widely administered in real-world settings for mpox infection prevention, have been reported to show comparable effectiveness based on surrogate endpoints [12–15]. These efforts are critical but coverage will be limited as the total population size in these countries is more than 20 million as of 2024, rising to over 1.2 billion for sub-Saharan Africa where more countries could be potentially at risk [16]. However, limited research exists on how mpox vaccination campaigns should be carried out in sub-Saharan African countries currently experiencing or at risk of outbreaks [17], leaving local authorities with little guidance on how to allocate donated doses. With Clade I mpox outbreaks expected to continue occurring across this region due to the persistent zoonotic reservoir, the increase in transboundary movement of people and transmissibility of Clade I MPXV, the future demand of vaccines for a responsive mass vaccination campaign against mpox outbreaks needs to be estimated for stockpiling estimations and effective outbreak control.

Firstly considerations are required on how these vaccines should be distributed, taking into account both waning smallpox immunity and the contribution of sexual and non-sexual contact to overall transmission. Secondly, estimates of the country-specific differences in the number of doses required to avert outbreaks, according to differences in demographics and size of high-risk populations, are required to assist in short-term vaccine stockpiling. Thirdly, in the event that mass immunisation coverage is limited, determining which age groups require prioritisation can assist in minimising health impacts.

To explore these, we constructed a mathematical model of a mpox vaccination program, adapted from a preliminary study [18], to project the transmission potential of mpox in 47 sub-Saharan African countries at high risk of case importation and outbreak. This model was then utilised to estimate the minimum vaccine stockpiles required to prevent epidemic growth under four transmissibility scenarios of increasing levels in sexual transmission. We further explored the impacts of diverse vaccine allocation strategies across age groups when supplies were limited. Our findings aim to provide detailed and practical guidance for policy-makers to tailor efficient vaccination strategies, ensuring the optimal use of available resources and enhancing outbreak preparedness.

## 2. Methods

### 2.1 Overview

Our analyses focused on the impact of a one-time, proactive mpox vaccination campaign in 47 sub-Saharan African countries, assessing its effectiveness in reducing disease transmissibility during an outbreak occurring immediately afterward. We used country-specific data to model geospatial variations in transmission dynamics, including population projections by age and sex, age-dependent contact matrices, smallpox vaccination coverage, and estimates of the population sizes of the high-sexual-activity groups (Table S1–S2) [9,16,19,20]. Outcomes were evaluated at five-year intervals between 2025 and 2050.

Considering the large regional discrepancies in the contribution of sexual transmission to overall disease dynamics and the differences in transmission patterns between Clade Ia and Ib mpox outbreaks, as identified in our preliminary study [18], we modelled four hypothetical Clade I outbreak scenarios with varying levels of sexual transmission. The contribution of sexual transmission to overall transmission in each scenario was assumed to range between that observed in historical Clade Ia outbreaks in the endemic provinces (i.e., zero) and that inferred from the epidemiological data for the Clade Ib outbreak in North and South Kivu provinces, the DRC as of August 2024 [18], when the confirmed cases surged exponentially. These levels were informed by frequency of high-sexual-activity males contacting females, including:

1. Community contact only: with disease transmission exclusively explained by social contacts within the general population (which we assumed can be represented by synthetic home contact matrices, as household transmission was reported to account for 93% of the non-sexually acquired cases from Burundi between 25 July and 26 October 2024 [21]);
2. Low sexual transmission: social contact with additional sexual contact contributing 9.1% (95% confidence interval [CI]: 6.1%–13%) to overall transmission in the DRC in 2024;
3. Moderate sexual transmission: social contact with additional sexual contact contributing 31% (95% CI: 26%–35%) to overall transmission in the DRC in 2024; and
4. High sexual transmission: social contact with additional sexual contact contributing 52% (95% CI: 48%–55%) to overall transmission in the DRC in 2024.

The parameterisation of these four scenarios is also summarised in the Supplementary Information (Table S3).

### 2.2 Model of Clade I MPXV transmission dynamics

To model the effect of vaccination on the Clade I MPXV transmission dynamics in sub-Saharan African countries, we employed the next generation matrix model from the previous study [18] calibrated to historical and recent Clade I MPXV outbreaks. The next generation matrix characterises the heterogeneous transmission patterns across population groups stratified by different age, sex, and sexual activity level. It accounts for community contact routes relevant to both Clade Ia and Ib, as well as the sexual transmission pathway specific to Clade Ib. The effective reproduction number, *R_eff_*, could be derived as the largest eigenvalue of this matrix. The original model used empirical contact survey data from Zimbabwe and age-dependent susceptibility parameters to characterise transmission of Clade Ia MPXV through regular community contact between age groups. For Clade Ib, the population was further stratified by sex and sexual activity level, and frequent sexual contact among the high-sexual-activity group (a subset of individuals aged 15–49 years) was also assumed to contribute to transmission in addition to community contact. To ensure generalisability and consistency across countries beyond the scope of the original model (i.e., the DRC and Burundi), we introduced minor adaptations to the model: we replaced the empirical contact matrix with the synthetic contact matrices (home contact only, with all contact assessed in a sensitivity analysis), which are available for most of the African countries; the assumed size of high-sexual-activity groups in the model was rescaled for each country according to the reported population size estimates of sex workers and males engaging in commercial sex [20,22]; age-specific coverage of the historical smallpox vaccination was varied between countries using previous estimates by Taube et al [9]. The adapted model was then re-calibrated to mpox case data in the DRC. A total of 1000 sets of updated parameter estimates were generated through bootstrapping and used in the subsequent analysis for the 47 sub-Saharan African countries included in this study. These countries were selected from regions with documented community transmission of Clade I MPXV as of December 2024 [23], based on the availability of synthetic contact matrices, a key input to the model. Detailed descriptions of the model structure and parameterisation, the calibration process, and validation are provided in the Supplementary Information (Fig S1–S4, Table S1–S3).

### 2.3 Projection of the possible future effective reproduction numbers of Clade I MPXV in sub-Saharan African countries

Using the estimated smallpox vaccine effectiveness and country-specific smallpox vaccine coverage estimates by year of birth [9], we projected the time evolution of the next generation matrix in each country from 2025 to 2050, where we adjusted the susceptibility among different age groups (reflecting the smallpox-immunised proportion) as they age, assuming age-dependent contact patterns remain similar over time. Following Murayama et al. [18], we derived the time- and country-dependent *R_eff_* as the largest eigenvalue of the next generation matrix. The matrix was scaled to match an estimated *R_eff_*= 0.82 (with uncertainty introduced in a subsequent sensitivity analysis) for Clade Ia MPXV in 2015 (midpoint of the study period of 2013–2017) in the DRC [24], implicitly assuming the same secondary attack rate across Clade I subclades [25,26]. The projected *R_eff_*s for Clade I outbreaks in the DRC in 2024 by such scaling were consistent with the estimates reported by Marziano et al. [27].

### 2.4 Minimum vaccine coverage to prevent epidemic growth

To inform potential future vaccine rollout strategies, we determined the minimum mpox vaccine coverage required to prevent the epidemic growth of Clade I MPXV, which would only be achieved with the optimal allocation of vaccines across age groups (henceforth referred to as the ‘optimal vaccine allocation strategy’). We used an iterative approach, starting from an entirely unvaccinated population. We allocated each incremental set of *n* vaccine series, where *n* was less than 1% of a country’s total population size, to the group which would most efficiently reduce *R_eff_*. We set an upper limit of 80% [28] on the vaccine uptake for any age group (which we varied in the sensitivity analysis). The effectiveness of mpox vaccines was assumed to be 80% following MVA-BN two-dose regimen (which we also varied in the sensitivity analysis) [12]. The iteration was repeated until the herd immunity threshold *R_eff_*< 1 was achieved. Further details are described in the Supplementary Information (S1 Text).

We included all age groups as being vaccine-eligible in the main analysis but as additional analyses we also explored two other scenarios: (i) only individuals aged 20 years or older eligible for vaccination; (ii) prioritised vaccination campaign for females in the high-sexual-activity group, who may have a higher uptake under the optimal vaccine allocation strategy.

### 2.5 Mass vaccination strategies with limited vaccine supply

With current limited vaccine supply, we assumed fixed dose availability of 5%, 10%, 20%, or 30% of the population and evaluated different prioritisation strategies across age groups. A total of 10 vaccination strategies were proposed, targeting age groups prioritised in the optimal vaccine allocation strategies in 2.4. Within each set of prioritised age groups, vaccines were allocated by the following approaches:

A. Sequential vaccination: Vaccines were allocated to one group at a time until the maximum uptake threshold was reached before reallocating the remaining doses to the next priority group.
B. Equal vaccination: Vaccines were equally distributed across the selected age groups (≥ 2).

We also classified strategies based on whether they included or excluded childhood vaccination. The full list of vaccine strategies evaluated in this study is detailed below in the Results section (Section 3.3). The strategies were assessed through changes in the resulting *R_eff_*s.

All the analyses and visualizations were performed using the R software version 4.1.2 [29]. Analytical scripts are available at https://github.com/ShihuiJin/onetime_mpox_vaccination/tree/main.

## 3. Results

### 3.1 Projected effective reproduction number of Clade I MPXV in 47 sub-Saharan African countries

Without assuming mpox vaccination rollout, our model projected a gradual increase in *R_eff_* from 2025 to 2050 across the 47 sub-Saharan African countries. Minor-to-moderate geographical disparity was observed in the estimated *R_eff_*s across the 47 countries. Under the community-contact-only scenario, the interquartile range of the estimated *R_eff_*s across these countries was 0.14 at each time point between 2025 and 2050, whereas under the high-sexual-transmission setting, it remained below 0.02 throughout the same period (Fig 1).

**Fig 1.**
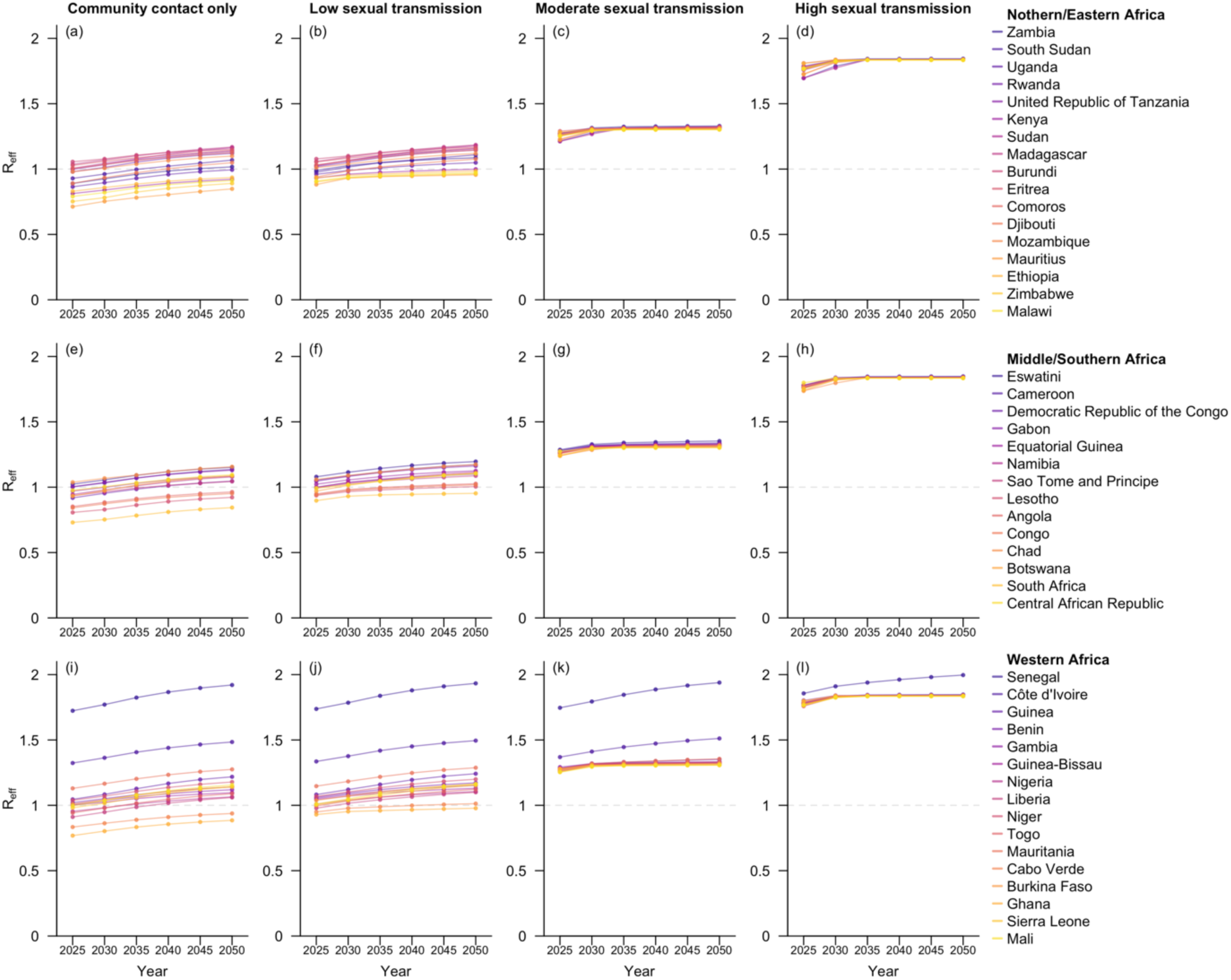
Projected *R_eff_* for the 47 sub-Saharan African countries at five-year intervals from 2025 to 2050. Countries within the same geospatial subregions are grouped in the same row and arranged in descending order by their estimated *R_eff_* in 2050. The four columns correspond to the four hypothetical scenarios with varying levels of sexual transmission.

In the scenario with community contact only, while the *R_eff_*s in 27 of these countries were projected to be lower than one in 2025, only 11 may remain so by 2050, mainly explained by the reduced smallpox-vaccine-induced population immunity as cohorts previously vaccinated against smallpox would age out of the population. The added route of sexual transmission did not qualitatively affect *R_eff_*in the low-sexual-transmission scenario. In contrast, in the moderate- or high-sexual-transmission scenarios, the projected *R_eff_* substantially exceeded one for all countries, mostly around 1.3 for the moderate-sexual-transmission setting and 1.8 for the high-sexual-transmission setting in 2025 (Fig 1, Fig S6).

### 3.2 Minimum vaccine coverage required for preventing epidemic growth

The estimated minimum vaccine coverage required to prevent epidemic growth (i.e., *R_eff_*< 1) increased substantially over time and with higher levels of sexual transmission. In the scenario with only community contact, 20 out of the 47 modelled countries would require vaccination in 2025, as the corresponding *R_eff_* was projected to be at least one. This number was projected to rise to 36 by 2050. However, in the high-sexual-transmission scenario, more closely representing the spread of Clade Ib in South Kivu, the DRC in 2024, all these sub-Saharan African countries would require a coverage of at least 14.5% (95% CI: 12.2%–17.4%) to prevent epidemic growth by 2025 and the number was projected to rise to 16.9% (95% CI: 12.6%–21.1%) by 2050. Particularly, among the 11 countries with documented local transmission Clade I MPXV as of December 2024 [23], the expected minimum coverage required in 2025 ranged from 0%–4.8% under the community-contact-only scenario to 14.5%–19.5% under the high-sexual-transmission scenario. By 2050, these coverage requirements were projected to increase to 0–10.8% and 22.6%–26.0%, respectively (Fig 2). The numbers of vaccine series required for each country in 2025 and 2050 are provided in the Supplementary Information (Table S4–S5)

**Fig 2.**
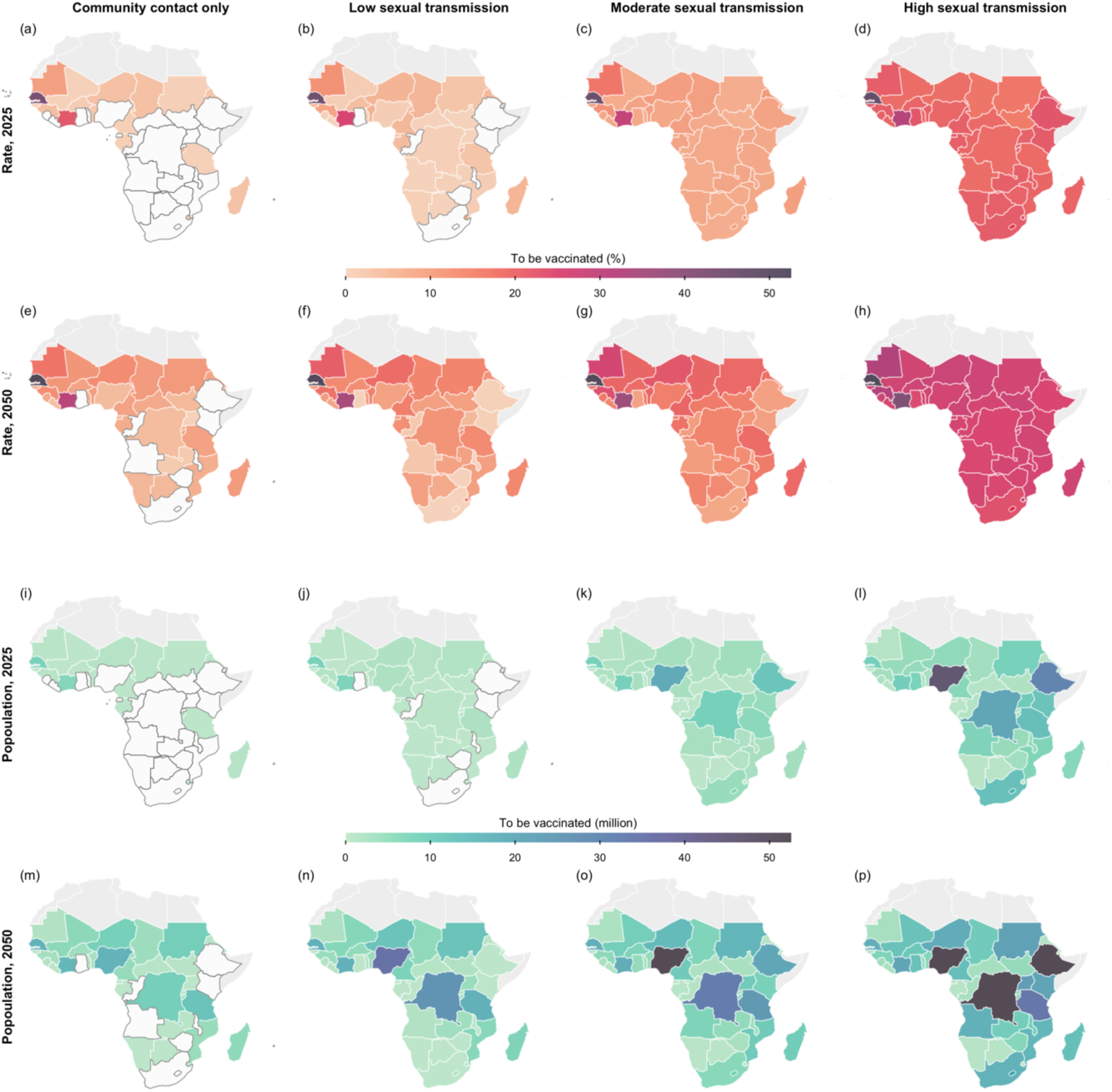
Projected minimum vaccine demand to prevent epidemic growth. Shown are the vaccine coverage (Row 1 and 2) and the number of individuals requiring vaccination (Row 3 and 4) to prevent epidemic growth in each sub-Saharan country modelled for 2025 (Row 1 and 3) and 2050 (Row 2 and 4), assuming the vaccine target group is defined only by age. The four columns correspond to the four hypothetical scenarios with varying levels of sexual transmission. Countries which do not require vaccination are coloured in white, with borders outlined in dark grey. The base map layer (boundaries of African countries) is sourced from Natural Earth (https://www.naturalearthdata.com), available under the Public Domain license (https://www.naturalearthdata.com/about/terms-of-use/).

Should children and young adults aged under 20 years be ineligible for vaccination, a surge in vaccine demand would be expected across countries in 2050 under scenarios with moderate or lower levels of sexual transmission, where the increase in coverage was estimated at roughly five percentage points [pp] when assuming community contact only. Nonetheless, this change was largely unsubstantial for most countries in 2025 (Fig S7–S8).

A significant reduction in the number of vaccine series required was observed when prioritising high-sexual-activity females. The impact of this prioritisation became more prominent under moderate or high levels of sexual transmission. In the high-sexual-transmission setting, for example, prioritising high-sexual-activity females aged 15–49 was projected to reduce the minimal coverage required by up to 14.7 pp (95% CI: 12.0pp– 17.2pp) in individual countries compared to scenarios without such prioritisation, while in the country projected to require the largest number vaccine series (Nigeria), this strategy was estimated save 25.3 (95% CI: 21.2–39.2) million vaccine series out of the original 41.5 (95% CI; 31.3–49.0) million expected in 2025. Particularly, the total number of vaccine series needed for the DRC, Burundi, and Uganda was estimated to decrease by 18.5 (95% CI: 15.6–22.2) million from 30.8 (95% CI: 23.4–36.4) million under the scenario without prioritising the high-sexual-activity females (Fig S9–S10).

Regarding the distribution of vaccines in the population under the optimal vaccine allocation strategies, age group prioritisation within countries requiring larger vaccination coverage was similar (Fig 3). With community contact only, the age group of 0–4 years generally required the highest vaccine coverage across time and space, followed by the 5–9 years group in a few countries including Senegal, Côte d’Ivoire and Mauritania by 2025.

**Fig 3.**
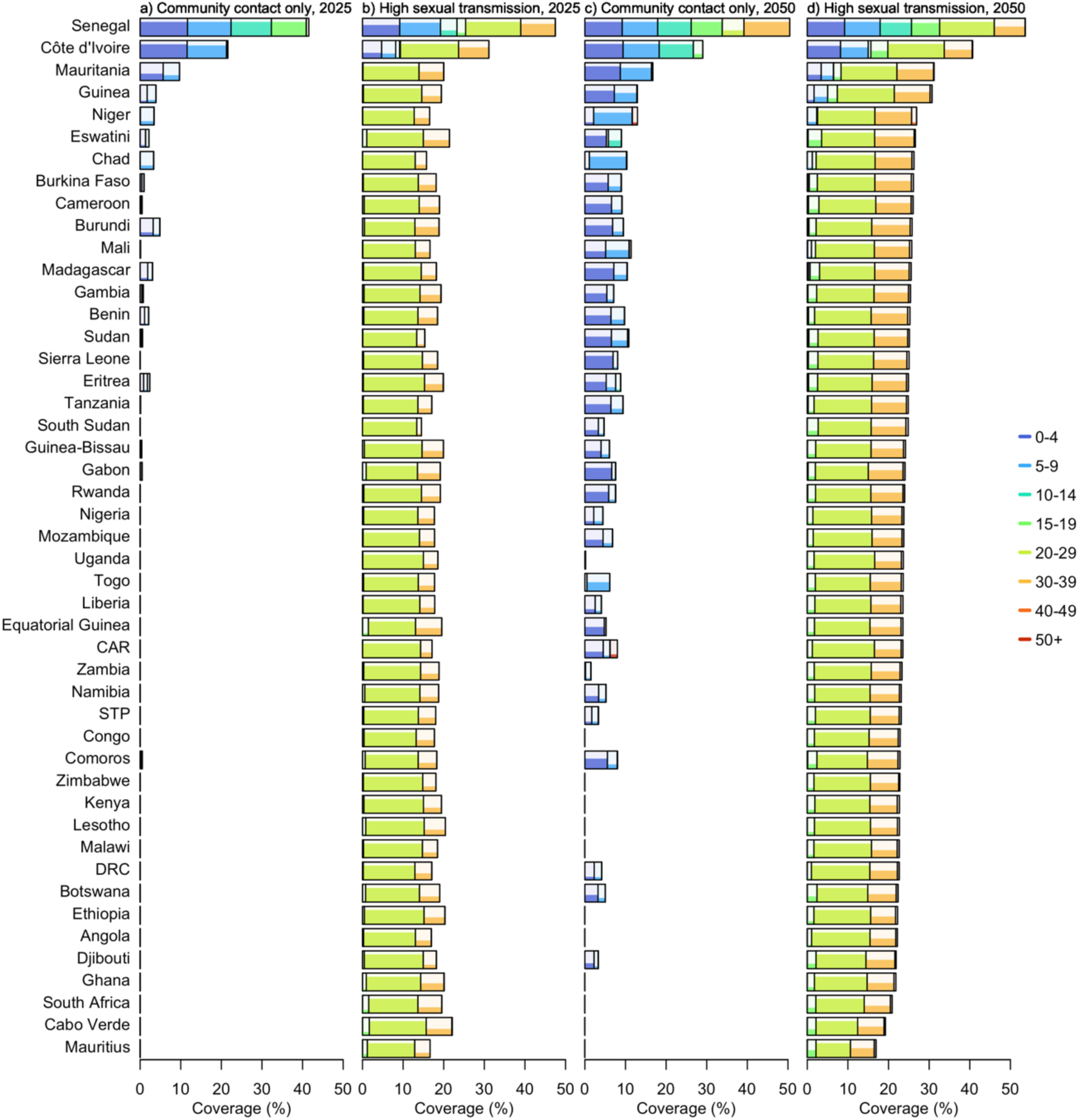
Optimal vaccine allocation strategy to prevent epidemic growth by age group across the 47 sub-Saharan African countries. Rectangles represent age groups allocated vaccines under the optimal strategies. The width of each rectangle indicates the average proportion of the general population (across 1000 simulations) which both belong to the corresponding age group and are receiving vaccination. The shading within each rectangle reflects the vaccine coverage level within that age group. A maximum coverage of 80% was applied to account for vaccine hesitancy. Countries are arranged in descending order by the projected minimum vaccine demand to reduce *R_eff_*to below 1 in 2050.

The projections revealed that the highest possible coverage rates among children aged under 10 years would be required for Senegal in particular as they have high household contact [30]. In total, 18 countries would require at least 1% coverage. By 2050, this number of countries would increase to 36. In the high-sexual-transmission scenario, the age group requiring the highest uptake rate shifted to 20–29 years, where the highest possible coverage (80%) would be necessary for each country to maximise the effectiveness of mass vaccination. The 30–39-year age group emerged as the second priority group, albeit with lower coverage levels ranging from 13% to 68% in 2025 and increasing to 42%–68% in 2050. The third group to target was 15–19 years, though minimal coverage was required for this age group to prevent epidemic growth in most countries (Fig 3).

### 3.3 Impacts of vaccination strategies assuming limited vaccine supply and sequential allocation

When determining the minimal vaccine coverage required, we allowed the algorithm to explore age-group prioritisation based on the calibrated parameter values. On the basis of the findings, sequential allocation strategies across prioritised age groups were explored assuming varying coverage levels in the population, as presented in Table 1.

**Table 1.** Fixed vaccination strategies assessed in this study.

| No. | Strategy |
| --- | --- |
| **A. Sequential vaccination** |  |
| A1 | Prioritising in the order of 0–4, 5–9, and 10–14 years |
| A2 | Prioritising in the order of 0–4, 20–29, and 30–39 years |
| A3 | Prioritising in the order of 20–29, 0–4, and 30–39 years |
| A4 | Prioritising in the order of 20–29, 15–19, and 30–39 years |
| A5 | Prioritising in the order of 20–29, 30–39, and 40–49 years |
| **B. Equal vaccination** |  |
| B1 | Uniform coverage across 0–15 years |
| B2 | Uniform coverage across 0–4 and 20–29 years |
| B3 | Uniform coverage across 20–39 years |

Compared to the equal distribution approaches (see Supplementary Information for details), the sequential allocation strategies were found to be equally or more effective. Under the community-contact-only scenario, prioritising 0–4 years was projected to lead to only three of the 47 countries being susceptible to epidemic growth in 2025, even at a 5% coverage of the overall population. Meanwhile, in the high sexual transmission scenario, prioritising individuals aged 20–29 years, followed by 30–39 years for remaining doses, could lower the median *R_eff_*across the 47 countries in 2025 to 1.28 with 10% vaccine coverage and further to 0.97 when the coverage doubled. A coverage rate of 20% was projected to suffice for preventing epidemic growth in most countries by 2025 where prioritising individuals aged 20–29 years, then 30–39 and 40–49 was estimated to lower the expected *R_eff_* of 38 countries among the 47 modelled to below one across all four hypothetical scenarios, increasing to 45 countries with 30% coverage (Fig 4, Fig S11).

**Fig 4.**
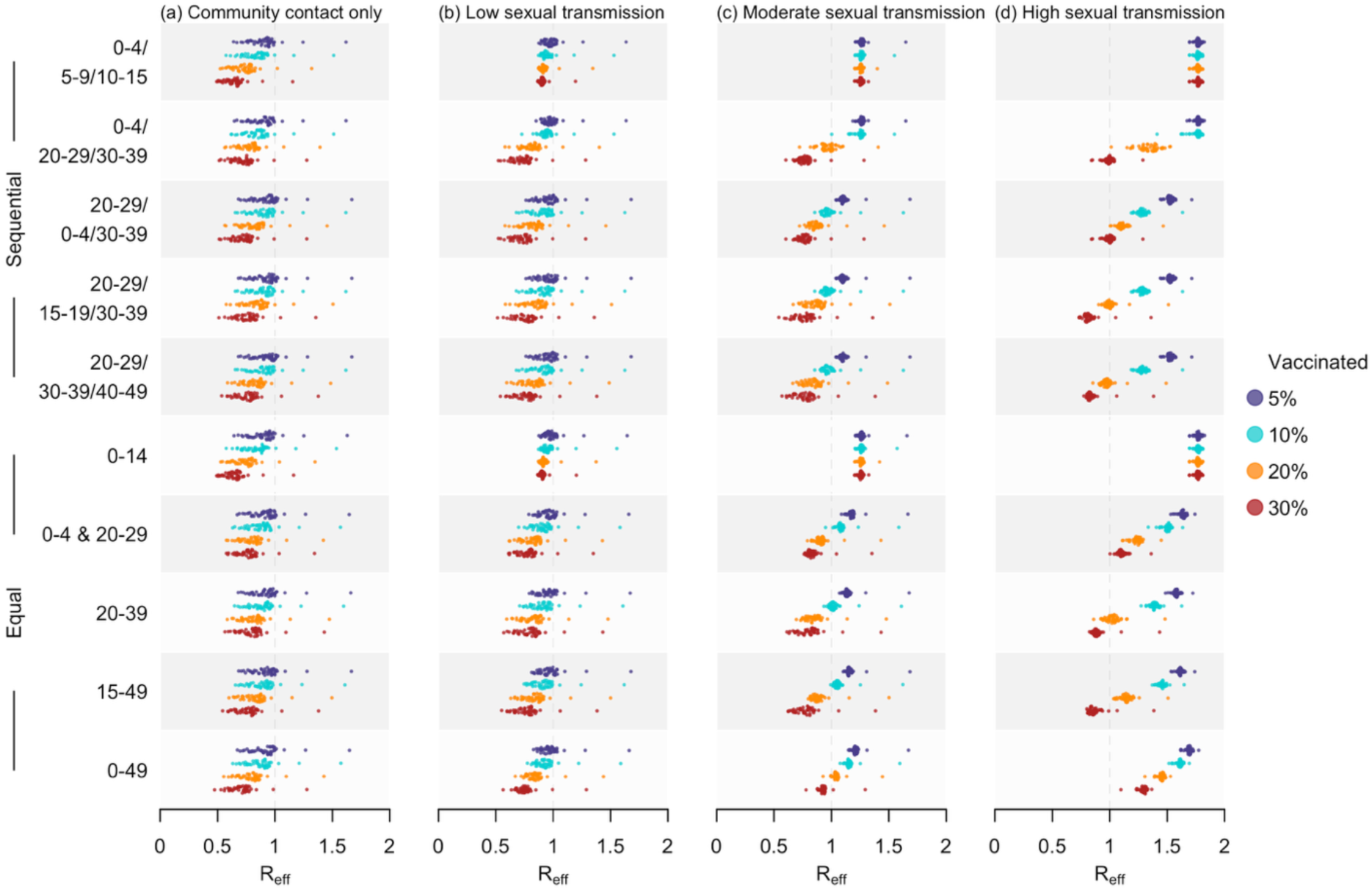
Estimated *R_eff_* in 2025 under diverse vaccination strategies. For each of the 47 countries, doses were allocated using two allocation methods, including sequential (first five rows) and equal (last five rows). Four coverage rates, including 5%, 10%, 20%, and 30%, were assessed, with outcomes summarised as shaded distributions in purple, blue, orange, and red, respectively. The four columns correspond to the four hypothetical scenarios with varying levels of sexual transmission.

The temporal variation in impacts of two vaccination strategies on reducing transmissibility is visualised in Fig 5, in which we focused on 11 countries affected by Clade I MPXV as of December 2024 [23]. Their effectiveness in reducing transmissibility, assuming fixed coverage, diminished over time, with the strategy prioritising children consistently outperforming the other in scenarios with community contact only or low sexual transmission. However, at 30% coverage, the adult-targeted strategy was the only one to maintain *R_eff_* below one across all four hypothetical scenarios in the 11 countries from 2025 to 2050 (Fig 5, Fig S12).

**Fig 5.**
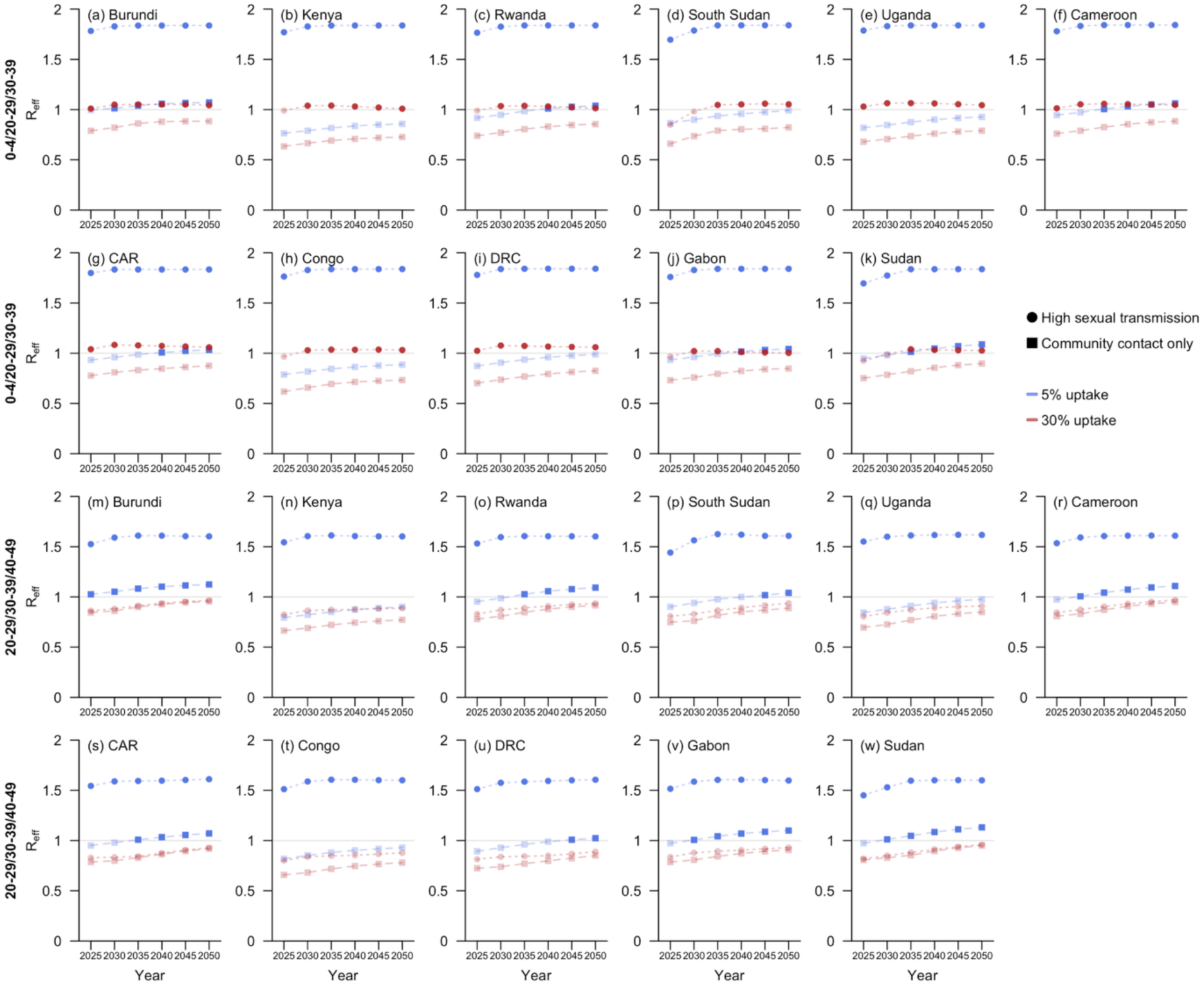
Projected *R_eff_* at five-year intervals from 2025 to 2050 under two sequential vaccination strategies for sub-Saharan African countries with documented local transmission of Clade I MPXV as of December 2024 [23]. The two vaccination strategies prioritised children aged 0–4 (Row 1 and 2) and young adults aged 20–29 (Row 3 and 4), respectively, with detailed allocation order across age groups presented in bold text along the left side of the figure. Two coverage rates, 5% (blue) and 30% (red), were assessed. Outcomes are presented as scattered rectangles for the hypothetical scenario with community contact only and as circles for the scenario with high sexual transmission. Data points corresponding to an expected *R_eff_* greater than one are highlighted in darker shades.

### 3.4 Effects of alternative model assumptions on mass vaccination effectiveness

Sensitivity analyses were conducted to project *R_eff_* and vaccine demand under alternative model assumptions. Generally, an increase in vaccine demand would be expected in scenarios with waning immunity induced by historical smallpox vaccination over time (Fig S17–S21), low mpox vaccine uptake rate among the targeted groups (Fig S30–S32), or suboptimal mpox vaccine effectiveness (Fig S33–S38). The demand also varied with changes in key transmission parameters of the next generation matrix model, such as the scale (Fig S26–S29) and the choice of social contact matrix used to approximate mixing patterns (Fig S14–S17). However, using all-contact matrices instead of home-contact matrices worsened model fits, potentially introducing error to the projections (Fig S13). Nonetheless, the prioritisation of age groups for vaccination in the case of limited vaccine supply remained largely consistent across scenarios. Please refer to the Supplementary Information for further details on the projected outcomes under these alternative scenarios.

## 4. Discussion

In this study, we developed a mathematical framework to quantify the future trajectory of the transmissibility of Clade I MPXV under different vaccination strategies. By integrating country-specific demographic profiles and contact patterns, we projected *R_eff_* for potential Clade I outbreaks across 47 sub-Saharan African countries between 2025 and 2050, under hypothetical scenarios with varying levels of sexual transmission and assuming a one-time mass vaccination campaign targeting specific age groups prior to the outbreak. The population-wide vaccine coverage necessary to prevent epidemic growth would range between 15%–20% in a typical sub-Saharan country, but would exceed 30% for countries with high community contact in 2025 under the scenario reflecting transmission dynamics of the 2024 Clade Ib outbreak in South Kivu, the DRC.

Our projections indicate the notable influence of diminishing population-level immunity from smallpox vaccination on the overall *R_eff_*, with an increase of up to 19% in individual countries over the projection window from 2025 to 2050. This decline in immunity led to a substantial rise in mpox vaccine demand. The sensitivity analysis suggests that the mpox vaccine demand could further increase if immunity induced by smallpox vaccines wanes over time (Fig S18–S21). Meanwhile, the growing population in many sub-Saharan African countries, standing at 1.24 billion people as of 2024 with nearly half of the individuals aged below 20 years in 2024, further intensifies vaccine needs. In the DRC, for instance, the minimum vaccine stockpile size required to prevent epidemic growth in 2050 was estimated to increase by over 150% compared to that in 2025 assuming a similar level of sexual transmission as that in the 2024 Clade Ib outbreak. This underscores the need to enhance vaccine production capacity and to train healthcare workers to ensure smooth vaccination rollouts.

The comparison among various mass vaccination strategies supports prioritising subpopulations with high transmission risks [31]. When sexual contact beyond general social interactions did not contribute to disease spread (the community-contact-only scenario), vaccinating children aged 0–4 years proved the most efficient in reducing *R_eff_*. This is mainly because, given the same contact rates, they were estimated to be 41% (95% CI: 12%–82%) more likely to become infected compared to older individuals. This finding aligns with the recommendation by Savinkina et al. to focus on vaccinating children aged under five years when resources are limited [17]. However, with an increase in the contribution of sexual transmission to overall transmission, the advantage of prioritising the 20–29 years cohort, the age group most likely to be highly sexually active, gradually surpassed that of targeting the 0–4 years cohort. This shift in the prioritisation of age groups underscores the importance of real-time monitoring of the age and gender composition of infections, as well as furthering our understanding on the transmission pathways of mpox. Detailed surveillance information could inform the current contribution of sexual transmission to overall disease spread, facilitating the adaptation of vaccine allocation strategy to match the prevailing transmission dynamics.

Under scenarios involving additional sexual transmission, prioritising high-sexual-activity females proved more effective than assuming uniform vaccine uptake within selected age groups (Fig S9–S10). Nonetheless, it should be noted that challenges exist in approaching this population due to social stigma, discrimination, and criminalization [32]. Furthermore, the transient nature of individuals within this group [33], in terms of the limited duration they remain at high sexual activity levels, may diminish the effectiveness of prioritising them, as those at high transmission risk during a future outbreak could belong to a new cohort with substantially lower vaccine coverage.

Uncertainty also exists regarding the mpox vaccine effectiveness for Clade I MPXV. In this study, we extrapolated effectiveness estimates from data on Clade IIb [12], assuming equivalent protection against Clade I. Nevertheless, projections under alternative vaccine effectiveness scenarios consistently substantiated prioritising either children under five years or young adults aged 20–29 (Fig S33–S38). We also assumed uniform effectiveness across different age groups due to the lack of age-specific data, with effectiveness set constant during the relatively short interval between vaccination and outbreak. As more age-specific effectiveness evidence becomes available, re-evaluation of these strategies will be necessary. In addition, as of December 2024, the safety and effectiveness of mpox vaccines for young children remain uncertain [34], especially those under five years old who are more susceptible to the disease. This uncertainty, along with misinformation, may contribute to vaccine hesitancy or refusal among caregivers [35]. Logistical challenges, such as vaccine storage, delivery, and regulatory processes required for childhood immunisation, may further complicate efforts to vaccinate young children at risk. Therefore, in scenarios where sexual transmission is limited, promoting mass vaccination among paediatric populations, as suggested by our results, will require careful assessment and clear public communication.

The highest achievable vaccine uptake rate emerges as an additional essential factor influencing the effectiveness of vaccination strategies. A key determinant is vaccine confidence levels among the target groups [36]. In this study, we assumed 80% acceptance rate based on studies on COVID-19 vaccines [28]. The sensitivity analysis testing a lower coverage cap of 70% [37] reveals a significantly higher overall vaccine demand to achieve similar intervention outcomes as more individuals from suboptimal age groups require vaccination (Fig S30–S32). This increase suggests the significance of enhancing disease awareness among key populations to boost vaccine confidence and ramp up uptake rates, especially in scenarios with constrained vaccine availability.

Our analysis modelled a one-time, proactive vaccination campaign implemented immediately before a potential outbreak, aiming to inform countries at immediate risk of importations and outbreaks due to ongoing transmission in neighbouring regions, as is the case for many sub-Saharan African countries in 2024 and 2025. The findings are also relevant to the earliest phase of an outbreak characterised by sporadic importations and limited local transmission, when infection-derived immunity remains negligible. However, it should be noted that the timing of such campaigns could influence their effectiveness. Initiating vaccination campaigns in advance, provided adequate vaccine production and delivery capacity, can reduce disease transmissibility not only during the immediate outbreak but also potentially over the long term, as mpox vaccines can confer protection lasting for decades, if not a lifetime [38]. Nonetheless, population-level immunity may wane over time due to factors such as declining vaccine-induced protection, the aging out of vaccinated individuals, and the continuous addition of unvaccinated young children. These dynamics suggest the potential need for periodic catch-up campaigns or routine immunisation, especially if outbreaks occur years after the initial campaign. These long-term strategies are not captured in our current one-time campaign model, but warrant further investigation to evaluate their feasibility, optimal timing, and effectiveness.

It is also worth highlighting that our model assumed community transmission of Clade I MPXV predominantly occurred within households [21]. The next generation matrix model in the main analysis included only the home contact components of the synthetic contact matrices. Although transmission in non-household settings, such as schools and nursing homes, might also contribute to community spread, evidence of large-scale spread in these institutions remained limited as of December 2024. Incorporating all contact components into the model resulted in a poorer fit to the observed age and sex distributions of cases in the DRC and consequently distinct projections in mpox vaccine demands (Fig S13–S17). This substantiates the plausibility of home contacts as a reasonable proxy of the mixing patterns which drive the Clade Ib MPXV transmission.

Furthermore, our model parameters were calibrated to and validated with the epidemiological data in the DRC (Fig S1), and extrapolated to 46 other sub-Saharan African countries. This approach implicitly presumes that transmission dynamics across age, sex, and sexual activity strata are sufficiently similar across settings. While the extrapolation harnessed the estimated country-specific community contact patterns, smallpox vaccination coverage, and demographic structures in literature [9,16,19,20,22], it did not account for potential biases in these estimated values and other contextual differences in social behaviours or economic factors. Nonetheless, the projected case demographics in Burundi, Uganda and Zambia, all of which experienced localised Clade Ib MPXV outbreaks in 2024–2025, closely matched the observed data and the estimates in a previous study [26], lending credibility to our modelling approach (Fig S2–S5).

Another limitation of this study pertains to the focus on localized human-to-human transmission. Zoonotic transmission and importation were not accounted for in the model due to the complexities of quantifying interactions between animals and humans, as well as between travellers and residents. Under the condition of *R_eff_*< 1, cases arising from zoonotic spillovers or introduced from foreign countries with subsequent human-to-human transmission may still be observed, although the outbreak is expected to be self-limiting. We also did not incorporate cross-protection from previous infections due to the limited number of reported cases relative to the overall population and the unclear age distribution [39]. In addition, the model was based on the simplified assumptions of equal contributions to secondary infections per unit in the community contact matrix and static contact patterns across age groups over time. Variations in secondary attack rates between contacts of different age groups or in different countries, together with potential behaviour changes, may introduce uncertainty to the long-term projections.

Despite the limitations, the flexible modelling framework established in our study facilitates application across a broad geographical range of 47 sub-Saharan African countries over an extended period from 2025 to 2050, enabling comparative analysis of regional variations in outbreak scales while highlighting the consistent effectiveness of pre-outbreak vaccination in curbing viral transmission. Our findings underscore the critical need for differentiated mass vaccination strategies to maximise the impact of limited vaccine supplies. Prioritising age groups with higher infection risks, specifically young children aged 0–4 years in scenarios with community contact only and adults aged 20–29 years in settings with high sexual transmission, can substantially reduce disease transmissibility. Nonetheless, the most effective approach should be customised to the evolving epidemiological landscape, informed by real-time surveillance data. This alignment between vaccination efforts and prevailing transmission patterns could facilitate better preparedness and even pre-emption of future mpox outbreaks at minimal cost.

## Abbreviations

MPXV: monkeypox virus
DRC: Democratic Republic of Congo
WHO: World Health Organization
CI: confidence interval
pp: percentage point.

## Ethics Approval

No ethics approval is needed for this study.

## Author contributions

S.J., A.E., and B.L.D. conceived and designed the study. S.J. implemented the statistical analysis and created the figures and tables. S.J. wrote the original draft of the manuscript. T.R.A., H.M., D.N., D.S.N., J.T.L., A.E., and B.L.D. reviewed and edited the manuscript.

## Data and code availability

Data and analytical codes utilized for this study are available at https://github.com/ShihuiJin/onetime_mpox_vaccination/tree/main.

## Acknowledgement

S.J. and B.L.D. are supported by the Ministry of Education Reimagine Research Grant and PREPARE, Ministry of Health. H.M., T.R.A., and A.E. are supported by the Japan Science and Technology Agency (JST) (JPMJPR22R3, to A.E.) and Japan Agency for Medical Research and Development (JP223fa627004). H.M. and A.E. are supported by the Japan Society for the Promotion of Science (JSPS) (JP22K17329, to A.E.). T.R.A. is also supported by the Rotary Foundation (GG2350294), the Nagasaki University World-leading Innovative & Smart Education (WISE) Program of the Japanese Ministry of Education, Culture, Sports, Science and Technology (MEXT) and the JSPS KAKENHI Grant-in-Aid for JSPS Fellows (JP24KJ1827). A.E. is also supported by National University of Singapore Start-Up Grant. J.T.L. is supported by Nanyang Technological University, Singapore – Imperial Research Collaboration Fund (INCF-2023-007). The funding sources are not involved in study design, in collection, analysis, and interpretation of data, in the writing of the report, or in the decision to submit the paper for publication.

