## Supplementary Information for "Vaccination strategies to achieve outbreak control for MPXV Clade I with a one-time mass campaign in sub-Saharan Africa: a scenario-based modelling study"

### Supplementary information: further explanations, results and sensitivity analyses

**Projecting the effective reproduction number**

*Summary of the next generation matrix model*

We used an existing model based on the next-generation matrix incorporating both community and sexual contact developed elsewhere [1] with minor adaptation and re-calibration. The model describes the transmission between individuals stratified age, sex and sexual activity level by through both social and sexual contacts in a form of a matrix whose eigenvector is expected to represent the age/sex distribution of mpox cases. Full details of the model settings and calibration approaches can be found in the original study.

The population is divided into eight age groups: 0–4, 5–9, 10–14, 15–19, 20–29, 30–39, 40–49, and 50+, and further stratified by sex and sexual activity level [1]. The next generation matrix is modelled as the product of group-specific susceptibility and a contact matrix incorporating both social (household) and sexual contact, described as below in the block-matrix format (each block is of size $8 \times 8$ to represent age group mixing):

$$\begin{pmatrix} D_{M_h} & O & O & O \\ O & D_M & O & O \\ O & O & D_{F_h} & O \\ O & O & O & D_F \end{pmatrix} \begin{pmatrix} O & O & S_{MF} & \Sigma_{MF}Q \\ C_{MM} & C_{MM} & C_{MF} & C_{MF} \\ S_{FM} & \Sigma_{FM}P & O & O \\ C_{FM} & C_{FM} & C_{FF} & C_{FF} \end{pmatrix}, \quad (1)$$

where $O$ is a zero matrix. Subscripts $M$ and $F$ represent males and females, respectively. The high-sexual-activity population, characterised by more frequent sexual activities and increased likelihood of acquiring or transmitting the virus through sexual contact, is limited to individuals aged 15–49 years. The parameters $p_a$ and $q_a$ represent the proportion high-sexual-activity among males and females of age group $a$, respectively. Namely, both parameters $p_a$ and $q_a$ are assumed to be zero outside the 15–49 age groups. $P = \mathrm{diag}(p_a)$ and $Q = \mathrm{diag}(q_a)$ are two diagonal matrices representing the high-sexual-activity proportions in the two subpopulations. The number of high-sexual-activity females is approximated by sex worker population, while for males, it is determined by the number engaging in commercial sex [1]. Group-specific susceptibility is represented by the block diagonal matrix in (1). It consists of four diagonal matrices, with $D_X = \mathrm{diag}(d_a)$ capturing the age-specific susceptibility within each subpopulation. These matrices are identical in the scenario with equal vaccine uptake rates across high- and low-risk groups and between sexes. They account for both increased susceptibility ($\sigma_c$) among children aged 0–4 years and reduced infection risk (by $e_s$) due to immunity conferred by prior smallpox vaccination, i.e.,

$$d_a = \begin{cases} \sigma_c(1 - e_s \cdot p_a^{vs}), 0 \le a < 5 \\ 1 - e_s \cdot p_a^{vs}, a \ge 5 \end{cases}, \quad (2)$$

where $p_a^{vs}$ is the smallpox vaccine coverage for age group $a$, as estimated by Taube et al. [2]. We assumed a constant level of smallpox vaccine effectiveness over time [3], but explored the potential waning of immunity [4] in a sensitivity analysis.

Both sexual and social contacts are incorporated into the block contact matrix in (1). $C_X$ represents the community contact matrix informed by existing datasets. We assumed the presence of heterosexual contacts beyond those captured by the community contact matrix, particularly among the high-sexual-activity individuals. To reflect behaviour heterogeneity across individuals with different sexual activity levels (i.e., those who are more sexually active are more likely to be sexually-acquired cases), different sexual transmission rates were assumed between from sexually-acquired cases (limited to high-sexual-activity individuals) and from community-acquired cases (applicable to the general community), denoted by sexual contact matrices $S_X$ and $\Sigma_X$, respectively.

For heterosexual contacts, we assumed a proportionate mixing pattern, with the elements of $S_{MF}$ and $S_{FM}$ modelled as $(S_{MF})_{ab} = w_F n_a p_a / \sum_i n_i p_i$ and $(S_{FM})_{ab} = w_M m_a q_a / \sum_i m_i q_i$, respectively. In these, $n_a$ and $m_a$ denote the number of males and females in age group $a$, while the parameters $w_F$ and $w_M$ represent the mean neighbour degrees (i.e., average number of sexual contacts) of sexually-acquired female and male cases, respectively, scaled relative to one unit of daily community contact. These two parameters are essential in determining the relative role of sexual transmission. We further presumed proportionality between $S_X$ and $\Sigma_X$, such that $\Sigma_{MF} = \alpha_F S_{MF}$ and $\Sigma_{FM} = \alpha_M S_{FM}$, where $\alpha_F$ and $\alpha_M$ are scaling factors informed by Inungu et al. [5], the sizes of high-sexual-activity populations, and the reciprocal nature of heterosexual contact, given by $\alpha_F w_F \sum_a m_a q_a = \alpha_M w_M \sum_a n_a p_a$. See Table S1–S2 for the full list of model parameters, and the original study [1] for a more detailed explanation on how they were incorporated into the next generation matrix model.

**Table S1**. Model parameters and data sources. Estimates are reported as means, with the corresponding 95% confidence intervals (CIs) shown in the parentheses.

| Parameter | Definition | Source/ Value |
|---|---|---|
| *Susceptibility* | | |
| $\sigma_c$ | Increased susceptibility among children aged 0–4 years, relative to older cohorts | Estimated, 1.41 (1.12–1.82) |
| $e_s$ | Effectiveness of smallpox vaccines | Estimated, 0.91 (0.84–0.97) |
| $e_m$ | Effectiveness of mpox vaccines | Pischel et al. [6] |
| $p_a^{vs}$ | Smallpox vaccine coverage rate for age group $a$ | Taube et al.[4] |
| *Contact* | | |
| $n_a$ | Population of males in age group $a$ | World Bank [7] |
| $m_a$ | Population of females in age group $a$ | World Bank [7] |
| $p_a^{male}$ | Proportion of males within age group $a$ | World Bank [7] |
| $p_a$ | Proportion of high-sexual-activity males within age group $a$ | Estimated[#] |
| $q_a$ | Proportion of high-sexual-activity females within age group $a$ | Estimated[#] |
| $C_X$ | Community contact matrix, approximated through synthetic home contact matrix; $X$ can be any of $MM$, $MF$, $FM$, $FF$, representing interactions within or between males and females | Prem et al. [8] |
| $w_F$, $w_M$ | Age-independent contribution to transmission of modelled sexual contact relative to one unit of daily home contact | Estimated, $w_F$: 13.4 (11.0–16.4) $w_M$: 9.3 (7.6–11.3) |
| $\alpha_F$, $\alpha_M$ | Ratio of number of sexual partners between community- and sexually-acquired infections | Inungu et al. [5], $\alpha_F$: 0.46 $\alpha_M$: $0.46 w_F \sum_a m_a q_a$ / $(w_M \sum_a n_a p_a)$ |
| $S_X$ | Sexual contact matrix for interactions between sexually-acquired, high-sexual-activity infections or exposures | Estimated[*] |
| $\Sigma_X$ | Sexual contact matrix for interactions between sexually-acquired and community-acquired high-sexual-activity infections or exposures | Estimated[*] |

[#] Presented in Table S2

[*] Constructed from other estimated parameters

**Table S2.** Estimated proportions (%, including means and 95% CIs) of high-sexual-activity individuals in the DRC in 2024.

| Age group ($a$) | $p_a$ | $q_a$ |
|---|---|---|
| 15–19 | 2.6 (1.1–4.0) | 2.0 (1.5–2.5) |
| 20–29 | 14.4 (12.8–16.2) | 3.9 (3.5–4.3) |
| 30–39 | 10.8 (8.8–12.8) | 0.98 (0.55–1.4) |
| 40–49 | 9.9 (6.3–13.7) | 0.22 (0–0.84) |

*Model adaptation and re-calibration*

In this study, for the community contact components $C_X$, we used synthetic contact matrix for the DRC in place of Zimbabwe contact survey data used in the original study [8]. According to the validation performance reported in the original study, we opted to use the home contact components of the synthetic contact matrix as opposed to all contact, but assessed the all-contact version in a subsequent sensitivity analysis. Since the only sex-aggregated synthetic contact matrix was available, we used country- and age group-specific sex ratios to split the contact matrix elements assuming proportionate mixing. We additionally assumed that the high-sexual-activity individuals constituted 10% of males and 2.1% of females aged between 15–49 years for the DRC [9,10]. To accommodate our adaptations to the model for the purpose of generalizability, we re-calibrated the model using the DRC outbreak datasets in the same process as the original study. Namely, we used the DRC synthetic contact matrix and estimated the susceptibility parameters $\sigma_c$ and $e_s$ from the Clade Ia outbreak datasets (2013–2017 data in Tshuapa province and 2024 data in Clade Ia-endemic provinces, the DRC), and the sexual-transmission-related parameters ($w_M$, $w_F$, $p_a$, $q_a$) from the Clade Ib outbreak dataset in North and South Kivu provinces, the DRC in 2024. We modelled the case counts, stratified by age and sex (where applicable), to follow a multinomial distribution, with the vector of probabilities proportional to the dominant eigenvector of the next generation matrix (1). Maximum likelihood estimates (MLEs) were then derived using the `optim` function in R software [11]. We obtained 1000 samples for parameter estimates using bootstrapping for convenience for the simulation.

*Model validation*

The effective reproduction number ($R_{eff}$) for Clade Ia transmission in the DRC in 2024 was estimated at 0.91 (95% CI: 0.90–0.92), while that for Clade Ib transmission was 1.76 (95% CI: 1.66–1.88). Both estimates fall within the range of the time-varying reproduction number previously reported by Marziano et al. [12]. In addition, the projected distribution of cases by age and sex, as presented in Fig S1, closely approximated the observed data for both clades.

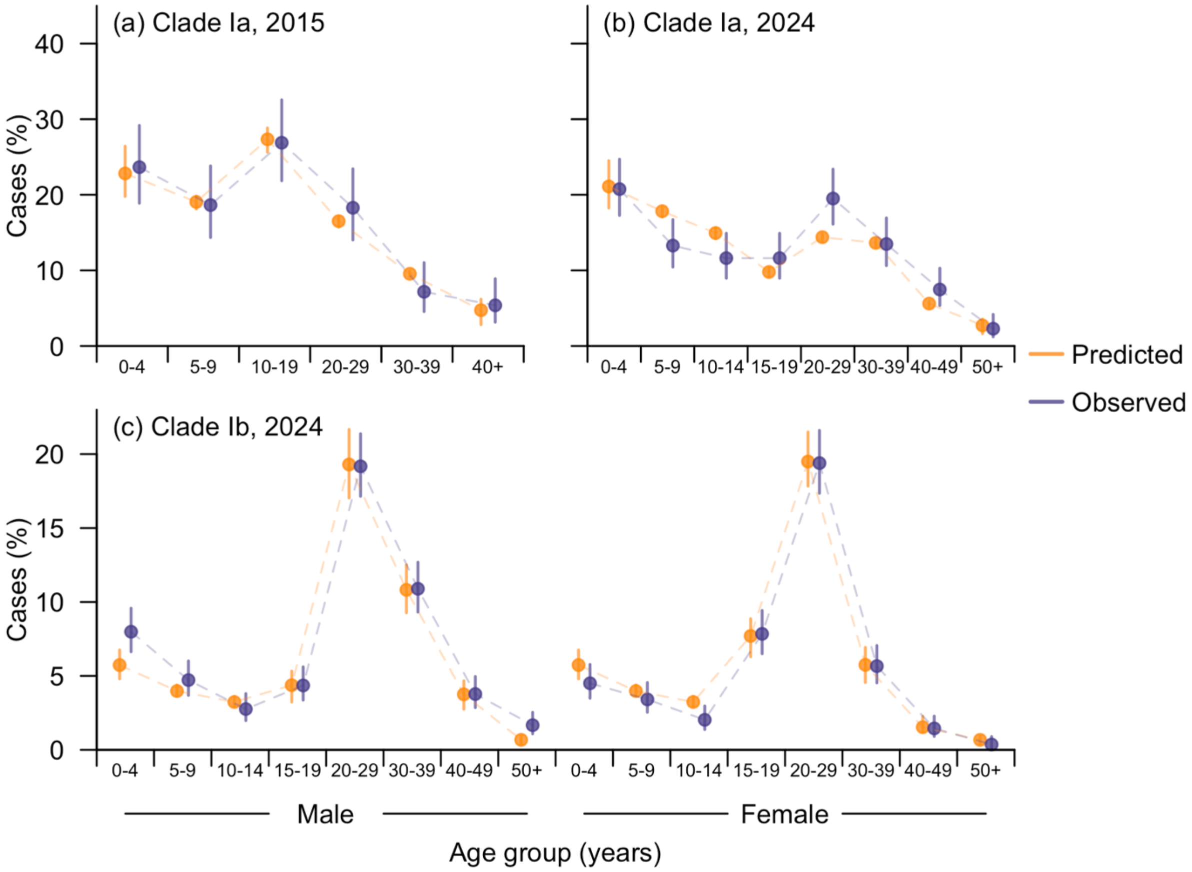

**Fig S1.** Fitted case demographics for the DRC. Subfigure (a) and (b) show the age distribution of case counts in endemic provinces in the DRC for the years 2015 and 2024, respectively, while subfigure (c) presents the age and sex distribution of cases counts in South Kivu, the DRC in 2024. Observed case demographics were obtained from Murayama et al [1].

*Application of the re-calibrated model to other sub-Saharan African countries*

To project the $R_{eff}$ for the 47 sub-Saharan African countries, we customised the next generation matrix using their respective synthetic home contact matrices [8]. The number of high-sexual-activity females in each country was approximated by the estimates of the sex worker population [9], while the number of high-sexual-activity males was computed by scaling the population to maintain the same ratio of high-sexual-activity females to males as the number for the DRC. The susceptibility of each age group was calculated based on the smallpox vaccine coverage rate by year of birth [2] and an assumed vaccine effectiveness of 80% (which we varied in a subsequent sensitivity analysis).

**Scenarios for varying sexual transmission levels**

The contribution of sexual contact to overall transmission was calculated by the sum of entries corresponding to sexually-acquired infections in the scaled dominant eigenvector.

This contribution was assumed to range between that observed in historical Clade Ia outbreaks in the endemic provinces and the 2024 Clade Ib outbreak in North and South Kivu provinces, the DRC, as preliminary findings from the original study suggested that sexual contact in Kamituga might contribute significantly to the apparent increase in $R_{eff}$ for Clade Ib in the South Kivu province [1]. In this high-sexual-transmission setting, sexual transmission accounted for approximately half of the total $R_{eff}$ (52%, 95% confidence interval [CI]: 48%–55%). For scenarios with low and moderate sexual transmission, we modified the degree of sexual contact contributions relative to home contact in populations of interest for males ($w_M$) by scaling it to 25% or 50% of the estimated value for the South Kivu setting in the DRC in 2024. These adjustments led to sexual contact contributing 9.2% (95% CI: 6.1%–13%) of $R_{eff}$ in the low-sexual-transmission scenario and 31% (95% CI: 26%–35%) in the moderate-sexual-transmission scenario (Table S3).

**Table S3.** Parameter values for the four hypothetical scenarios with varying levels of sexual transmission. The estimated contribution of sexual contact to overall transmission are reported as means, with the corresponding 95% CIs shown in the parentheses.

| Level of sexual transmission | Scaling factor for $w_M$ | Estimated contribution (%) of sexual contact to $R_{eff}$ |
|---|---|---|
| **No (Community contact only)** | 0 | 0 |
| **Low** | 0.25 | 9.1 (6.1–13) |
| **Moderate** | 0.5 | 31 (26–35) |
| **High** | 1 | 52 (48–55) |

*Projected transmission dynamics of Clade I MPXV in the absence of vaccination in 2024/early 2025*

The projected $R_{eff}$ for most of the 47 sub-Saharan African countries in 2024 ranged 0.8–2.0 (Fig S2), aligning with mpox transmissibility estimates reported in literature [13]. While the majority of the countries experienced only marginal impacts from Clade I MPXV, with limited documented local transmission in the general community, Burundi and Uganda were the most severely affected countries aside from the DRC as of 2024. Here, we visualised the projected age and sex distribution of cases in Burundi and Uganda under scenarios with varying levels of sexual transmission, and compared them with reported values from the initial stages of the outbreaks. As shown in Fig S3, moderate sexual transmission of Clade Ib MPXV in Burundi appears plausible. Under this assumed moderate-sexual-transmission scenario, the $R_{eff}$ calculated from the projected next generation matrix was 1.27 (95% CI: 1.20–1.35), consistent with findings by Otshudiema et al. [14]. In Uganda, projections suggest potentially moderate to high levels of sexual transmission, with $R_{eff}$ estimates being 1.28 (95% CI: 1.20–1.36) under the moderate-sexual-transmission scenario and 1.78 (95% CI: 1.67–1.90) under the high-sexual-transmission scenario (Fig S4).

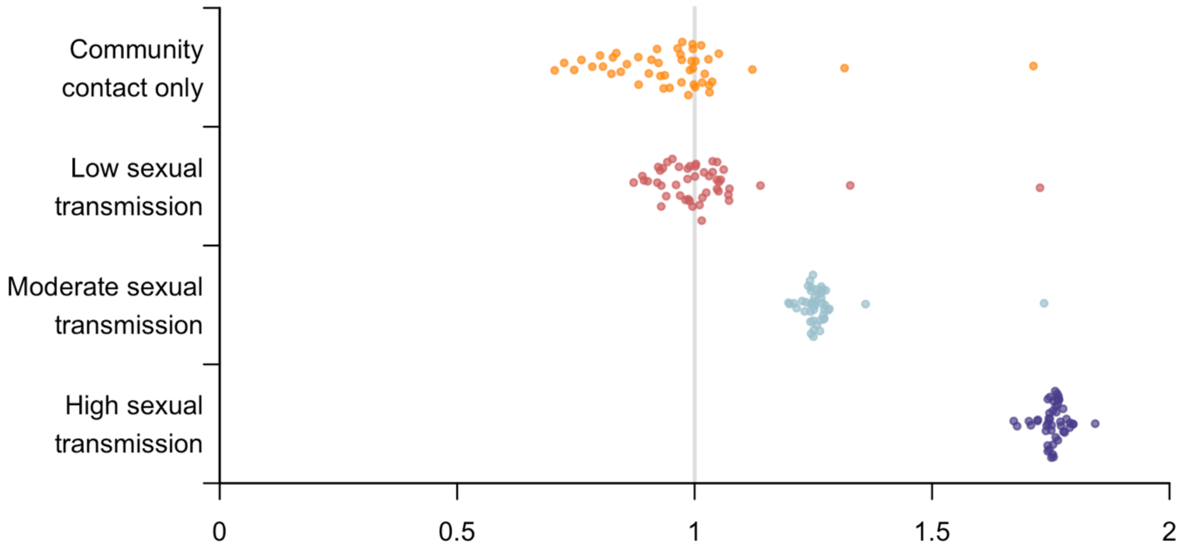

**Fig S2.** Projected mean $R_{eff}$ for the 47 sub-Saharan African countries in 2024.

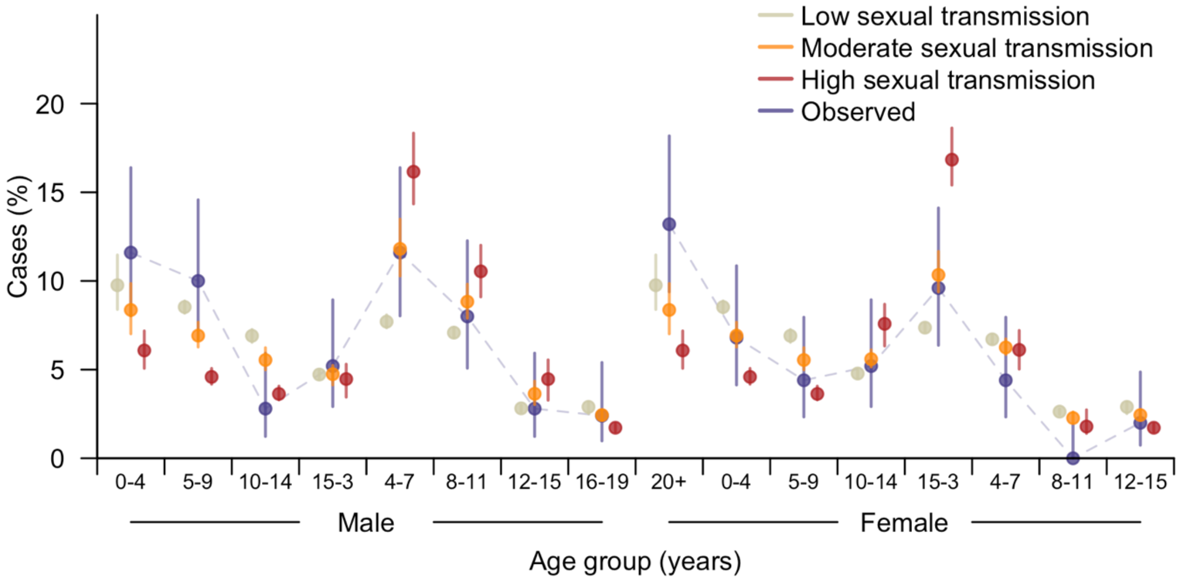

**Fig S3.** Projected and observed distribution of Clade Ib MPXV cases in Burundi in 2024, under the three scenarios assuming different levels of sexual transmission. The observed case demographics were obtained from the situation report published by the Burundi Ministry of Public Health on 27 August 2024 [1]. Projected means are represented by dots, with lines indicating the corresponding 95% CIs.

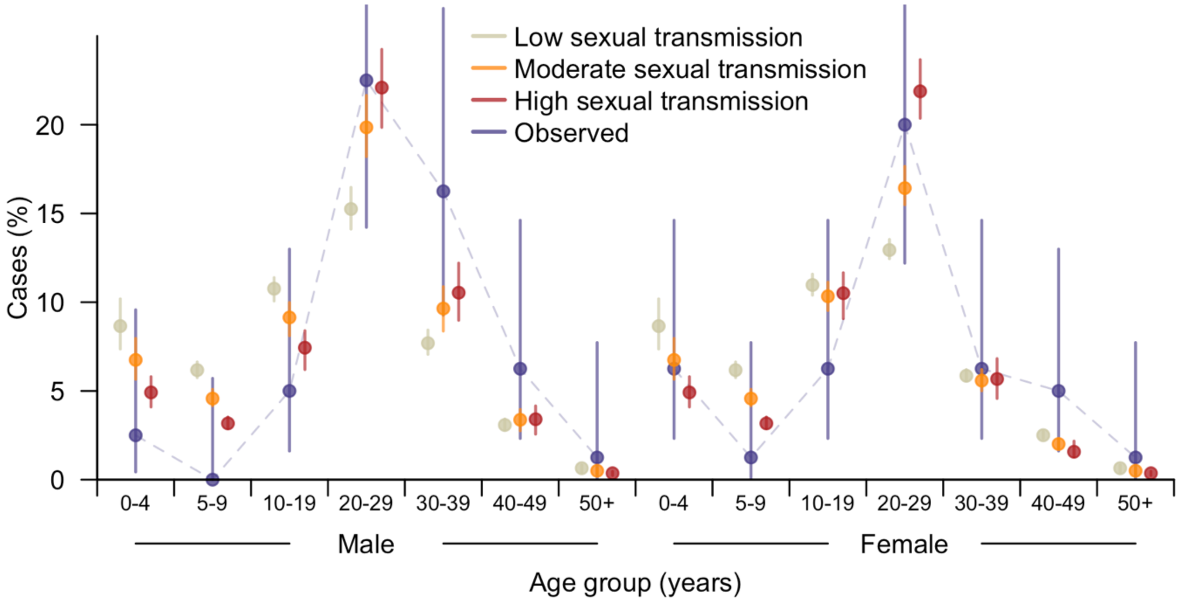

**Fig S4.** Projected and observed distribution of Clade Ib MPXV cases in Uganda in 2024, under the three scenarios assuming different levels of sexual transmission. The observed case demographics were obtained from the situation report published by World Health Organization Uganda on 21 October 2024 [15]. Projected means are represented by dots, with lines indicating the corresponding 95% CIs.

We also compared the age distribution of confirmed cases as of 28 March 2025 in Zambia against our model projections. Zambia was the only country aside from Burundi, and Uganda that had a Clade Ib MPXV-limited community outbreak and that disclosed case demographic data in situation reports as of April 2025 [16]. The country experienced a surge in Clade Ib cases in early 2025 and had not launched mpox vaccination campaigns [17]. Our projections indicate the plausibility of low to moderate levels of sexual transmission, with $R_{eff}$

estimates being 1.00 (95% CI: 0.97–1.04) under the low-sexual-transmission scenario and 1.28 (95% CI: 1.21–1.36) under the moderate-sexual-transmission scenario (Fig S5).

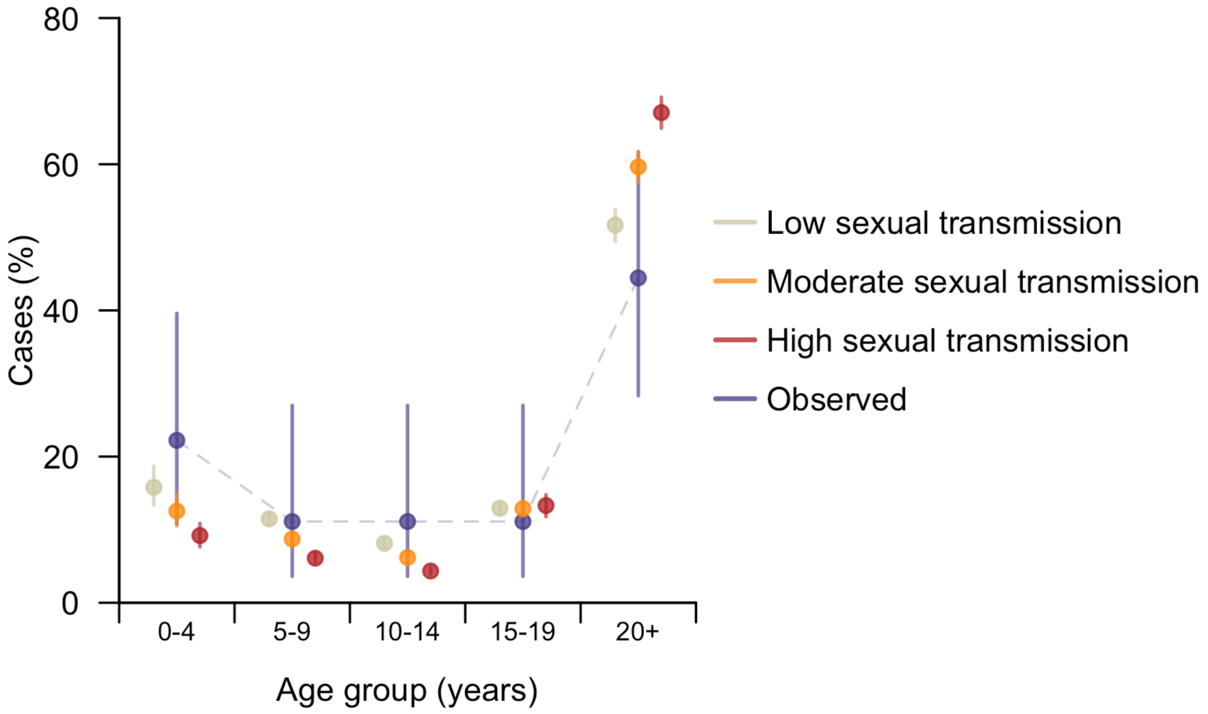

**Fig S5.** Projected and observed distribution of Clade Ib MPXV cases in Zambia in 2025, under the three scenarios assuming different levels of sexual transmission. The observed case demographics were obtained from the situation report published by Zambia National Public Health Institute on 28 March 2025 [16]. Projected means are represented by dots, with lines indicating the corresponding 95% CIs.

**Vaccination strategies**

*Age-specific susceptibility accounting for immunity from vaccines*

Let $ve_m$ denote the efficacy of mpox vaccines. The susceptibility of age group $a$ adjusted by pre-existing immunity from smallpox and mpox vaccination is

$$d_a = \begin{cases} (\sigma_c + 1 - e_s \cdot p_a^{vs})(1 - e_m \cdot p_a^{vm}), a \in \{0\text{–}4\} \\ (1 - e_s \cdot p_a^{vs})(1 - e_m \cdot p_a^{vm}), \text{otherwise} \end{cases}, \quad (3)$$

where $p_a^{vs}$ and $p_a^{vm}$ are vaccine coverage rates for smallpox and mpox vaccines, respectively.

*Determining the optimal vaccine allocation strategy*

We employed a stepwise gradient descent algorithm to estimate the minimal vaccine coverage rate required for outbreak prevention. Let $R_{eff} = f(v)$ be the function of the vaccine uptake rate vector $v$, with $v_0$ denoting the uptake rates prior to a new round of allocation (starting with a vector of zeros). In each round, up to $n$ individuals would be vaccinated, where $n$ was set to less than 1% of the national population. The allocation algorithm was implemented as follows:

i. The gradient $\nabla f(v_0)$ was computed and scaled by the inverse of the population size for each age group, to account for group-specific population differences.
ii. The group with the smallest scaled gradient which had not reached the maximum uptake threshold was selected as the target group. Vaccines were allocated to up to $n$ individuals within this group, while the uptake rate in this group would be kept below the maximum threshold.
iii. Step i and ii were repeated iteratively until the overall $R_{eff}$ fell below the threshold of 1.

*Vaccination strategies informed by the optimal allocation plans*

Under the scenario with limited vaccine supply, a total of 10 mass vaccination strategies were proposed and evaluated. Age groups to be prioritised in these strategies were selected based on the optimal vaccine allocation plans. For sequential vaccination strategies, the first targeted group in each strategy was chosen as the age group most frequently identified as having the highest coverage under at least one scenario, while subsequent groups were selected from those with non-zero coverage in at least one country and scenario. A maximum of three groups were included in each strategy. However, for equal vaccination strategies, at least two age groups were chosen on the basis of the estimated coverage in the optimal allocation strategy for each country.

According to the overall pattern shown in Fig 3, we assumed that individuals aged under 5 years old and/or those aged between 20 and 29 would be generally granted the highest priority, while a subset of other age groups under 50 years would also be deemed eligible for mpox vaccination (Table 1).

### Main analysis results

*Contribution of sexual contact to overall transmission*

Under the high-sexual-transmission scenario, the contribution of additional sexual contact to overall transmission was projected to range from 40% to 61% for over 90% of the sub-Saharan African countries in 2025. However, under the assumption of fixed sexual contact patterns over time, this contribution would decline to 34%–55% for these countries by 2050 (Fig S6). This decrease in the relative contribution of sexual contact can be explained by the waning population immunity from smallpox vaccination, which would increase the susceptibility of older age groups (particularly individuals over 45 years old) who were not assumed to primarily contribute to sexual contact, thereby elevating risk of infection through community contact.

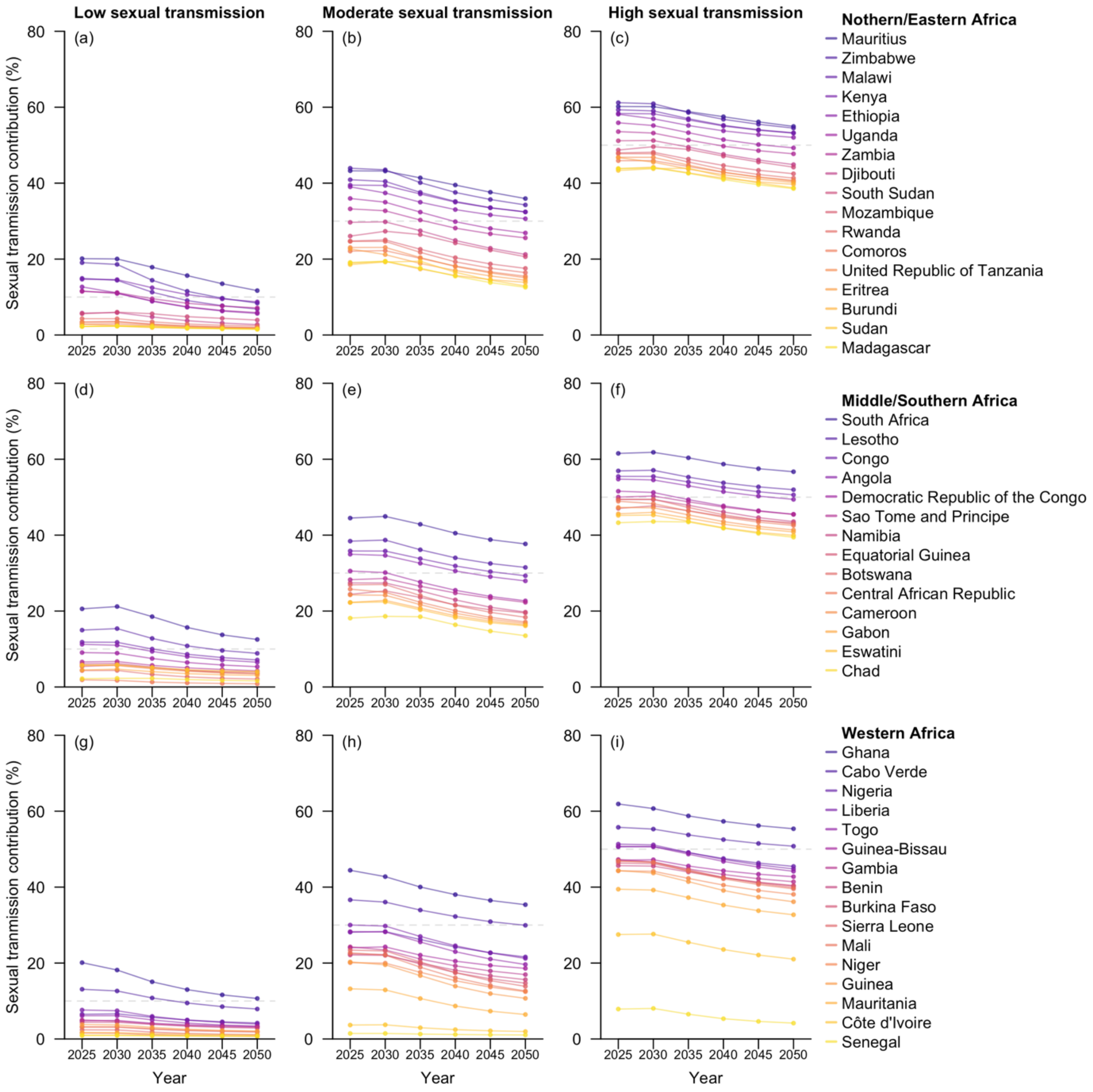

**Fig S6**. Projected contribution of sexual contact to overall transmission for the 47 sub-Saharan African countries at five-year intervals from 2025 to 2050. Countries within the same geospatial subregions are grouped in the same row and arranged in descending order by their estimated contribution in 2050. The four columns correspond to the four hypothetical scenarios with varying levels of sexual transmission.

*Minimum number of vaccine series required to prevent epidemic growth*

Given the relatively large population sizes in many sub-Saharan African countries, the number of individuals to be vaccinated could be substantial. Under the high-sexual-transmission scenario, vaccinating the DRC alone, with an estimated population size of 109 million in 2025, would require 17.1% (95% CI: 13.2%–20.2%) of the population being vaccinated, equivalent to 18.6 (95% CI: 14.4–22.0) million vaccine series in 2025. If the mass vaccination campaign were to roll out in 2050, an additional 2.9 (95% CI: 1.6–5.0) million vaccine series would be necessary to account for population growth and diminishing population immunity from smallpox vaccination. To further prevent epidemic growth in Burundi and Uganda–two other countries with local transmission of Clade Ib MPXV as of November 2024 [18]–an estimated total of 30.8 (95% CI: 23.4–36.4) million vaccine series would be needed in 2025. When expanding the target to include all the sub-Saharan African countries with clusters of Clade Ib cases as of November 2024 [18], including the DRC, Burundi, Kenya, Rwanda, and Uganda, up to 44.7 (95% CI: 34.0–53.0) million individuals would require immunisation to prevent epidemic growth in these countries in 2025. This number would increase to 62.5 (95% CI: 48.1–74.5) million for the 11 countries with documented local transmission of Clade I MPXV as of December 2024 (Burundi, Cameroon, Central African Republic, the DRC, Gabon, Kenya, Republic of the Congo, Rwanda, South Sudan, Sudan, and Uganda) [18], further to 146 (95% CI: 123–170) million by 2050. However, the vaccine burden would be greatly alleviated if the circulating MPXV strain(s) exhibited low levels of sexual transmission. Under the community-contact-only scenario, four of the 11 countries affected by Clade I MPXV would require vaccination, leading to a total demand of 1.1 (95% CI: 0.5–1.9) million vaccine series in 2025. Nonetheless, this number was projected to raise significantly to 29.0 (95% CI: 19.7–38.7) million in 2050 mainly due to the loss of protection from historical smallpox vaccination campaigns, with the DRC alone accounting for 9.0 (95% CI: 5.0–13.0) million of these vaccine series (Fig 2, Table S4–S5).

**Table S4.** Project minimum number of vaccine series required to prevent epidemic growth for the 47 sub-Saharan African countries in 2025. The countries are arranged in descending order by the number of vaccine series required under the high-sexual-transmission scenario.

| Country | Community contact only (mean, 95%CI) | Low sexual transmission (mean, 95%CI) | Moderate sexual transmission (mean, 95%CI) | High sexual transmission (mean, 95%CI) |
|---|---|---|---|---|
| **Nigeria** | 0.00 (0.00-0.00) | 2.16 (0.00-5.00) | 19.6 (15.5-23.5) | 41.5 (32.1-49.0) |
| **Ethiopia** | 0.00 (0.00-0.00) | 0.00 (0.00-0.00) | 11.9 (9.00-14.5) | 27.0 (21.0-32.0) |
| **Democratic Republic of the Congo** | 0.00 (0.00-0.00) | 0.65 (0.00-2.00) | 8.87 (7.00-11.0) | 18.6 (14.4-22.0) |
| **United Republic of Tanzania** | 0.08 (0.00-0.45) | 1.89 (1.00-2.90) | 5.89 (4.70-6.95) | 12.2 (9.55-14.6) |
| **South Africa** | 0.00 (0.00-0.00) | 0.00 (0.00-0.00) | 4.45 (3.15-5.65) | 12.0 (9.24-14.3) |
| **Kenya** | 0.00 (0.00-0.00) | 0.00 (0.00-0.00) | 4.73 (3.50-5.85) | 11.2 (8.49-13.3) |
| **Uganda** | 0.00 (0.00-0.00) | 0.01 (0.00-0.05) | 4.35 (3.35-5.31) | 9.51 (7.50-11.3) |
| **Côte d'Ivoire** | 6.55 (5.90-7.10) | 7.46 (6.75-8.10) | 8.79 (8.30-9.10) | 9.44 (9.10-9.85) |
| **Senegal** | 7.76 (7.50-8.00) | 8.27 (7.95-8.58) | 8.48 (8.20-8.80) | 8.87 (8.58-9.10) |
| **Sudan** | 0.28 (0.00-0.65) | 1.30 (0.75-1.95) | 3.66 (2.90-4.45) | 7.77 (6.25-9.55) |
| **Ghana** | 0.00 (0.00-0.00) | 0.00 (0.00-0.00) | 2.95 (2.20-3.65) | 7.13 (5.55-8.55) |
| **Angola** | 0.00 (0.00-0.00) | 0.00 (0.00-0.00) | 2.85 (2.15-3.55) | 6.60 (5.03-7.85) |
| **Mozambique** | 0.00 (0.00-0.00) | 0.38 (0.00-0.85) | 2.96 (2.30-3.55) | 6.36 (4.94-7.55) |
| **Madagascar** | 0.97 (0.60-1.30) | 1.79 (1.25-2.50) | 3.20 (2.80-3.60) | 5.78 (4.55-6.85) |
| **Cameroon** | 0.13 (0.00-0.40) | 1.58 (1.20-1.90) | 2.95 (2.50-3.40) | 5.71 (4.55-6.70) |
| **Niger** | 1.00 (0.80-1.20) | 1.70 (1.40-2.50) | 2.99 (2.65-3.30) | 4.85 (3.95-5.70) |
| **Burkina Faso** | 0.24 (0.00-0.45) | 0.97 (0.60-1.40) | 2.21 (1.85-2.60) | 4.43 (3.45-5.20) |
| **Mali** | 0.02 (0.00-0.10) | 0.25 (0.00-0.50) | 1.85 (1.40-2.25) | 4.12 (3.20-4.90) |
| **Zambia** | 0.00 (0.00-0.00) | 0.06 (0.00-0.35) | 1.89 (1.50-2.30) | 4.09 (3.15-4.80) |
| **Malawi** | 0.00 (0.00-0.00) | 0.00 (0.00-0.00) | 1.74 (1.30-2.15) | 4.07 (3.15-4.90) |
| **Zimbabwe** | 0.00 (0.00-0.00) | 0.00 (0.00-0.00) | 1.35 (1.00-1.70) | 3.14 (2.50-3.80) |
| **Chad** | 0.64 (0.50-0.80) | 1.04 (0.90-1.20) | 1.92 (1.70-2.10) | 3.06 (2.50-3.60) |
| **Guinea** | 0.57 (0.40-0.70) | 1.32 (0.90-1.70) | 1.85 (1.70-2.00) | 2.89 (2.40-3.30) |
| **Rwanda** | 0.00 (0.00-0.00) | 0.30 (0.10-0.50) | 1.32 (1.10-1.60) | 2.83 (2.20-3.35) |
| **Benin** | 0.30 (0.20-0.40) | 0.94 (0.70-1.10) | 1.46 (1.30-1.70) | 2.67 (2.15-3.10) |
| **Burundi** | 0.68 (0.50-0.80) | 1.05 (0.80-1.40) | 1.63 (1.40-1.80) | 2.63 (2.15-3.07) |
| **Sierra Leone** | 0.00 (0.00-0.00) | 0.04 (0.00-0.14) | 0.75 (0.57-0.92) | 1.70 (1.31-2.03) |
| **Togo** | 0.00 (0.00-0.00) | 0.02 (0.00-0.11) | 0.75 (0.58-0.92) | 1.68 (1.29-2.00) |
| **South Sudan** | 0.00 (0.00-0.00) | 0.00 (0.00-0.00) | 0.77 (0.60-1.00) | 1.67 (1.40-2.00) |
| **Congo** | 0.00 (0.00-0.00) | 0.00 (0.00-0.00) | 0.47 (0.35-0.59) | 1.13 (0.86-1.34) |
| **Central African Republic** | 0.00 (0.00-0.00) | 0.00 (0.00-0.04) | 0.48 (0.37-0.59) | 1.04 (0.84-1.26) |
| **Mauritania** | 0.50 (0.41-0.56) | 0.63 (0.52-0.71) | 0.90 (0.86-0.94) | 1.03 (0.95-1.13) |
| **Liberia** | 0.00 (0.00-0.00) | 0.00 (0.00-0.03) | 0.45 (0.34-0.56) | 1.01 (0.78-1.20) |
| **Eritrea** | 0.09 (0.04-0.14) | 0.19 (0.14-0.31) | 0.43 (0.36-0.49) | 0.77 (0.62-0.92) |

| | | | | |
|---|---|---|---|---|
| **Gambia** | 0.02 (0.00-0.05) | 0.15 (0.10-0.20) | 0.29 (0.24-0.34) | 0.56 (0.45-0.66) |
| **Botswana** | 0.00 (0.00-0.00) | 0.00 (0.00-0.00) | 0.21 (0.16-0.27) | 0.53 (0.40-0.63) |
| **Namibia** | 0.00 (0.00-0.00) | 0.01 (0.00-0.04) | 0.22 (0.17-0.27) | 0.50 (0.38-0.60) |
| **Lesotho** | 0.00 (0.00-0.00) | 0.00 (0.00-0.00) | 0.20 (0.15-0.25) | 0.49 (0.37-0.58) |
| **Gabon** | 0.01 (0.00-0.04) | 0.11 (0.07-0.15) | 0.23 (0.19-0.27) | 0.49 (0.38-0.57) |
| **Guinea-Bissau** | 0.01 (0.00-0.03) | 0.10 (0.06-0.14) | 0.23 (0.19-0.26) | 0.45 (0.35-0.53) |
| **Equatorial Guinea** | 0.00 (0.00-0.00) | 0.03 (0.01-0.06) | 0.15 (0.12-0.18) | 0.35 (0.28-0.41) |
| **Eswatini** | 0.03 (0.01-0.04) | 0.10 (0.07-0.12) | 0.15 (0.12-0.17) | 0.26 (0.22-0.31) |
| **Djibouti** | 0.00 (0.00-0.00) | 0.00 (0.00-0.00) | 0.09 (0.06-0.11) | 0.21 (0.17-0.26) |
| **Mauritius** | 0.00 (0.00-0.00) | 0.00 (0.00-0.00) | 0.08 (0.06-0.10) | 0.21 (0.15-0.25) |
| **Comoros** | 0.00 (0.00-0.01) | 0.03 (0.01-0.04) | 0.07 (0.06-0.09) | 0.16 (0.13-0.19) |
| **Cabo Verde** | 0.00 (0.00-0.00) | 0.00 (0.00-0.00) | 0.05 (0.04-0.07) | 0.13 (0.10-0.16) |
| **Sao Tome and Principe** | 0.00 (0.00-0.00) | 0.00 (0.00-0.00) | 0.02 (0.01-0.02) | 0.04 (0.03-0.05) |

**Table S5.** Project minimum number of vaccine series required to prevent epidemic growth for the 47 sub-Saharan African countries in 2050. The countries are arranged in descending order by the number of vaccine series required under the high-sexual-transmission scenario.

| Country | Community contact only (mean, 95%CI) | Low sexual transmission (mean, 95%CI) | Moderate sexual transmission (mean, 95%CI) | High sexual transmission (mean, 95%CI) |
|---|---|---|---|---|
| **Nigeria** | 16.7 (11.0-21.5) | 35.8 (26.0-46.0) | 59.2 (53.5-65.0) | 89.7 (76.5-105.0) |
| **Democratic Republic of the Congo** | 8.96 (5.00-13.0) | 26.4 (18.0-32.5) | 32.3 (27.5-36.0) | 49.2 (42.0-57.0) |
| **Ethiopia** | 0.00 (0.00-0.00) | 0.31 (0.00-3.00) | 20.6 (16.0-25.5) | 47.7 (37.5-58.0) |
| **United Republic of Tanzania** | 12.2 (9.00-15.0) | 17.1 (13.5-21.0) | 24.7 (22.0-27.0) | 32.3 (29.0-36.0) |
| **Sudan** | 9.11 (6.90-10.9) | 12.2 (9.55-14.7) | 16.2 (14.9-17.6) | 21.3 (19.3-23.5) |
| **Côte d'Ivoire** | 14.9 (13.3-16.2) | 16.7 (15.3-18.0) | 18.4 (17.3-19.5) | 20.9 (19.4-22.3) |
| **Uganda** | 0.15 (0.00-1.00) | 4.43 (2.05-6.85) | 10.2 (8.35-12.2) | 20.7 (16.6-24.9) |
| **Kenya** | 0.00 (0.00-0.00) | 0.39 (0.00-1.90) | 8.29 (6.40-10.3) | 19.3 (15.2-23.8) |
| **Niger** | 8.68 (7.50-9.65) | 12.0 (10.3-13.8) | 14.8 (13.6-15.9) | 18.0 (16.9-19.7) |
| **Senegal** | 16.4 (15.6-17.1) | 16.9 (16.2-17.6) | 17.0 (16.3-17.7) | 17.5 (16.9-18.1) |
| **Angola** | 0.00 (0.00-0.00) | 1.59 (0.05-3.00) | 7.46 (5.90-9.05) | 16.1 (12.8-19.3) |
| **South Africa** | 0.00 (0.00-0.00) | 0.01 (0.00-0.21) | 6.14 (4.60-7.70) | 15.4 (11.7-18.8) |
| **Mozambique** | 4.29 (3.05-5.45) | 6.19 (4.70-7.80) | 10.4 (9.10-11.6) | 15.0 (13.0-17.5) |
| **Cameroon** | 4.69 (3.50-5.85) | 7.59 (6.05-9.30) | 10.1 (9.24-11.1) | 13.4 (11.9-14.8) |
| **Madagascar** | 5.40 (4.00-6.55) | 7.20 (5.65-8.80) | 9.80 (8.75-10.8) | 13.2 (12.0-14.7) |
| **Mali** | 5.40 (4.25-6.35) | 6.34 (5.05-7.65) | 9.27 (8.35-10.1) | 12.2 (11.2-13.2) |
| **Ghana** | 0.00 (0.00-0.00) | 0.06 (0.00-0.65) | 4.78 (3.65-5.95) | 11.4 (8.94-14.0) |
| **Burkina Faso** | 3.63 (2.80-4.40) | 5.11 (3.95-6.35) | 7.55 (6.75-8.35) | 10.6 (9.60-11.8) |
| **Chad** | 3.77 (3.30-4.30) | 5.13 (4.40-6.00) | 7.19 (6.50-7.80) | 9.58 (8.75-10.4) |
| **Zambia** | 0.56 (0.00-1.15) | 3.48 (2.45-4.50) | 4.93 (4.15-5.75) | 8.73 (7.15-10.4) |
| **Malawi** | 0.00 (0.00-0.00) | 0.02 (0.00-0.35) | 3.56 (2.70-4.40) | 8.41 (6.59-10.3) |
| **Guinea** | 3.07 (2.50-3.50) | 4.21 (3.55-5.00) | 5.54 (5.05-5.95) | 7.28 (6.50-7.95) |
| **Benin** | 2.46 (1.90-2.95) | 3.71 (3.00-4.40) | 4.98 (4.65-5.35) | 6.38 (5.80-7.10) |
| **Burundi** | 2.30 (1.80-2.80) | 3.09 (2.40-3.85) | 4.50 (4.00-5.00) | 6.25 (5.70-7.00) |
| **Zimbabwe** | 0.00 (0.00-0.00) | 0.01 (0.00-0.15) | 2.52 (1.90-3.10) | 6.03 (4.70-7.35) |
| **Rwanda** | 1.74 (1.25-2.25) | 2.61 (1.95-3.30) | 3.97 (3.50-4.40) | 5.52 (4.90-6.40) |
| **South Sudan** | 0.82 (0.50-1.10) | 1.69 (1.20-2.30) | 2.58 (2.28-2.95) | 4.34 (3.50-5.20) |
| **Togo** | 0.95 (0.70-1.10) | 1.61 (1.30-1.90) | 2.54 (2.30-2.80) | 3.66 (3.20-4.25) |
| **Sierra Leone** | 1.10 (0.80-1.40) | 1.40 (1.01-1.80) | 2.29 (2.00-2.60) | 3.41 (3.00-3.90) |
| **Mauritania** | 1.49 (1.25-1.70) | 1.79 (1.56-2.07) | 2.28 (2.08-2.46) | 2.78 (2.57-3.03) |
| **Central African Republic** | 0.92 (0.65-1.12) | 1.12 (0.80-1.40) | 1.89 (1.65-2.10) | 2.71 (2.30-3.20) |
| **Congo** | 0.00 (0.00-0.00) | 0.19 (0.00-0.40) | 1.07 (0.85-1.30) | 2.37 (1.90-2.90) |
| **Liberia** | 0.36 (0.21-0.50) | 0.79 (0.55-1.08) | 1.28 (1.11-1.44) | 2.09 (1.72-2.51) |
| **Eritrea** | 0.52 (0.38-0.65) | 0.72 (0.54-0.92) | 1.07 (0.94-1.19) | 1.48 (1.33-1.71) |
| **Gambia** | 0.33 (0.24-0.42) | 0.54 (0.41-0.67) | 0.83 (0.74-0.92) | 1.19 (1.06-1.36) |
| **Gabon** | 0.28 (0.21-0.37) | 0.46 (0.35-0.57) | 0.66 (0.59-0.74) | 0.90 (0.80-1.04) |
| **Namibia** | 0.20 (0.12-0.27) | 0.36 (0.26-0.48) | 0.56 (0.50-0.62) | 0.87 (0.72-1.04) |
| **Guinea-Bissau** | 0.21 (0.14-0.27) | 0.36 (0.27-0.45) | 0.57 (0.51-0.63) | 0.83 (0.71-0.98) |
| **Botswana** | 0.19 (0.11-0.25) | 0.28 (0.20-0.37) | 0.50 (0.44-0.56) | 0.82 (0.66-0.99) |
| **Lesotho** | 0.00 (0.00-0.00) | 0.02 (0.00-0.07) | 0.29 (0.22-0.35) | 0.66 (0.52-0.80) |
| **Equatorial Guinea** | 0.15 (0.10-0.20) | 0.28 (0.20-0.39) | 0.43 (0.39-0.48) | 0.66 (0.55-0.78) |
| **Eswatini** | 0.15 (0.11-0.19) | 0.26 (0.20-0.31) | 0.32 (0.29-0.36) | 0.44 (0.39-0.50) |
| **Djibouti** | 0.05 (0.02-0.08) | 0.09 (0.06-0.14) | 0.17 (0.14-0.19) | 0.33 (0.25-0.40) |
| **Comoros** | 0.10 (0.07-0.13) | 0.14 (0.10-0.18) | 0.20 (0.18-0.22) | 0.28 (0.25-0.33) |
| **Mauritius** | 0.00 (0.00-0.00) | 0.00 (0.00-0.01) | 0.08 (0.06-0.10) | 0.20 (0.15-0.25) |
| **Cabo Verde** | 0.00 (0.00-0.00) | 0.01 (0.00-0.02) | 0.06 (0.04-0.07) | 0.14 (0.11-0.17) |
| **Sao Tome and Principe** | 0.01 (0.01-0.02) | 0.03 (0.02-0.04) | 0.05 (0.04-0.06) | 0.08 (0.07-0.10) |

*Minimum coverage required when only adults aged 20 years or above were eligible for vaccination*

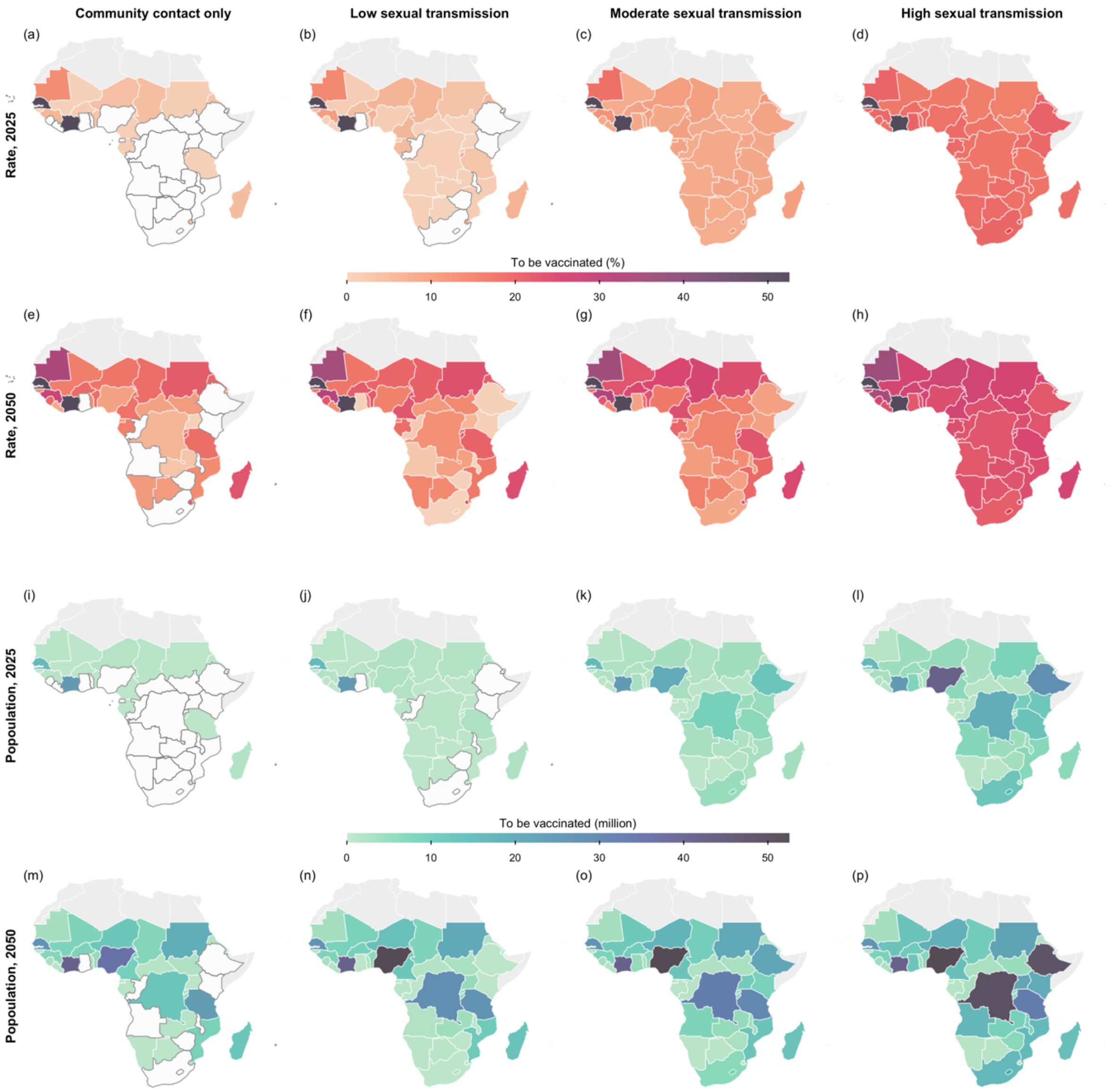

**Fig S7.** Projected minimal vaccine demand. Shown are the vaccine coverage rates (Row 1 and 2) and the number of individuals requiring vaccination (Row 3 and 4) to prevent secondary infections in each sub-Saharan African country modelled for 2025 (Row 1 and 3) and 2050 (Row 2 and 4), assuming vaccines accessible to adults aged at least 20 years. The four columns correspond to the four hypothetical scenarios with varying levels of sexual transmission. Countries which do not require vaccination are coloured in white, with borders outlined in dark grey. The base map layer (boundaries of African countries) is sourced from Natural Earth (https://www.naturalearthdata.com), available under the Public Domain license (https://www.naturalearthdata.com/about/terms-of-use/).

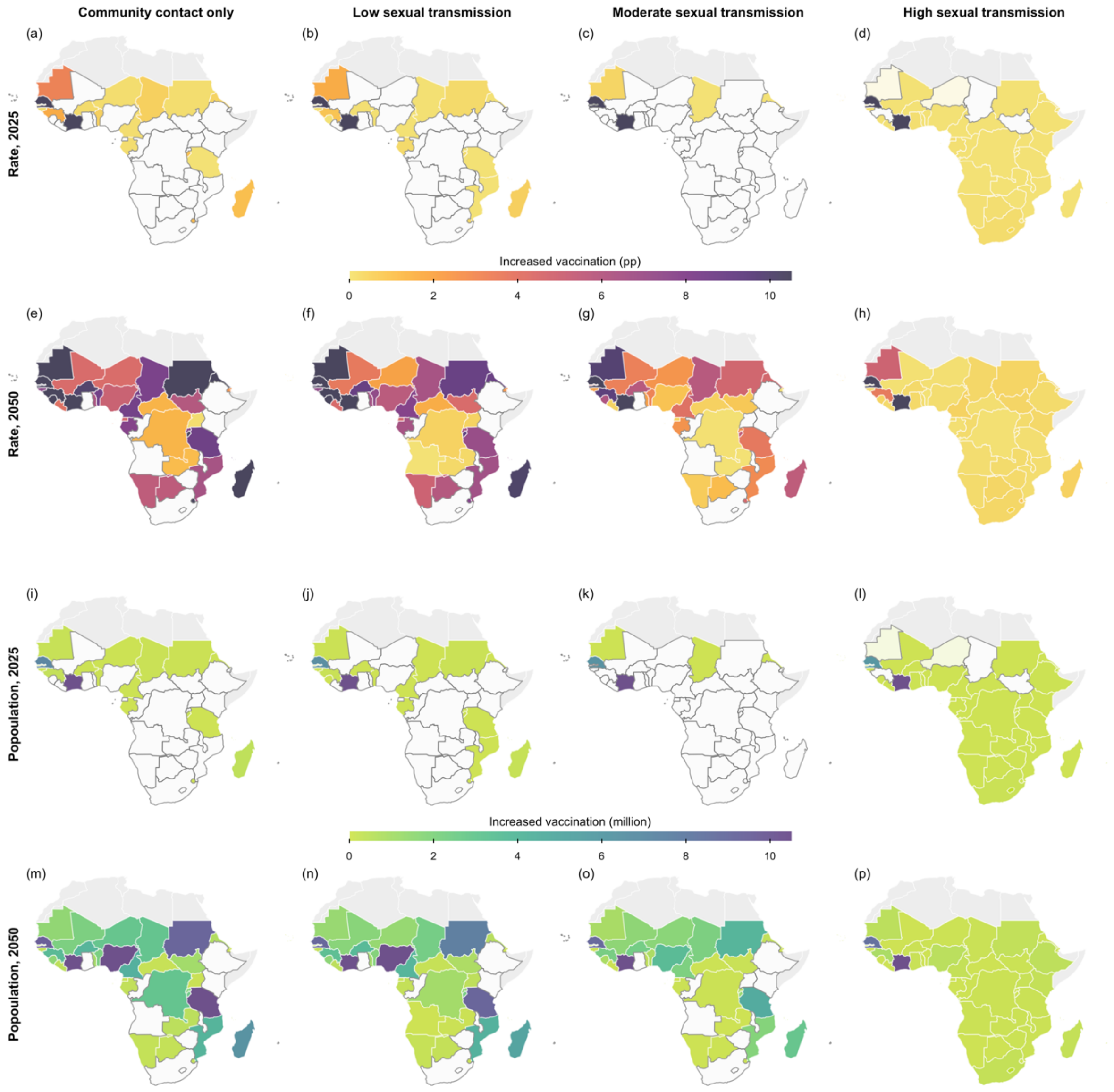

**Fig S8.** Projected increase in vaccine demand when vaccines were ineligible to individuals aged under 20 years of age. Shown are the differences in vaccine coverage rates (Row 1 and 2) and the number of individuals requiring vaccination (Row 3 and 4) to prevent secondary infections in each sub-Saharan African country modelled for 2025 (Row 1 and 3) and 2050 (Row 2 and 4). 'pp' stands for percentage point. Comparisons were made between the mass vaccination strategy and the strategy assuming vaccines accessible to adults aged at least 20 years. The four columns correspond to the four hypothetical scenarios with varying levels of sexual transmission. Countries unaffected by this age restriction in vaccine eligibility are coloured in white, with borders outlined in dark grey. The base map layer (boundaries of African countries) is sourced from Natural Earth (https://www.naturalearthdata.com), available under the Public Domain license (https://www.naturalearthdata.com/about/terms-of-use/).

*Minimum coverage required when prioritising high-sexual-activity females*

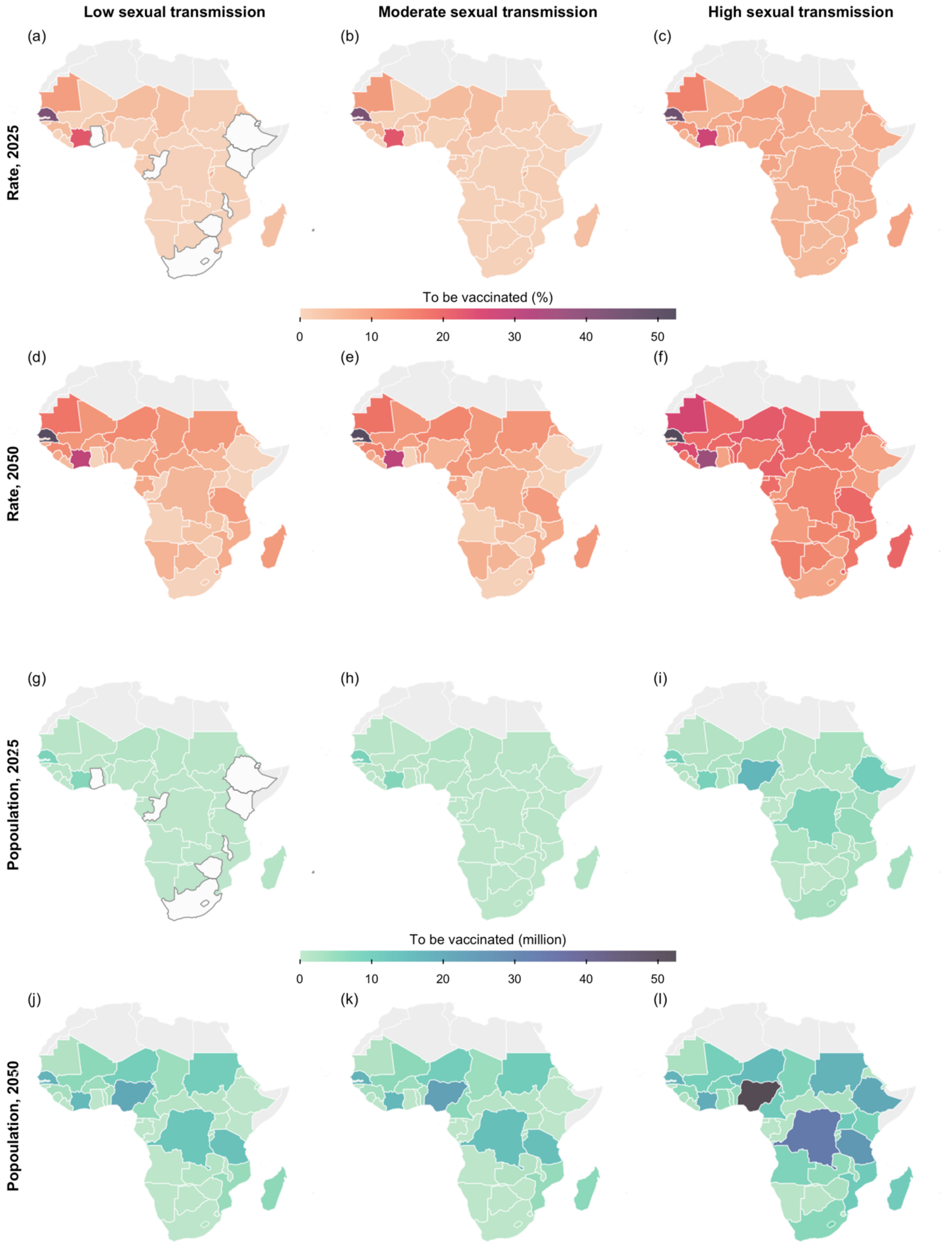

**Fig S9.** Projected minimal vaccine demand. Shown are the vaccine coverage rates (Row 1 and 2) and the number of individuals requiring vaccination (Row 3 and 4) to prevent secondary infections in each sub-Saharan African country modelled for 2025 (Row 1 and 3) and 2050 (Row 2 and 4), when prioritising high-sexual-activity females in conjunction with mass vaccination. The three columns correspond to the three hypothetical scenarios with low, middle, and high sexual transmission. Countries which do not require vaccination are coloured in white, with borders outlined in dark grey. The base map layer (boundaries of African countries) is sourced from Natural Earth (https://www.naturalearthdata.com), available under the Public Domain license (https://www.naturalearthdata.com/about/terms-of-use/).

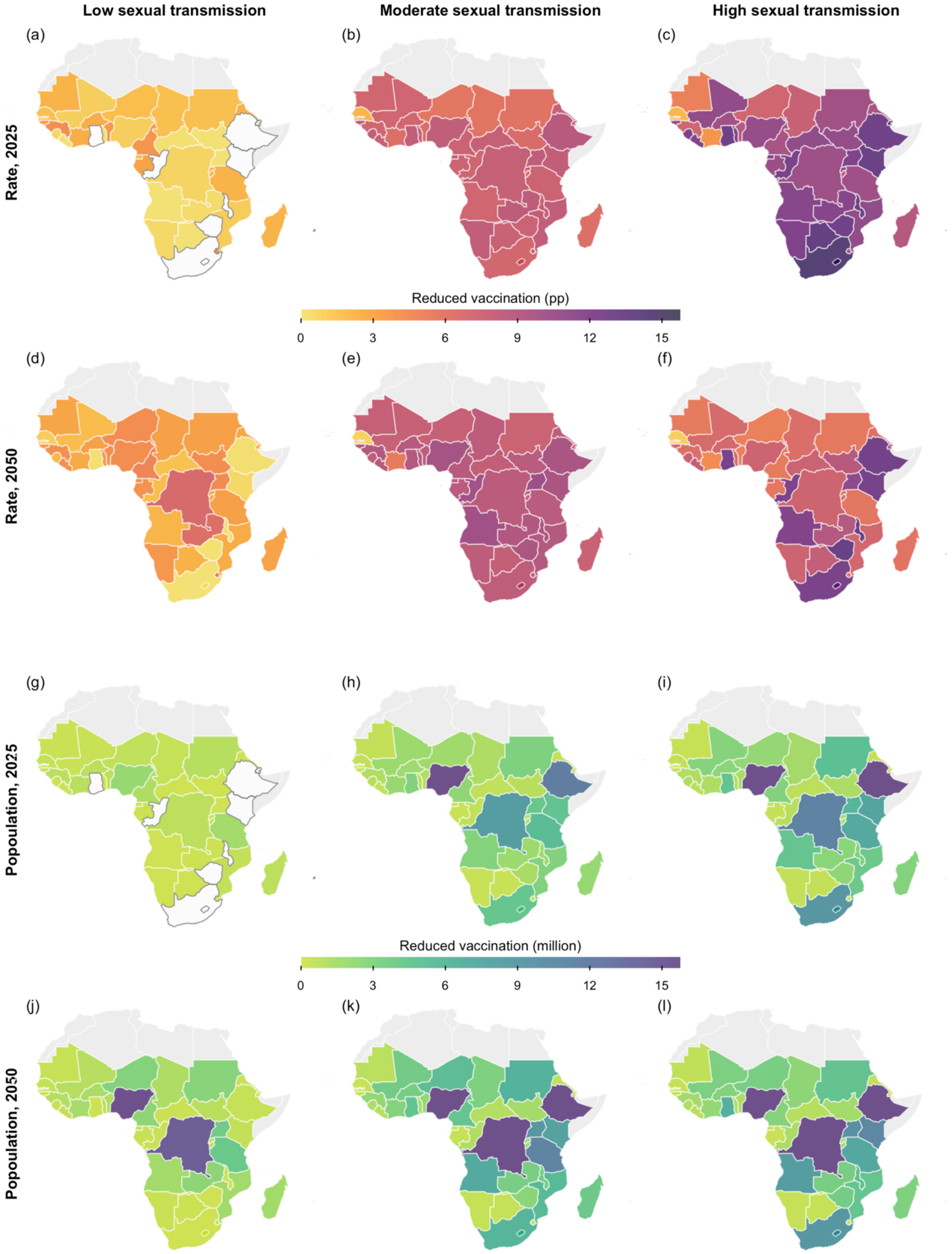

**Fig S10.** Projected reduction in vaccine demand through prioritisation of high-sexual-activity females. Shown are the differences in vaccine coverage rates (Row 1 and 2) and the number of individuals requiring vaccination

(Row 3 and 4) to prevent secondary infections in each sub-Saharan African country modelled for 2025 (Row 1 and 3) and 2050 (Row 2 and 4). 'pp' stands for percentage point. Comparisons were made between the mass vaccination strategy and the strategy prioritising high-sexual-activity females aged 15–49 years. The three columns correspond to the three hypothetical scenarios with low, middle, and high sexual transmission. Countries unaffected by this prioritisation are coloured in white, with borders outlined in dark grey. The base map layer (boundaries of African countries) is sourced from Natural Earth (https://www.naturalearthdata.com), available under the Public Domain license (https://www.naturalearthdata.com/about/terms-of-use/).

*Impacts of vaccination strategies assuming limited vaccine supply*

When targeting large cohorts or when the vaccine coverage was low, strategies employing the equal distribution approach, in which vaccines were allocated uniformly across selected age groups, were generally less effective compared to those sequentially allocating vaccines based on pre-specified orders. Nevertheless, their effectiveness might be comparable to sequential allocation in scenarios where the age groups were at similar risks (Fig S11).

In addition, a non-linear relationship was observed between vaccine coverage and reduction in $R_{eff}$ among sequential-allocation strategies, especially in settings involving higher levels of sexual transmission. This pattern was likely the result of disparities in the contribution of different age groups to overall transmission. The marginal reduction in $R_{eff}$ tended to be more pronounced when coverage in the general population rose from 10% to 20%, compared to a further 10 percentage points' increase. Nonetheless, a 20% coverage rate was projected to suffice for preventing epidemic growth in most countries by 2025. At this level, the most effective strategy (i.e., prioritising individuals aged 20–29 years and followed by those aged 30–39 and 40–49 years) was estimated to lower the expected $R_{eff}$ of 38 countries among the 47 modelled to below one across all four hypothetical scenarios, increasing to 45 countries with 30% coverage (Fig 4).

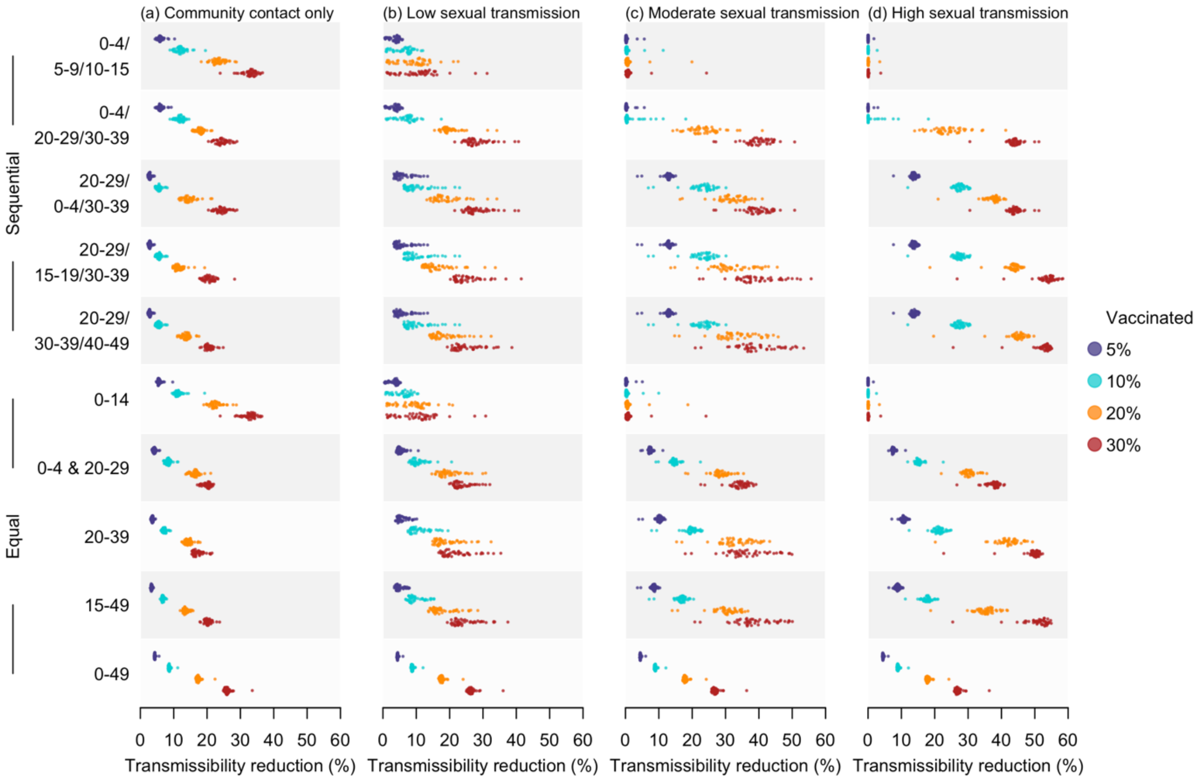

**Fig S11**. Estimated reduction in $R_{eff}$ in 2025 under diverse vaccination strategies, compared to the corresponding baseline scenarios with no vaccination. Doses were allocated using two allocation methods, including sequential (first five rows) and equal (last five rows). Four coverage rates, including 5%, 10%, 20%, and 30%, were assessed, with results summarised as shaded distributions in purple, blue, orange, and red, respectively. The four columns correspond to the four hypothetical scenarios with varying levels of sexual transmission.

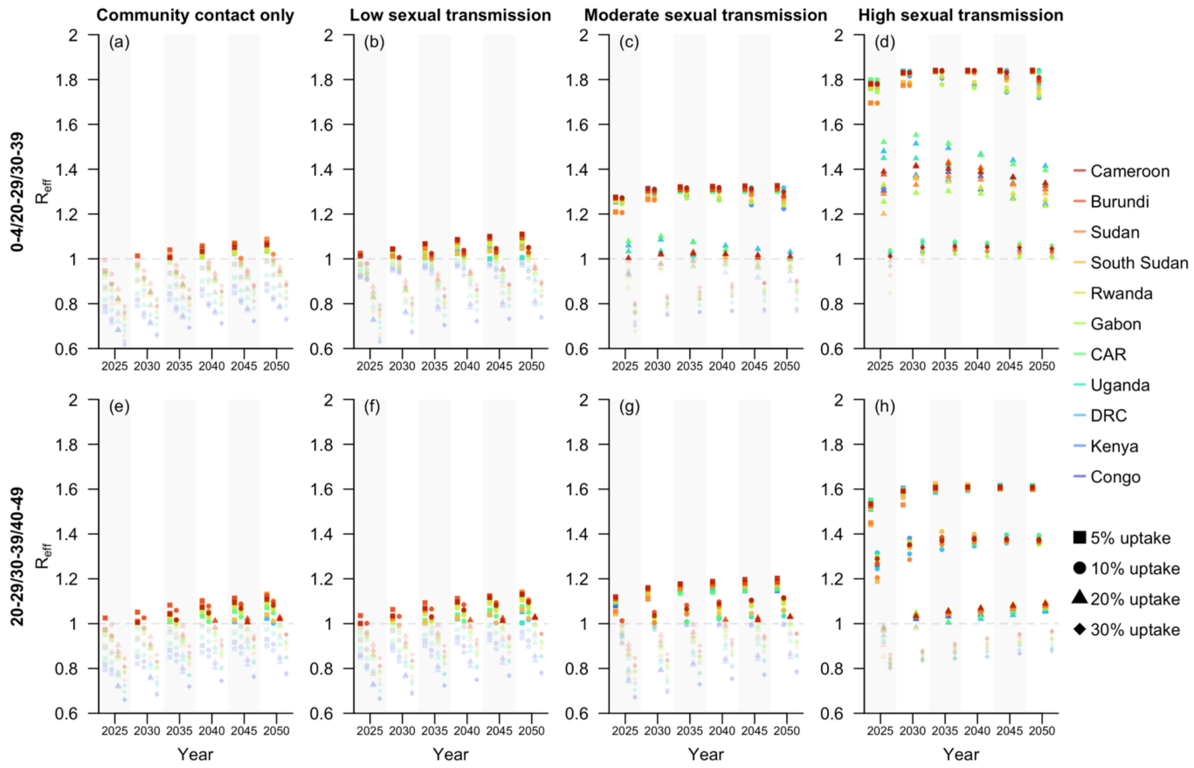

**Fig S12**. Projected $R_{eff}$ at five-year intervals from 2025 to 2050 under selected vaccination strategies for sub-Saharan African countries with documented local transmission of Clade I MPXV as of December 2024 [18]. These countries were ordered in descending estimated $R_{eff}$ for 2050, based on the scenario with high sexual transmission and the vaccination strategy prioritizing individuals aged 20–29, 30–39, and 40–49, sequentially (Row 2, Column 4). Four coverage rates, including 5%, 10%, 20%, and 30%, were assessed, with outcomes displayed as scattered dots across four columns within one subfigure. The rows of subfigures represent the two selected sequential vaccination strategies, while the columns correspond to the four hypothetical scenarios with varying levels of sexual transmission.

**Sensitivity analysis: model re-calibration using all components of the synthetic contact matrices**

To account for the potential presence of close contacts in non-household settings (e.g., schools), despite limited supporting evidence as of May 2025, we conducted a sensitivity analysis in which the model was re-calibrated using all components of the synthetic contact matrices [8]. As suggested in the preliminary study [1], this approach resulted in worse model fits compared to calibration using home contact components only (Fig S1, S13), with lower likelihood values and higher Akaike Information Criterion (AIC) scores.

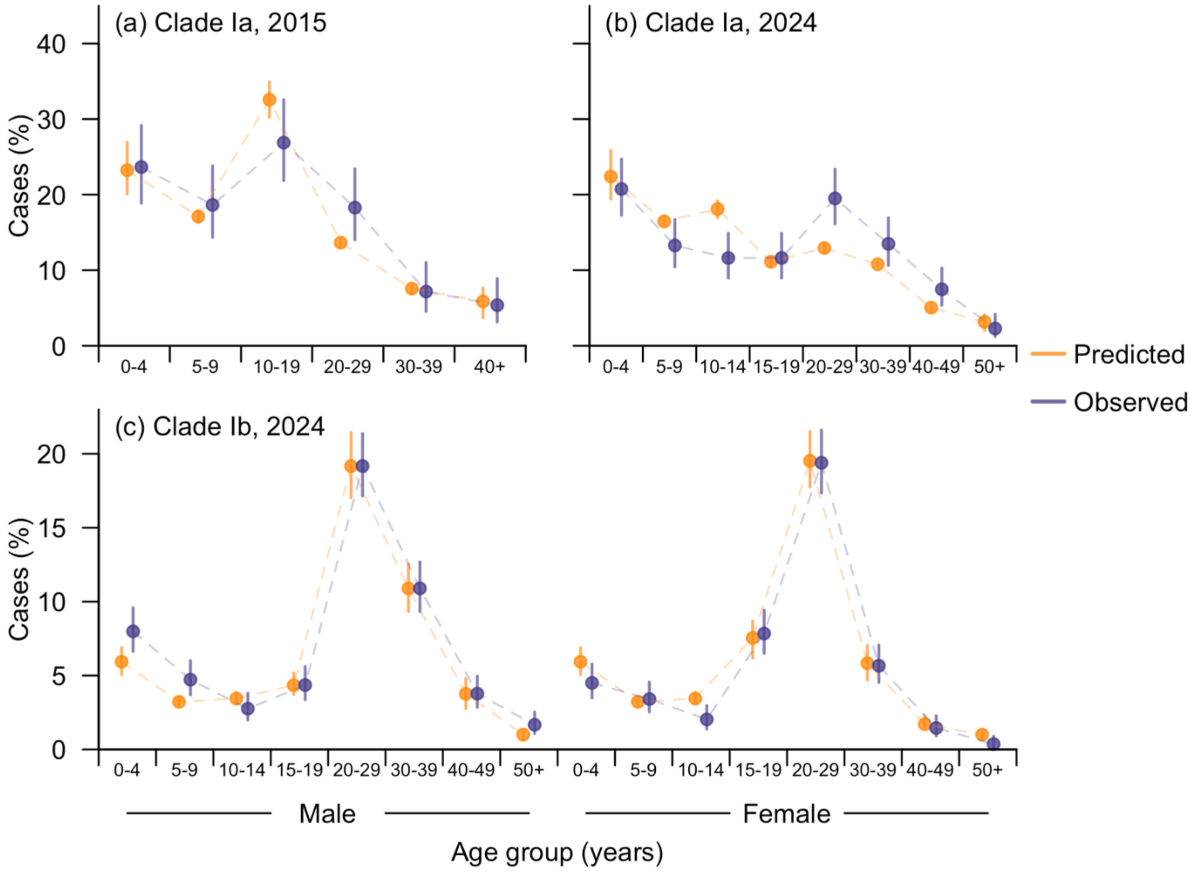

**Fig S13.** Fitted case demographics for the DRC, based on the model calibrated using all contact components of the synthetic contact matrices [8]. Subfigure (a) and (b) show the age distribution of case counts in endemic provinces in the DRC for the years 2015 and 2024, respectively, while subfigure (c) presents the age and sex distribution of cases counts in South Kivu, the DRC in 2024. Observed case demographics were obtained from Murayama et al [1].

The projected $R_{eff}$ values for the 47 sub-Saharan African countries were generally lower compared to those in the main analysis. For instance, in the DRC in 2024, the $R_{eff}$ for Clade Ia transmission was estimated at 0.85 (95% CI: 0.85–0.86), while that for Clade Ib transmission was 1.44 (95% CI: 1.36–1.51), with additional sexual transmission accounting for 43.6% (95% CI: 40.5%–46.6%) to overall transmission. This led to substantially lower projected mpox vaccine demands across all the 47 countries and under all four scenarios with varying levels of sexual transmission (Fig S14–S17). Nevertheless, the age prioritisation for vaccination in the case of limited vaccine supply was similar to the main analysis.

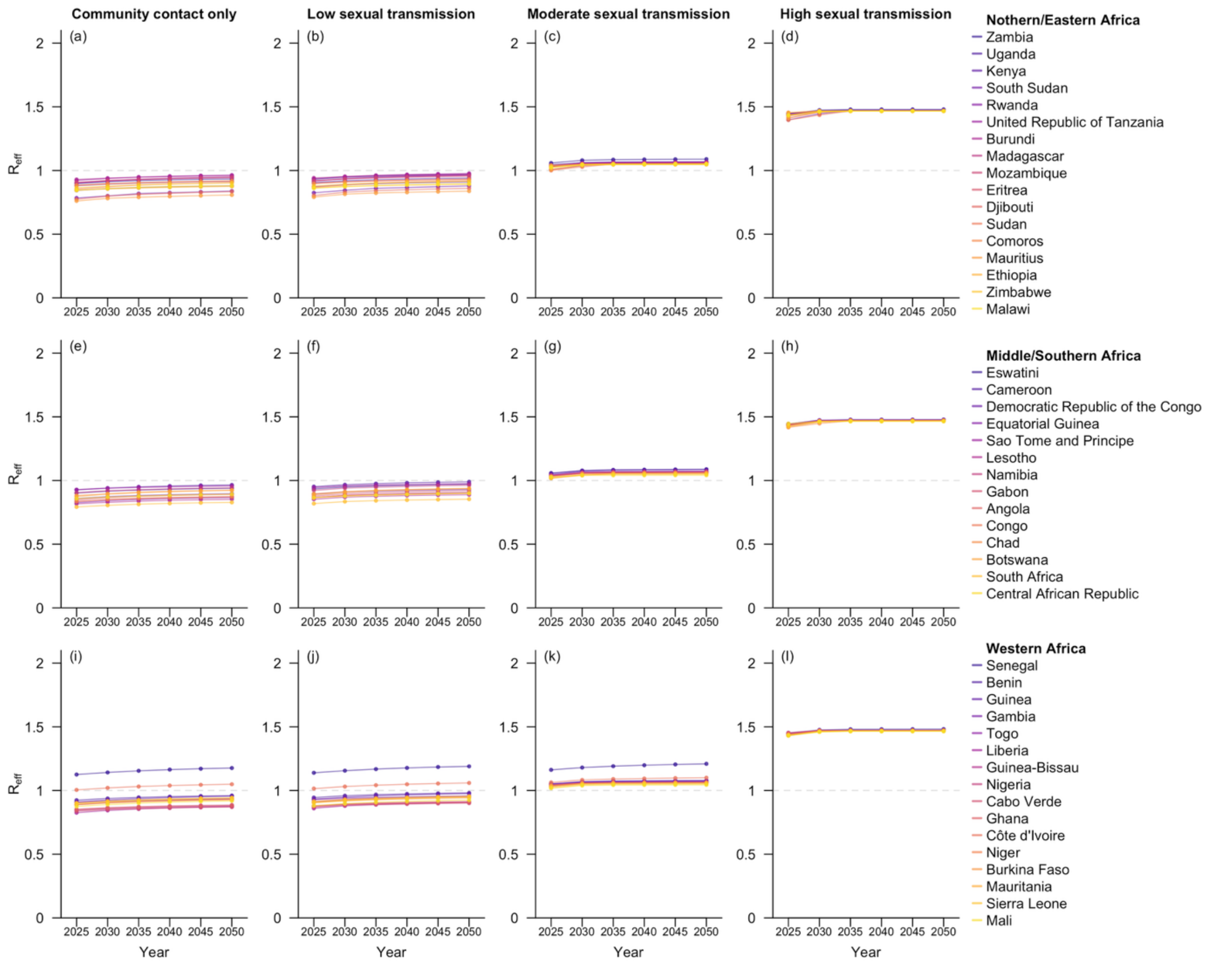

**Fig S14**. Projected $R_{eff}$ for the 47 sub-Saharan African countries at five-year intervals from 2025 to 2050, based on the model calibrated using all contact components of the synthetic contact matrices [8]. Countries within the same geospatial subregions are grouped in the same row and arranged in descending order by their estimated $R_{eff}$ in 2050. The four columns correspond to the four hypothetical scenarios with varying levels of sexual transmission.

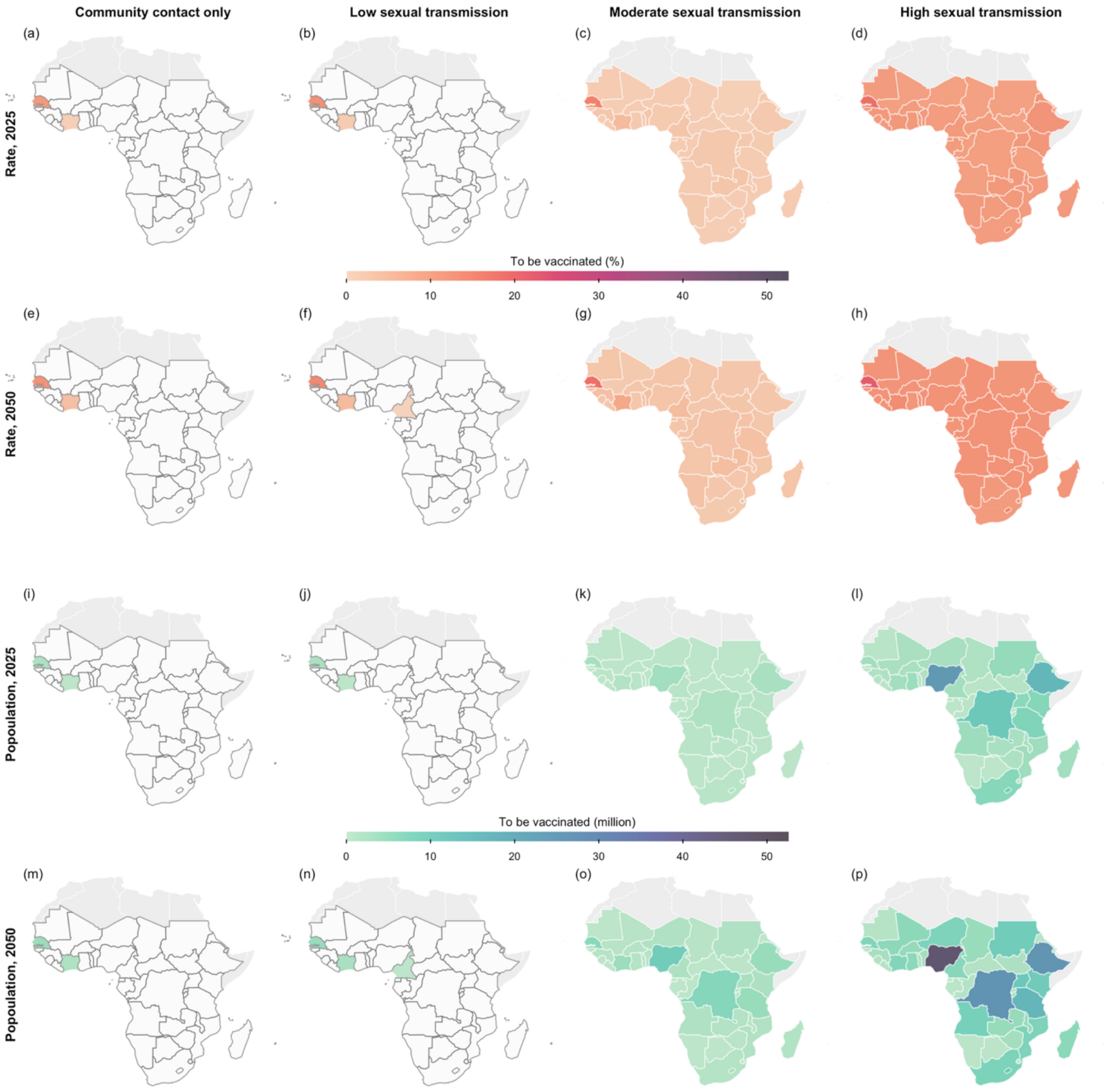

**Fig S15.** Projected minimal vaccine demand, based on the model calibrated using all contact components of the synthetic contact matrices [8]. Shown are the vaccine coverage rates (Row 1 and 2) and the number of individuals requiring vaccination (Row 3 and 4) to prevent secondary infections in each sub-Saharan African country modelled for 2025 (Row 1 and 3) and 2050 (Row 2 and 4), assuming mass vaccination. The four columns correspond to the four hypothetical scenarios with varying levels of sexual transmission. Countries which do not require vaccination are coloured in white, with borders outlined in dark grey. The base map layer (boundaries of African countries) is sourced from Natural Earth (https://www.naturalearthdata.com), available under the Public Domain license (https://www.naturalearthdata.com/about/terms-of-use/).

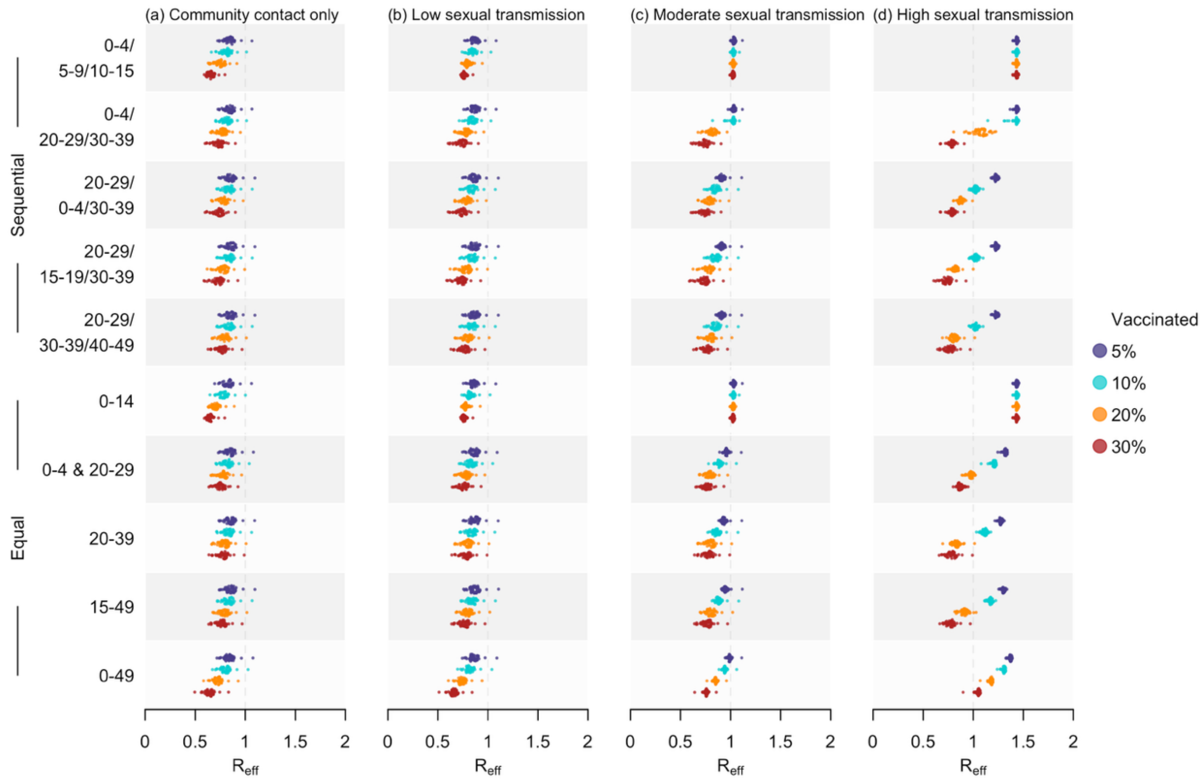

**Fig S16**. Estimated $R_{eff}$ in 2025 under diverse vaccination strategies, based on the model calibrated using all contact components of the synthetic contact matrices [8]. Doses were allocated using two allocation methods, including sequential (first five rows) and equal (last five rows). Four coverage rates, including 5%, 10%, 20%, and 30%, were assessed, with outcomes summarised as shaded distributions in purple, blue, orange, and red, respectively. The four columns correspond to the four hypothetical scenarios with varying levels of sexual transmission.

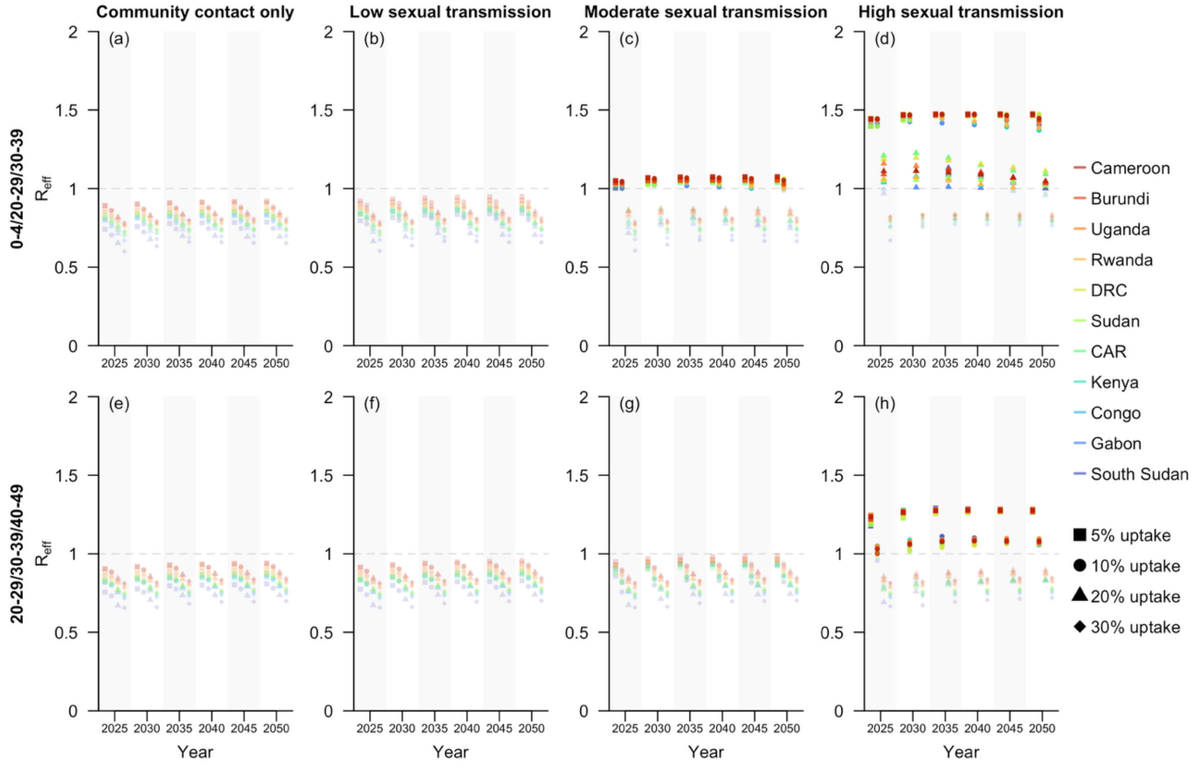

**Fig S17**. Projected $R_{eff}$ at five-year intervals from 2025 to 2050 under selected vaccination strategies for sub-Saharan African countries with documented local transmission of Clade I MPXV as of December 2024 [18], based on the model calibrated using all contact components of the synthetic contact matrices [8]. These countries were ordered in descending estimated $R_{eff}$ for 2050, based on the scenario with high sexual transmission and the vaccination strategy prioritizing individuals aged 20–29, 30–39, and 40–49, sequentially (Row 2, Column 4). Four coverage rates, including 5%, 10%, 20%, and 30%, were assessed, with outcomes displayed as scattered dots across four columns within one subfigure. The rows of subfigures represent the two selected sequential vaccination strategies, while the columns correspond to the four hypothetical scenarios with varying levels of sexual transmission.

**Sensitivity analysis: the potential waning immunity from smallpox vaccines**

In the main analysis, the smallpox vaccine effectiveness was assumed to remain constant over time. To examine the potential impact of waning immunity, we conducted a sensitivity analysis by modelling the time-varying effectiveness as a non-increasing function of year $t$:

$$e_{s,t} = e_{s,2015}(1 - r_e)^{t-2015},$$

where $r_e (\geq 0)$ is the annual reduction in vaccine effectiveness level. Due to the absence of individual-level vaccination timing data, we did not stratify vaccine effectiveness by age. We re-calibrated the model and estimated both $e_{s,2015}$ and $r_e$ using the Clade Ia outbreak data from the DRC in 2015 and 2024, while all other parameters were derived as the main analysis. The estimated vaccine effectiveness in 2015, $e_{s,2015}$, was 90.9% (95% CI: 84.4%–96.9%), while $r_e$ was estimated at 1.0% (95% CI: 0–2.4%), indicating an insignificant decline in effectiveness over time, which also aligns with literature [4]. Compared to the main analysis which assumed constant smallpox vaccine effectiveness, the projection $R_{eff}$s under this scenario were slightly higher, leading to increased demands for mpox vaccines, (Fig S18–S21).

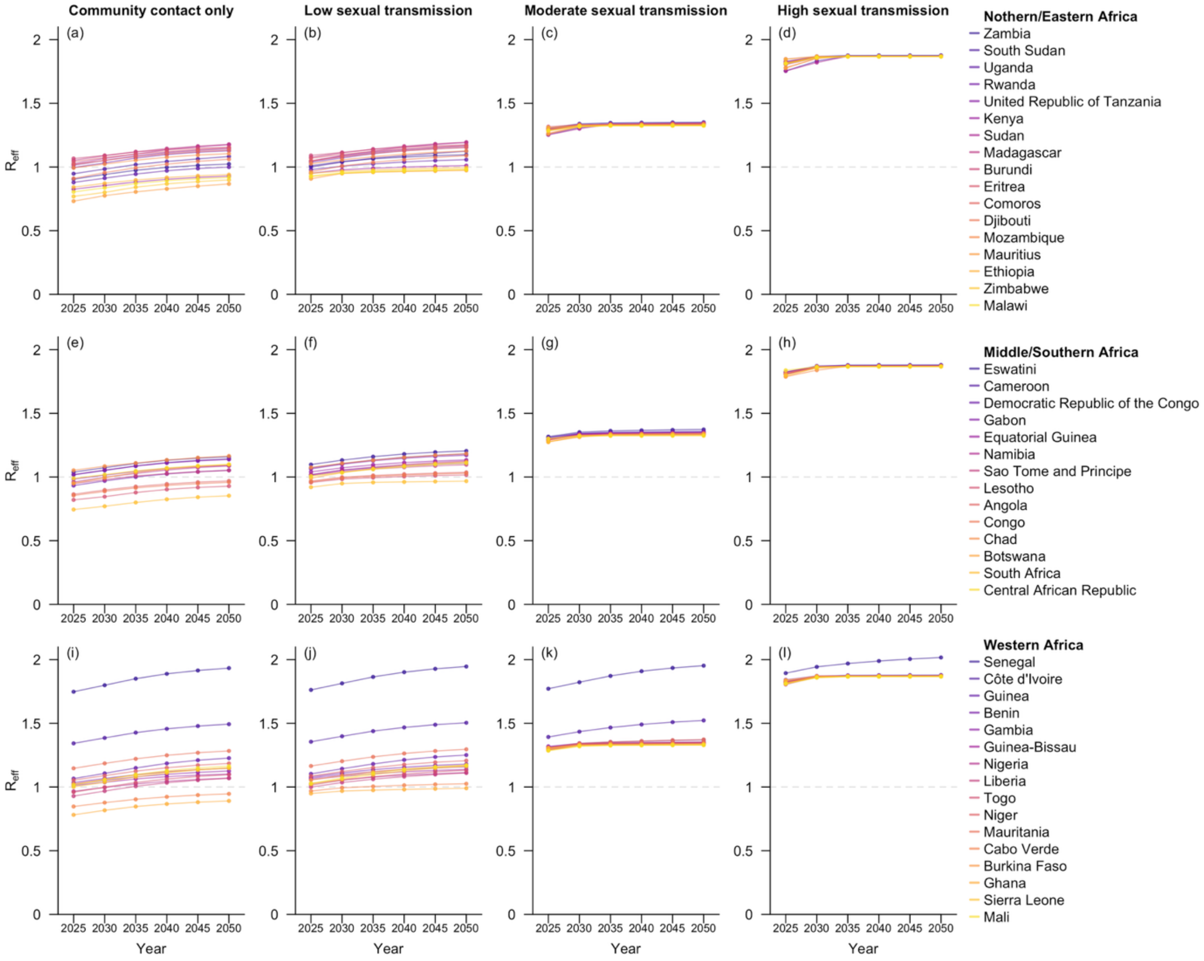

**Fig S18**. Projected $R_{eff}$ for the 47 sub-Saharan African countries at five-year intervals from 2025 to 2050, adjusted for potential waning immunity from historical smallpox vaccination. Countries within the same geospatial subregions are grouped in the same row and arranged in descending order by their estimated $R_{eff}$ in 2050. The four columns correspond to the four hypothetical scenarios with varying levels of sexual transmission.

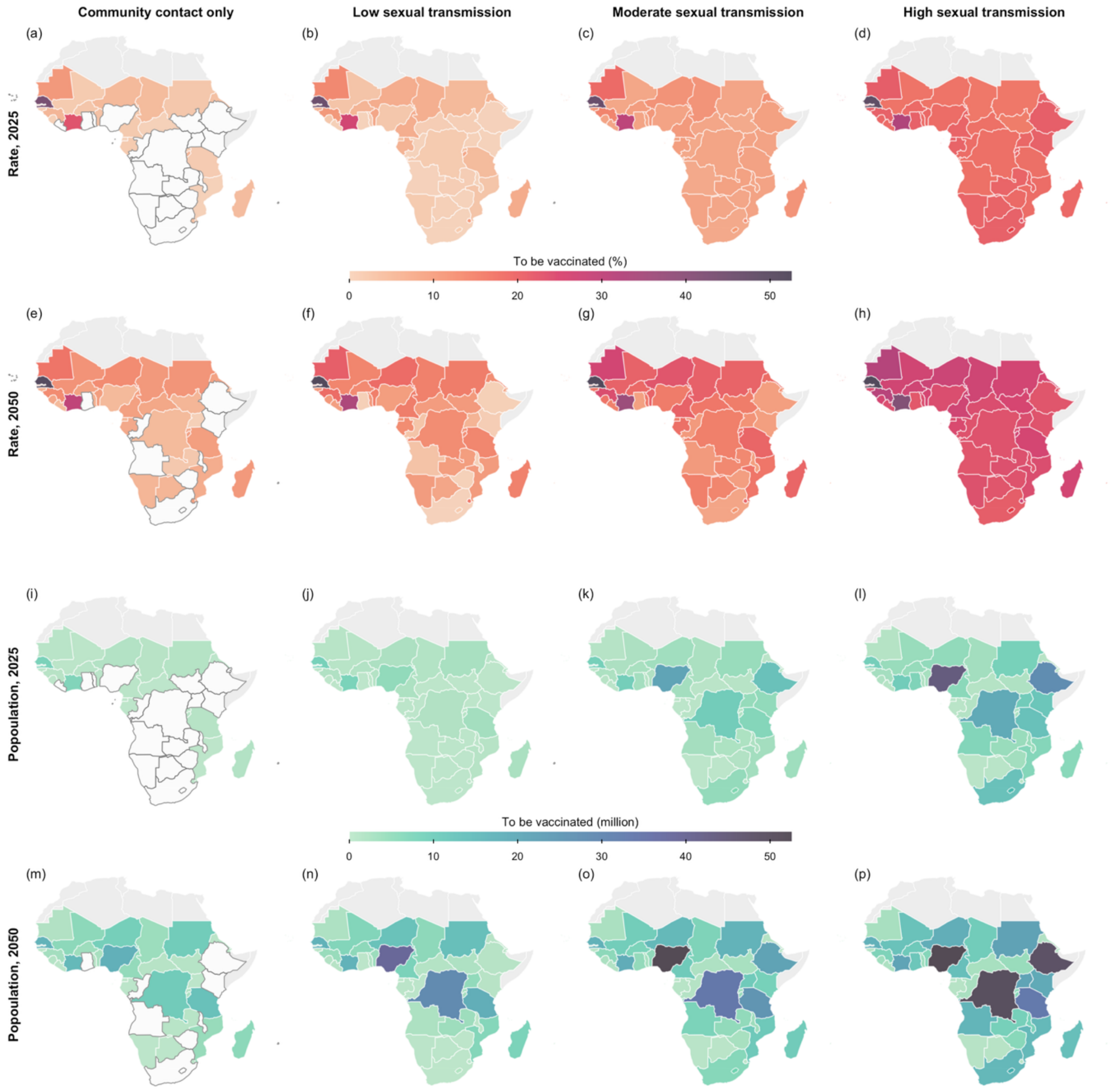

**Fig S19.** Projected minimal vaccine demand, adjusted for potential waning immunity from historical smallpox vaccination. Shown are the vaccine coverage rates (Row 1 and 2) and the number of individuals requiring vaccination (Row 3 and 4) to prevent secondary infections in each sub-Saharan African country modelled for 2025 (Row 1 and 3) and 2050 (Row 2 and 4), assuming mass vaccination. The four columns correspond to the four hypothetical scenarios with varying levels of sexual transmission. Countries which do not require vaccination are coloured in white, with borders outlined in dark grey. The base map layer (boundaries of African countries) is sourced from Natural Earth (https://www.naturalearthdata.com), available under the Public Domain license (https://www.naturalearthdata.com/about/terms-of-use/).

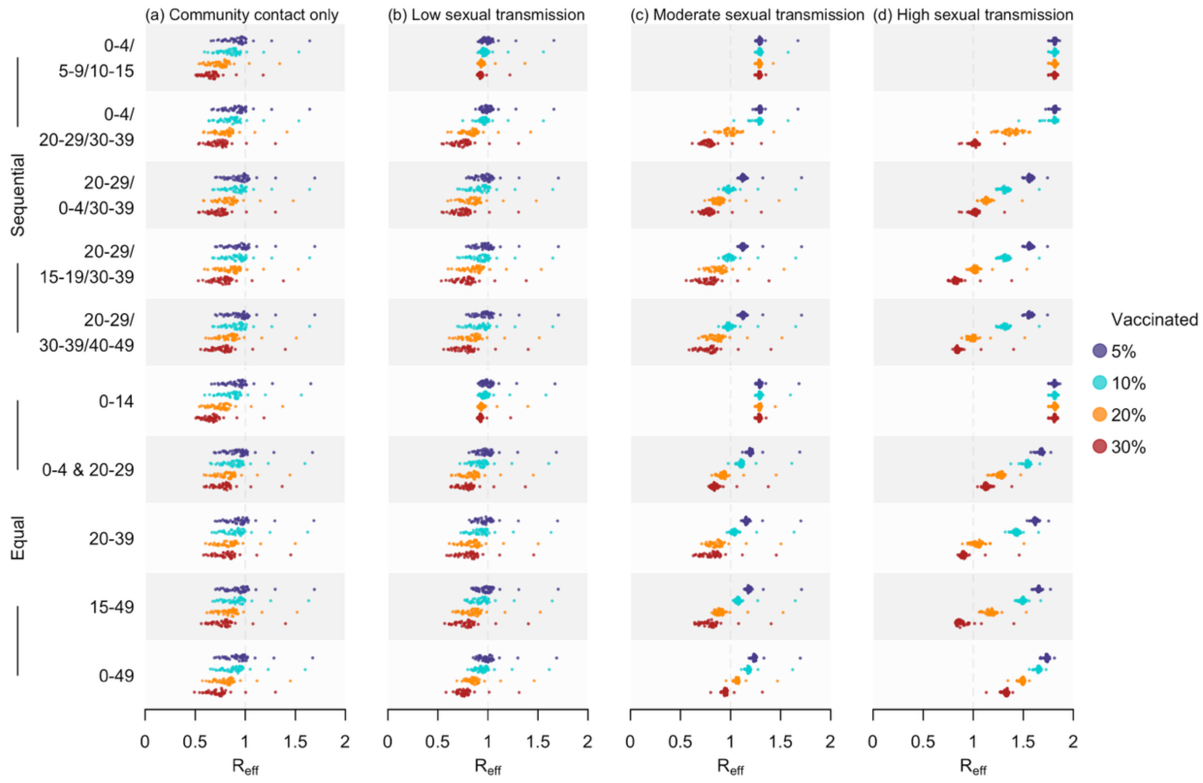

**Fig S20**. Estimated $R_{eff}$ in 2025 under diverse vaccination strategies, adjusted for potential waning immunity from historical smallpox vaccination. Doses were allocated using two allocation methods, including sequential (first five rows) and equal (last five rows). Four coverage rates, including 5%, 10%, 20%, and 30%, were assessed, with outcomes summarised as shaded distributions in purple, blue, orange, and red, respectively. The four columns correspond to the four hypothetical scenarios with varying levels of sexual transmission.

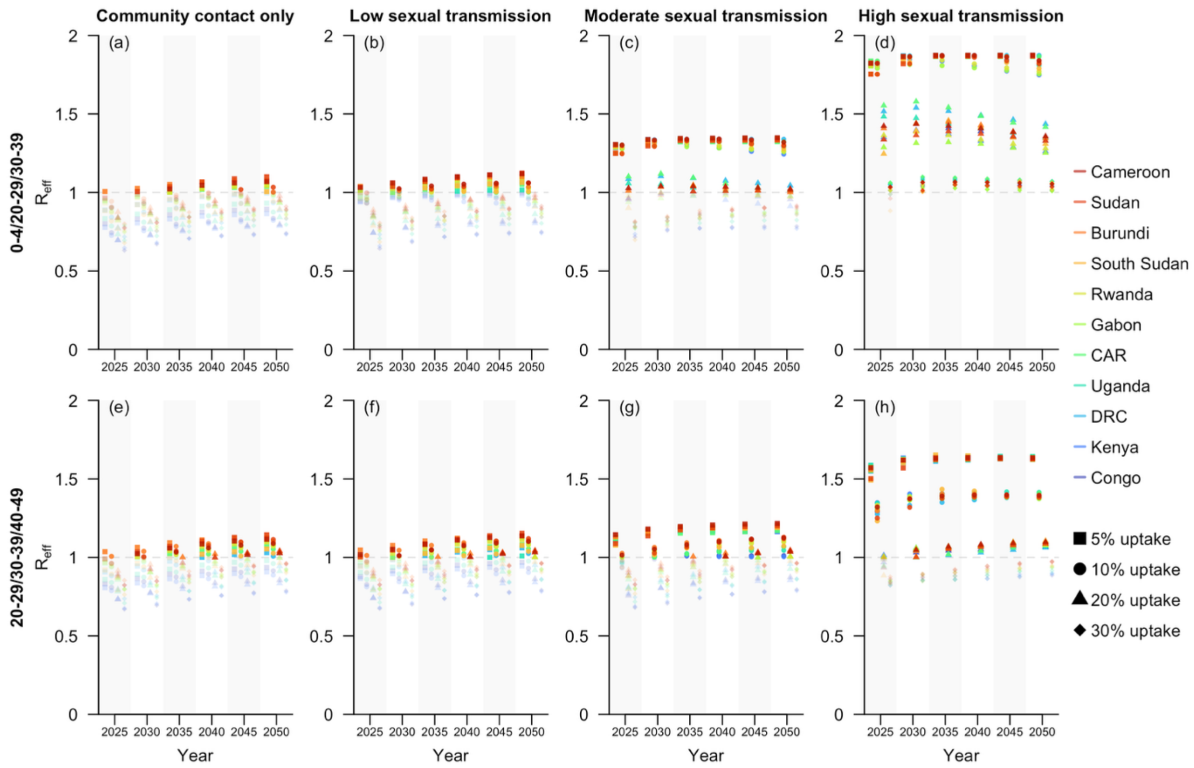

**Fig S21**. Projected $R_{eff}$ at five-year intervals from 2025 to 2050 under selected vaccination strategies for sub-Saharan African countries with documented local transmission of Clade I MPXV as of December 2024 [18], adjusted for potential waning immunity from historical smallpox vaccination. These countries were ordered in descending estimated $R_{eff}$ for 2050, based on the scenario with high sexual transmission and the vaccination strategy prioritizing individuals aged 20–29, 30–39, and 40–49, sequentially (Row 2, Column 4). Four coverage rates, including 5%, 10%, 20%, and 30%, were assessed, with outcomes displayed as scattered dots across four columns within one subfigure. The rows of subfigures represent the two selected sequential vaccination strategies, while the columns correspond to the four hypothetical scenarios with varying levels of sexual transmission.

**Sensitivity analysis: higher smallpox vaccine coverage in sub-Saharan African countries**

In the main analysis, the age-specific smallpox vaccine coverage was based on the mean estimates reported by Taube et al [2]. To evaluate the impact of higher-than-expected coverage on model projections, we alternatively utilized the upper bound of the 99% CI as the estimated vaccine coverage. The model was then re-calibrated, and the smallpox vaccine effectiveness, $e_s$, was estimated at 84.8% (95% CI: 78.5%–90.7%), lower than the estimate obtained in the main analysis. This change also introduced variation to the projected $R_{eff}$ values as well as vaccine demands in the sub-Saharan African countries (Fig S22–S25).

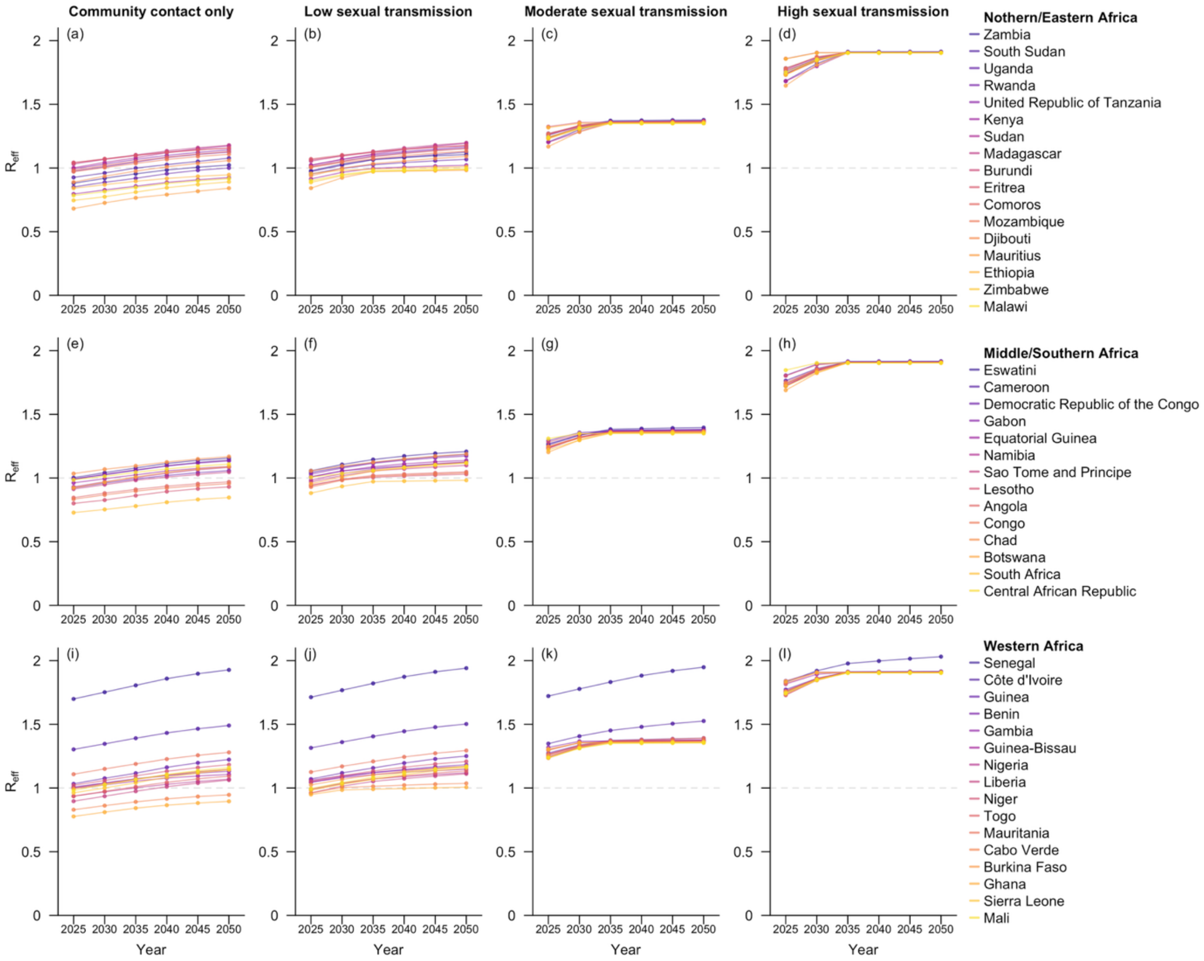

**Fig S22**. Projected $R_{eff}$ for the 47 sub-Saharan African countries at five-year intervals from 2025 to 2050, assuming higher-than-expected smallpox vaccine coverage. Countries within the same geospatial subregions are grouped in the same row and arranged in descending order by their estimated $R_{eff}$ in 2050. The four columns correspond to the four hypothetical scenarios with varying levels of sexual transmission.

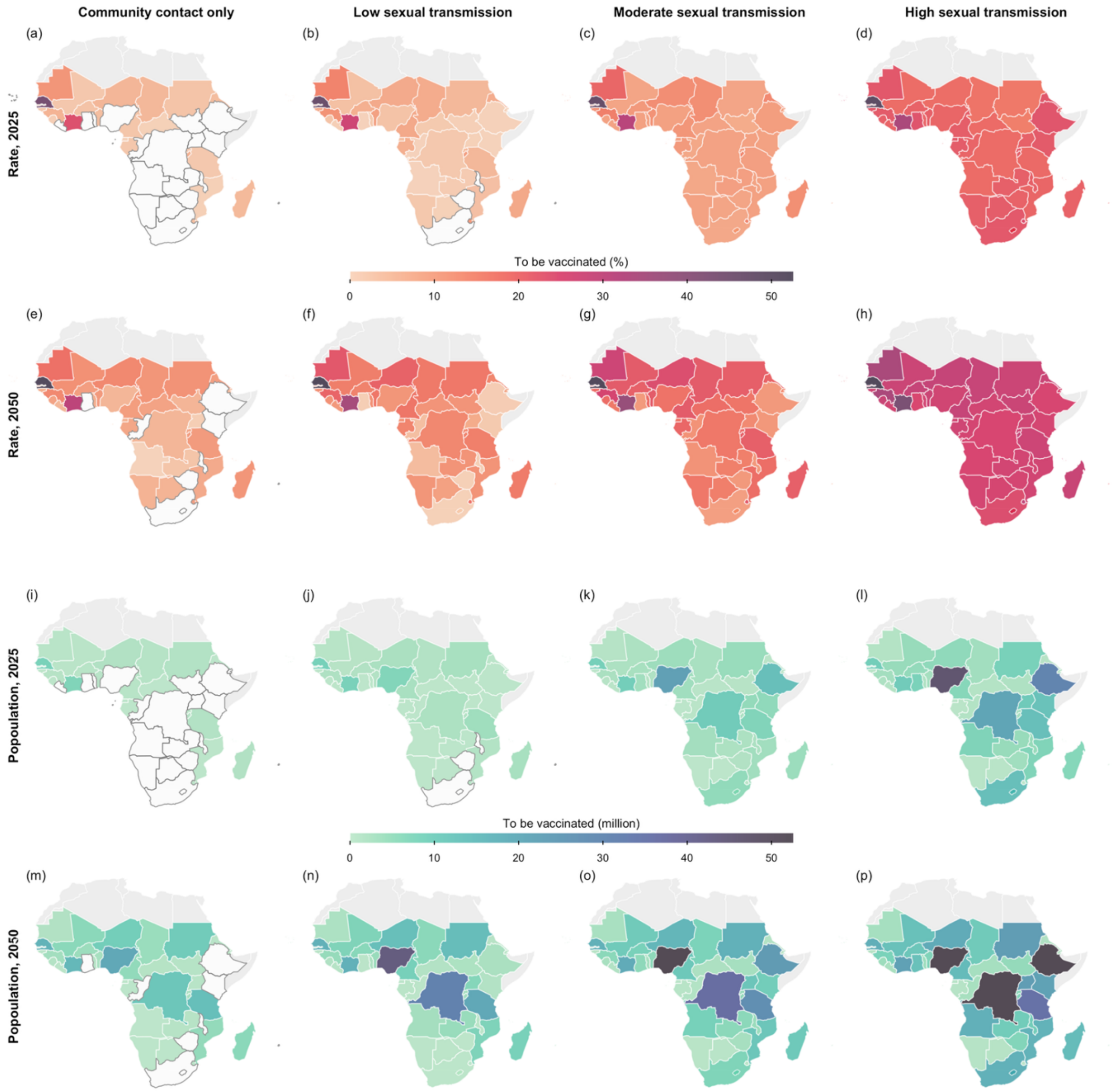

**Fig S23.** Projected minimal vaccine demand, assuming higher-than-expected smallpox vaccine coverage. Shown are the vaccine coverage rates (Row 1 and 2) and the number of individuals requiring vaccination (Row 3 and 4) to prevent secondary infections in each sub-Saharan African country modelled for 2025 (Row 1 and 3) and 2050 (Row 2 and 4), assuming mass vaccination. The four columns correspond to the four hypothetical scenarios with varying levels of sexual transmission. Countries which do not require vaccination are coloured in white, with borders outlined in dark grey. The base map layer (boundaries of African countries) is sourced from Natural Earth (https://www.naturalearthdata.com), available under the Public Domain license (https://www.naturalearthdata.com/about/terms-of-use/).

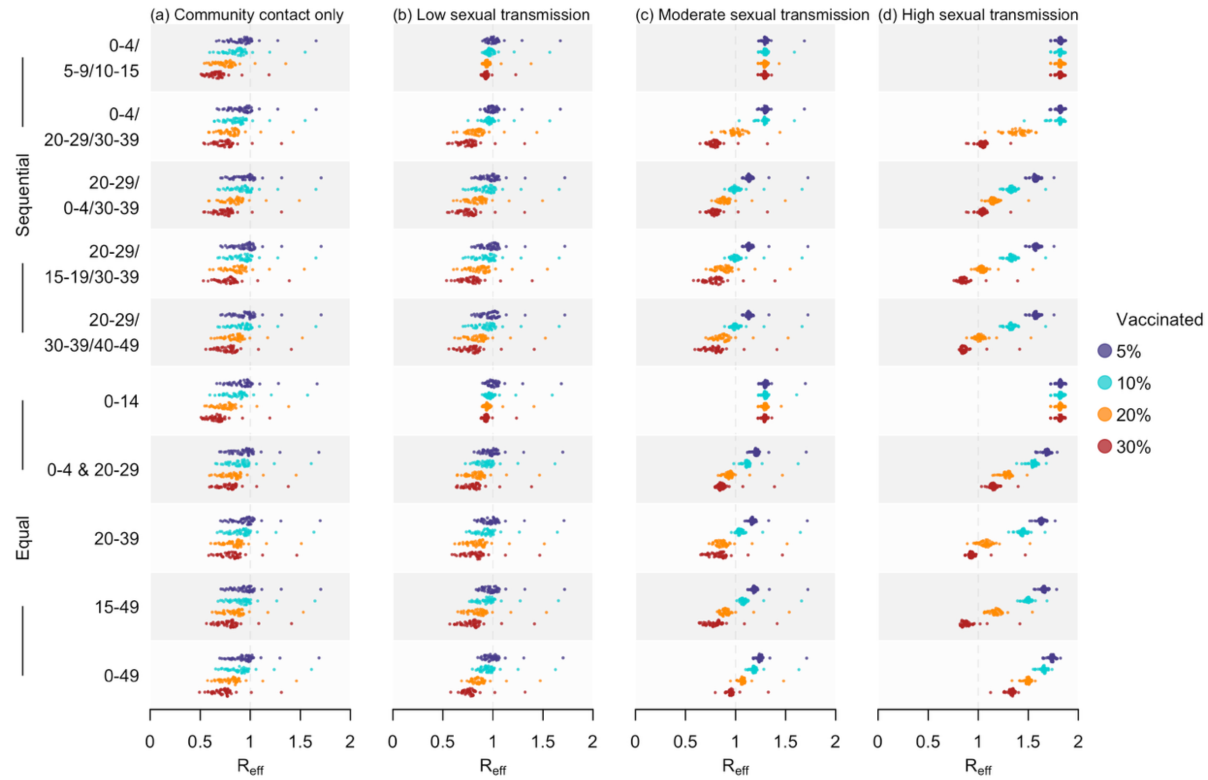

**Fig S24**. Estimated $R_{eff}$ in 2025 under diverse vaccination strategies, assuming higher-than-expected smallpox vaccine coverage. Doses were allocated using two allocation methods, including sequential (first five rows) and equal (last five rows). Four coverage rates, including 5%, 10%, 20%, and 30%, were assessed, with outcomes summarised as shaded distributions in purple, blue, orange, and red, respectively. The four columns correspond to the four hypothetical scenarios with varying levels of sexual transmission.

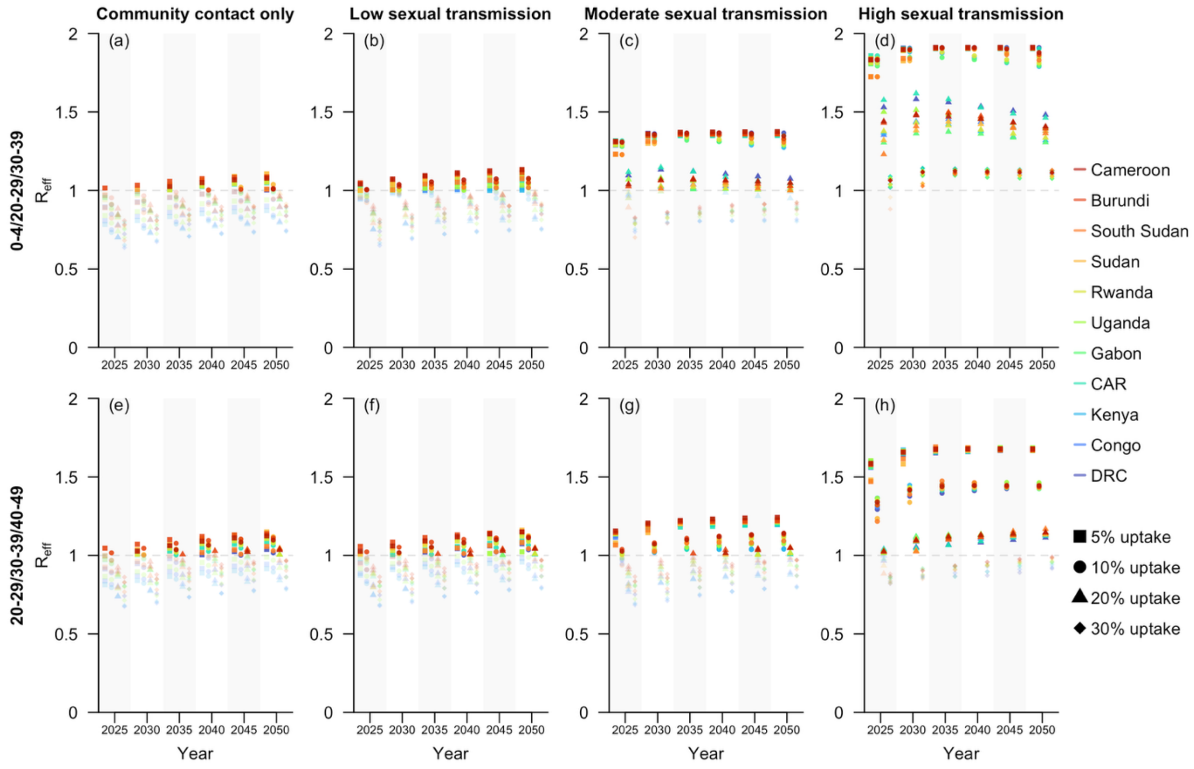

**Fig S25**. Projected $R_{eff}$ at five-year intervals from 2025 to 2050 under selected vaccination strategies for sub-Saharan African countries with documented local transmission of Clade I MPXV as of December 2024 [18], assuming higher-than-expected smallpox vaccine coverage. These countries were ordered in descending estimated $R_{eff}$ for 2050, based on the scenario with high sexual transmission and the vaccination strategy prioritizing individuals aged 20–29, 30–39, and 40–49, sequentially (Row 2, Column 4). Four coverage rates, including 5%, 10%, 20%, and 30%, were assessed, with outcomes displayed as scattered dots across four columns within one subfigure. The rows of subfigures represent the two selected sequential vaccination strategies, while the columns correspond to the four hypothetical scenarios with varying levels of sexual transmission.

**Sensitivity analysis: scaling the next generation matrix with uncertainty adjustments**

In the main analysis, we scaled the next generation matrix so that the estimated $R_{eff} = 0.82$ for Clade Ia MPXV in 2015. For this sensitivity analysis, we introduced uncertainty by allowing $R_{eff} \sim N(0.82, 0.015^2)$, corresponding to a 95% CI of approximately 0.79–0.85 [19]. We then re-ran the projections of $R_{eff}$s from 2025 to 2050 and re-estimated the mpox vaccine demand across the 47 sub-Saharan African countries. The results showed only minor differences from the main analysis (Fig S26–S29).

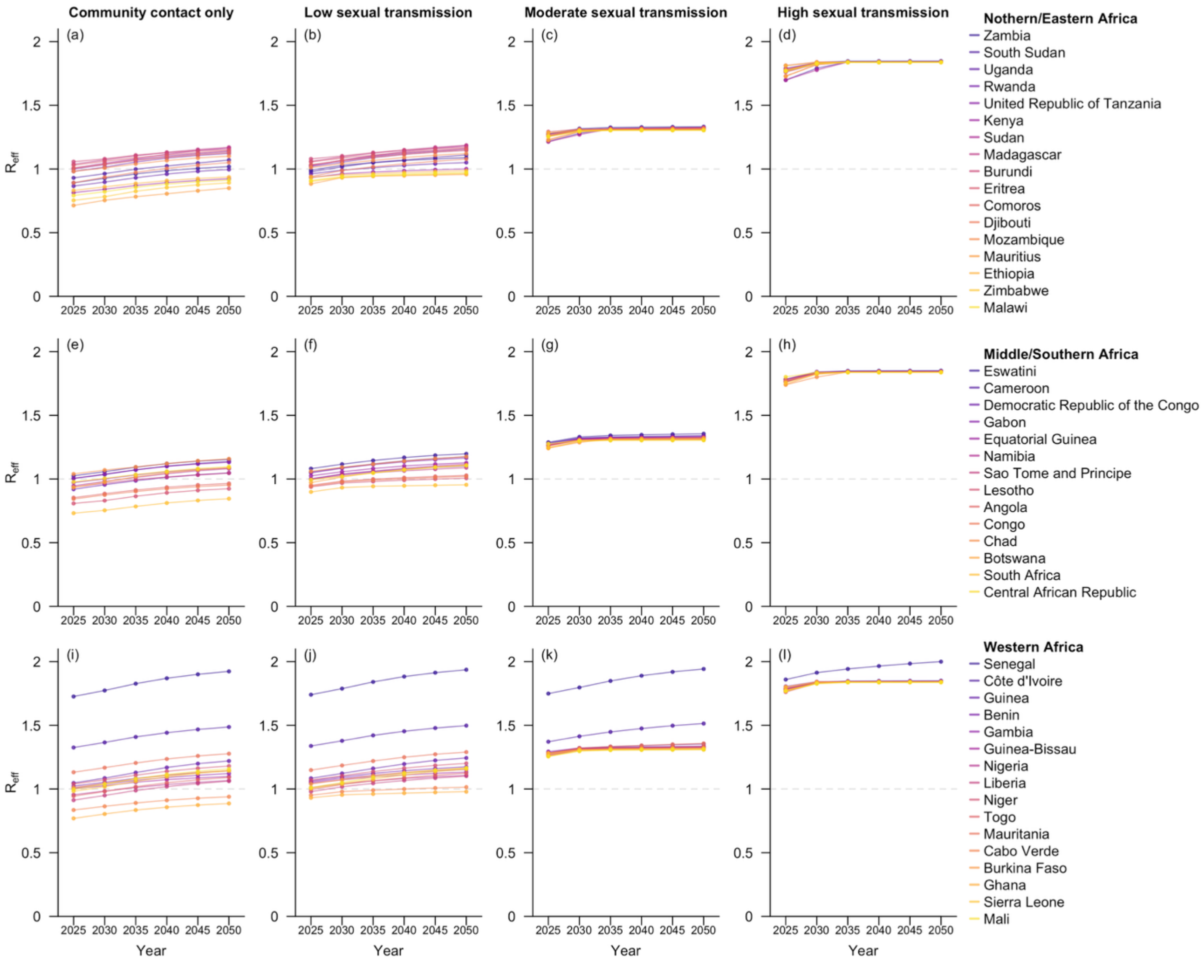

**Fig S26**. Projected $R_{eff}$ for the 47 sub-Saharan African countries at five-year intervals from 2025 to 2050, with uncertainty incorporated in the scaling of the next generation matrix. Countries within the same geospatial subregions are grouped in the same row and arranged in descending order by their estimated $R_{eff}$ in 2050. The four columns correspond to the four hypothetical scenarios with varying levels of sexual transmission.

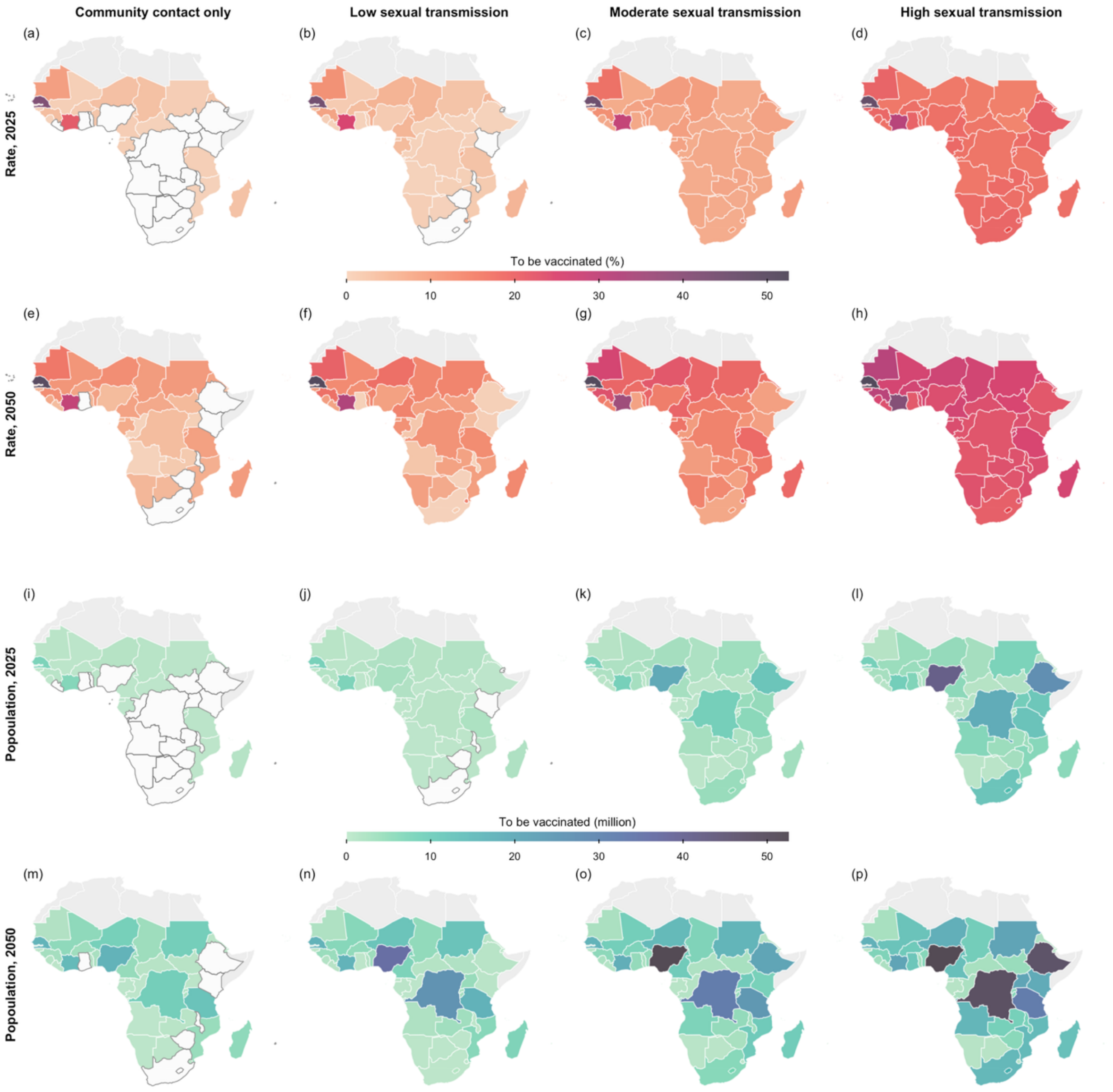

**Fig S27.** Projected minimal vaccine demand, with uncertainty incorporated in the scaling of the next generation matrix. Shown are the vaccine coverage rates (Row 1 and 2) and the number of individuals requiring vaccination (Row 3 and 4) to prevent secondary infections in each sub-Saharan African country modelled for 2025 (Row 1 and 3) and 2050 (Row 2 and 4), assuming mass vaccination. The four columns correspond to the four hypothetical scenarios with varying levels of sexual transmission. Countries which do not require vaccination are coloured in white, with borders outlined in dark grey. The base map layer (boundaries of African countries) is sourced from Natural Earth (https://www.naturalearthdata.com), available under the Public Domain license (https://www.naturalearthdata.com/about/terms-of-use/).

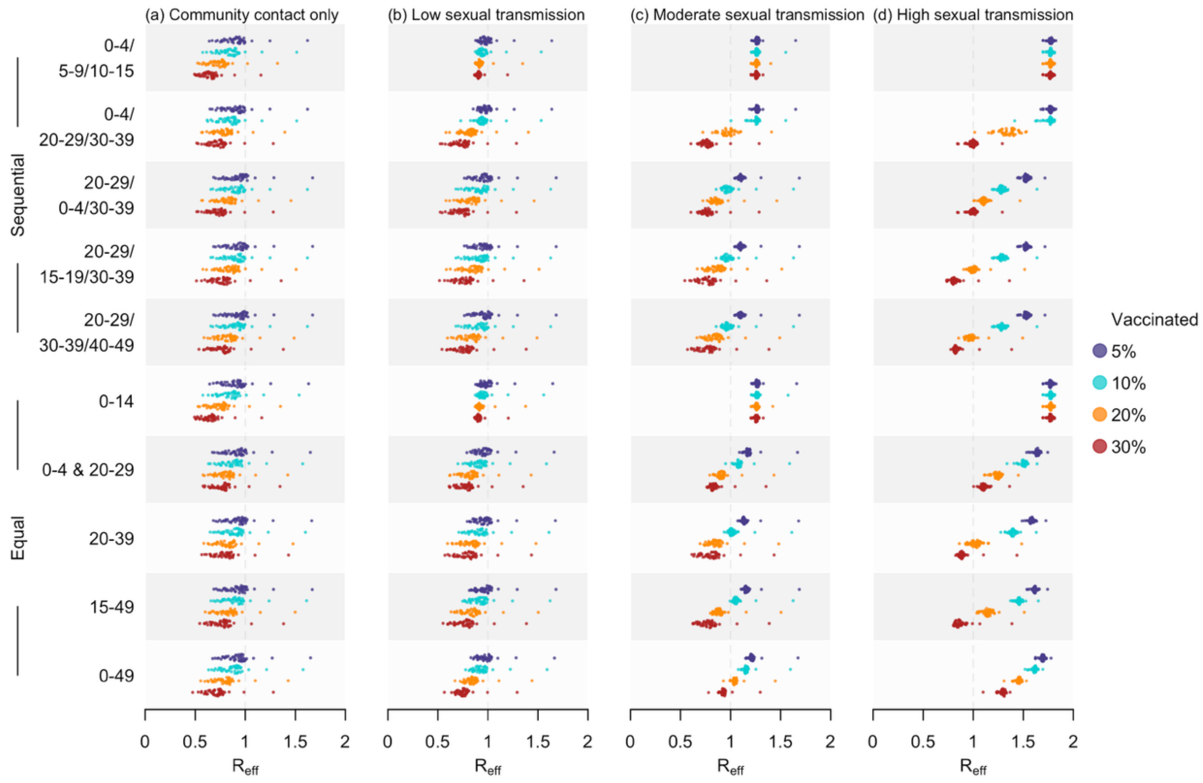

**Fig S28**. Estimated $R_{eff}$ in 2025 under diverse vaccination strategies, with uncertainty incorporated in the scaling of the next generation matrix. Doses were allocated using two allocation methods, including sequential (first five rows) and equal (last five rows). Four coverage rates, including 5%, 10%, 20%, and 30%, were assessed, with outcomes summarised as shaded distributions in purple, blue, orange, and red, respectively. The four columns correspond to the four hypothetical scenarios with varying levels of sexual transmission.

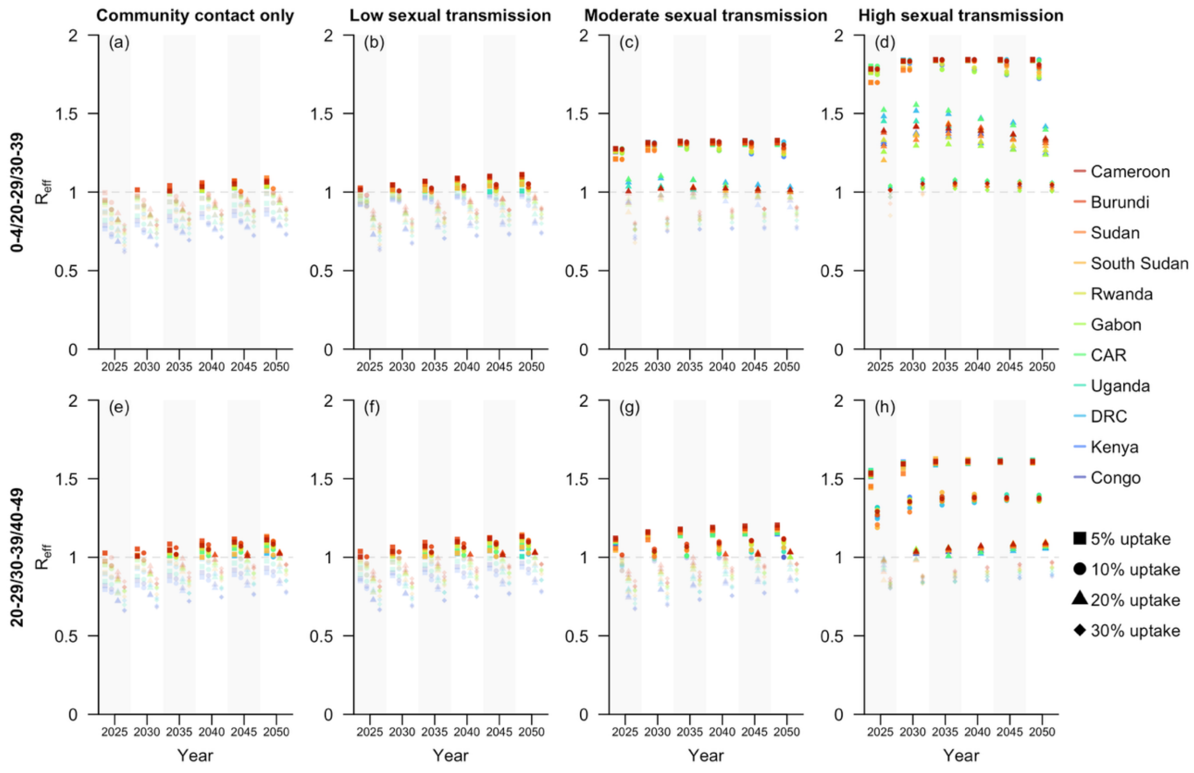

**Fig S29**. Projected $R_{eff}$ at five-year intervals from 2025 to 2050 under selected vaccination strategies for sub-Saharan African countries with documented local transmission of Clade I MPXV as of December 2024 [18], with uncertainty incorporated in the scaling of the next generation matrix. These countries were ordered in descending estimated $R_{eff}$ for 2050, based on the scenario with high sexual transmission and the vaccination strategy prioritizing individuals aged 20–29, 30–39, and 40–49, sequentially (Row 2, Column 4). Four coverage rates, including 5%, 10%, 20%, and 30%, were assessed, with outcomes displayed as scattered dots across four columns within one subfigure. The rows of subfigures represent the two selected sequential vaccination strategies, while the columns correspond to the four hypothetical scenarios with varying levels of sexual transmission.

**Sensitivity analysis: alternative maximum uptake threshold of 70%**

We performed a sensitivity analysis to assess the impact of higher-than-expected vaccine hesitancy on mpox vaccine demand, assuming that the vaccine uptake in any age group was capped at 70% [20]. This led to a greater number of vaccine series required to achieve outbreak control and reduced effectiveness of targeted vaccination strategies with limited supply (Fig S30–S32).

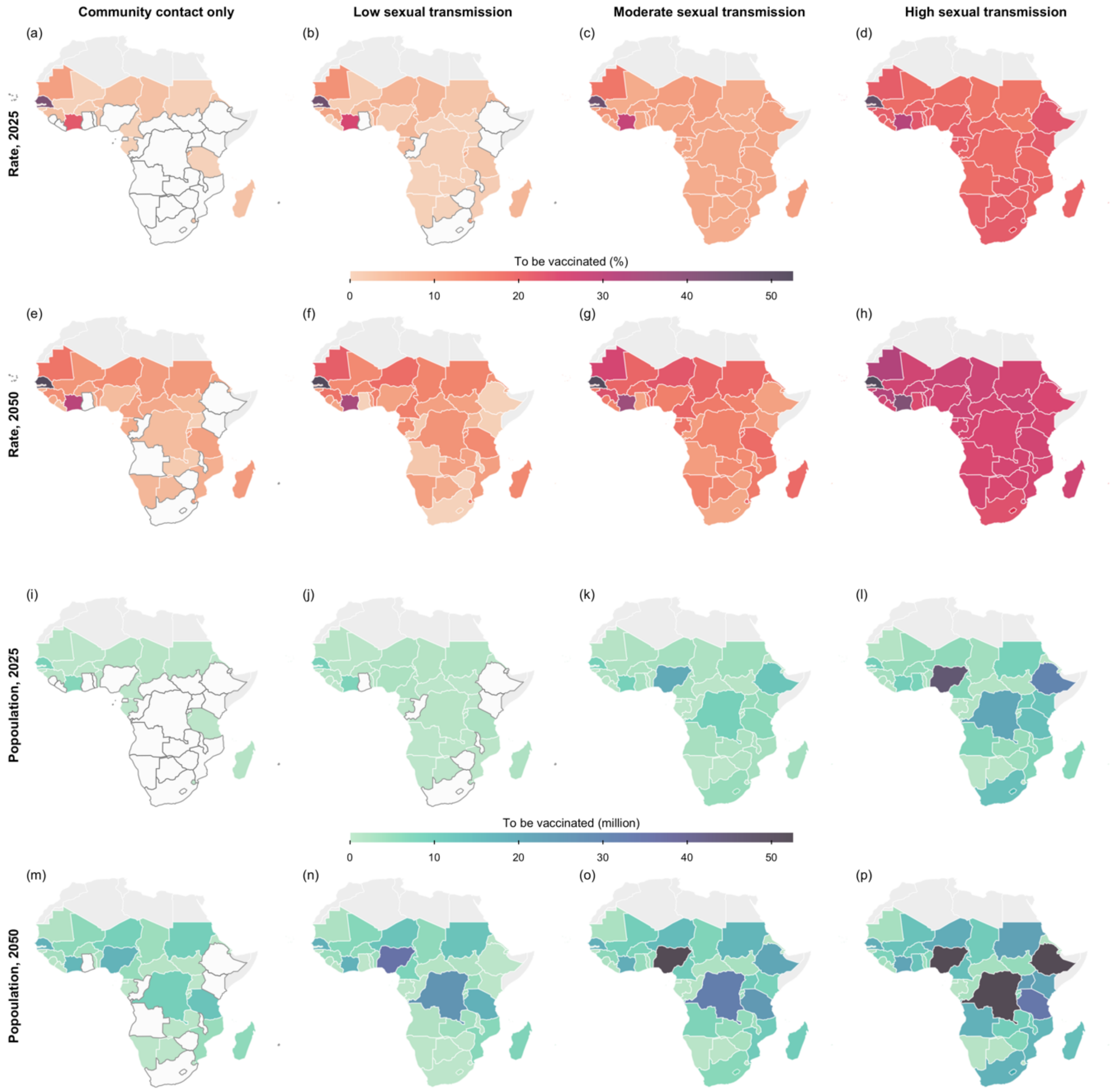

**Fig S30.** Projected minimal vaccine demand when the maximum coverage in any group was 70% [20]. Shown are the vaccine coverage rates (Row 1 and 2) and the number of individuals requiring vaccination (Row 3 and 4) to prevent secondary infections in each sub-Saharan African country modelled for 2025 (Row 1 and 3) and 2050 (Row 2 and 4), assuming mass vaccination. The four columns correspond to the four hypothetical scenarios with varying levels of sexual transmission. Countries which do not require vaccination are coloured in white, with borders outlined in dark grey. The base map layer (boundaries of African countries) is sourced from Natural Earth (https://www.naturalearthdata.com), available under the Public Domain license (https://www.naturalearthdata.com/about/terms-of-use/).

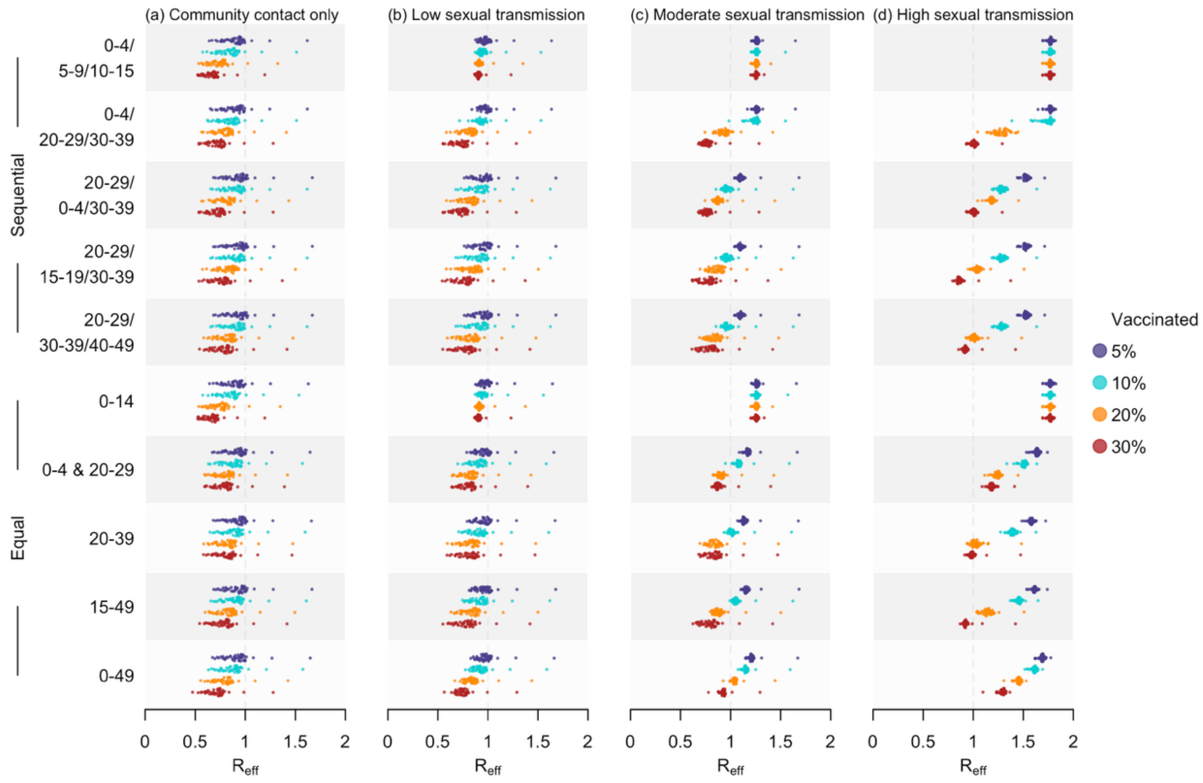

**Fig S31**. Estimated $R_{eff}$ in 2025 under diverse vaccination strategies. Doses were allocated using two allocation methods, including sequential (first five rows) and equal (last five rows), when the maximum coverage in any group was 70% [20]. Four coverage rates, including 5%, 10%, 20%, and 30%, were assessed, with outcomes summarised as shaded distributions in purple, blue, orange, and red, respectively. The four columns correspond to the four hypothetical scenarios with varying levels of sexual transmission.

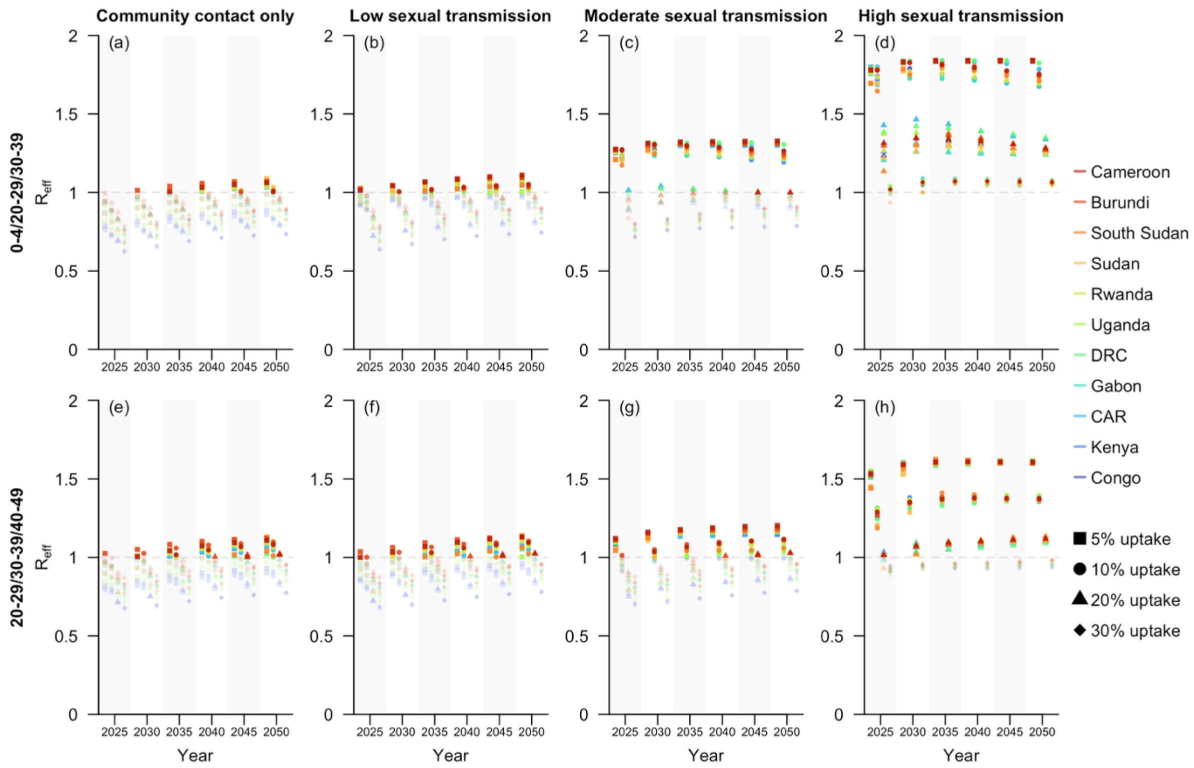

**Fig S32**. Projected $R_{eff}$ at five-year intervals from 2025 to 2050 under selected vaccination strategies for sub-Saharan African countries with documented local transmission of Clade I MPXV as of December 2024 [18], when the maximum coverage in any group was 70% [20]. These countries were ordered in descending estimated $R_{eff}$ for 2050, based on the scenario with high sexual transmission and the vaccination strategy prioritizing individuals aged 20–29, 30–39, and 40–49, sequentially (Row 2, Column 4). Four coverage rates, including 5%, 10%, 20%, and 30%, were assessed, with outcomes displayed as scattered dots across four columns within one subfigure. The rows of subfigures represent the two selected sequential vaccination strategies, while the columns correspond to the four hypothetical scenarios with varying levels of sexual transmission.

**Sensitivity analysis: alternative vaccine effectiveness of 75% or 85%**

In this sensitivity analysis, we assumed alternative mpox vaccine effectiveness of 75% or 85%. The projection results indicate that higher effectiveness is associated with lower demands for mpox vaccines, but consistent with the main analysis, both scenarios suggest to prioritise children under five years or young adults aged 20–29 for vaccination (Fig S33–S38).

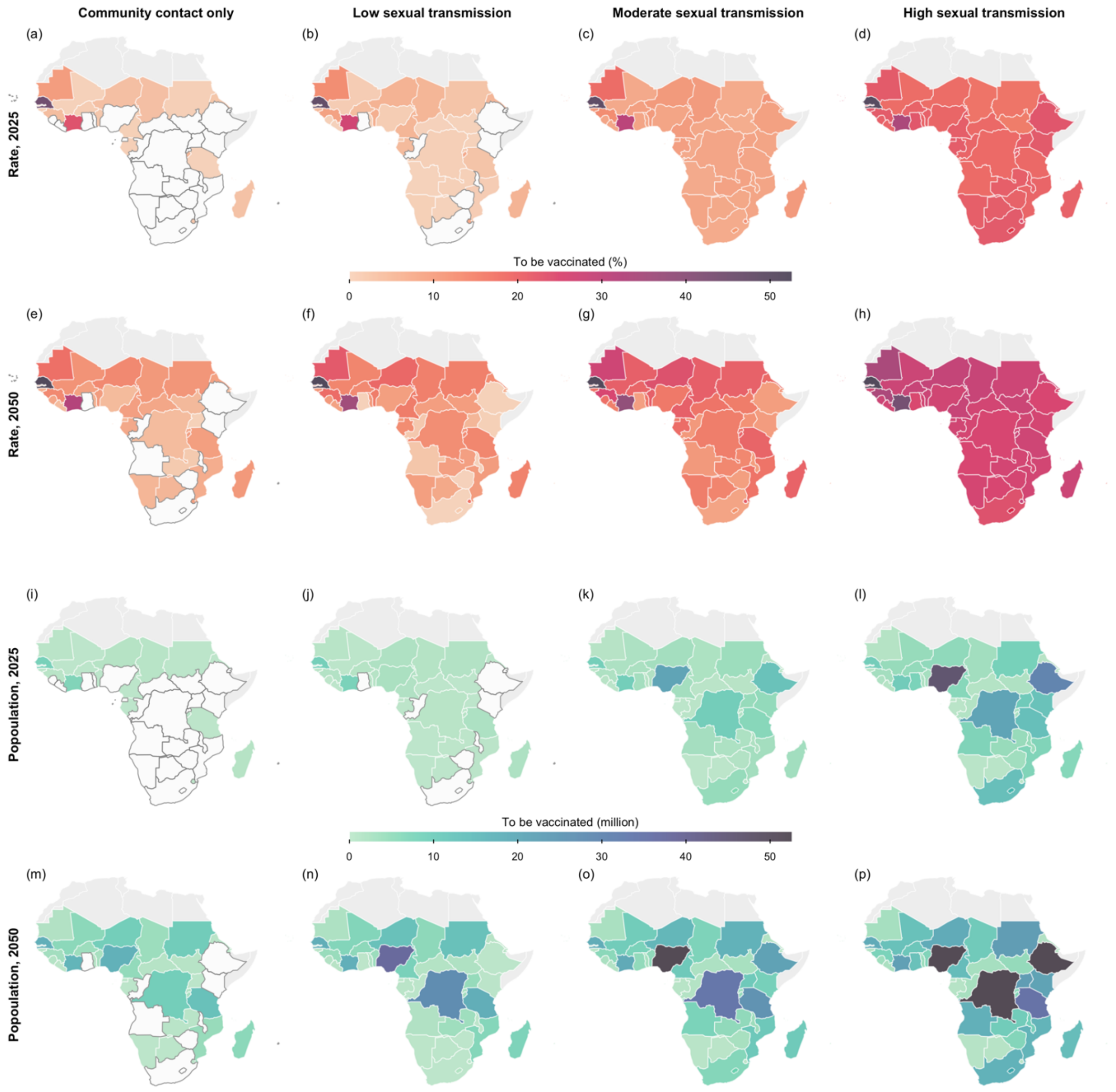

**Fig S33.** Projected minimal vaccine demand, assuming a vaccine effectiveness of 75%. Shown are the vaccine coverage rates (Row 1 and 2) and the number of individuals requiring vaccination (Row 3 and 4) to prevent secondary infections in each sub-Saharan African country modelled for 2025 (Row 1 and 3) and 2050 (Row 2 and 4), assuming mass vaccination. The four columns correspond to the four hypothetical scenarios with varying levels of sexual transmission. Countries which do not require vaccination are coloured in white, with borders outlined in dark grey. The base map layer (boundaries of African countries) is sourced from Natural Earth (https://www.naturalearthdata.com), available under the Public Domain license (https://www.naturalearthdata.com/about/terms-of-use/).

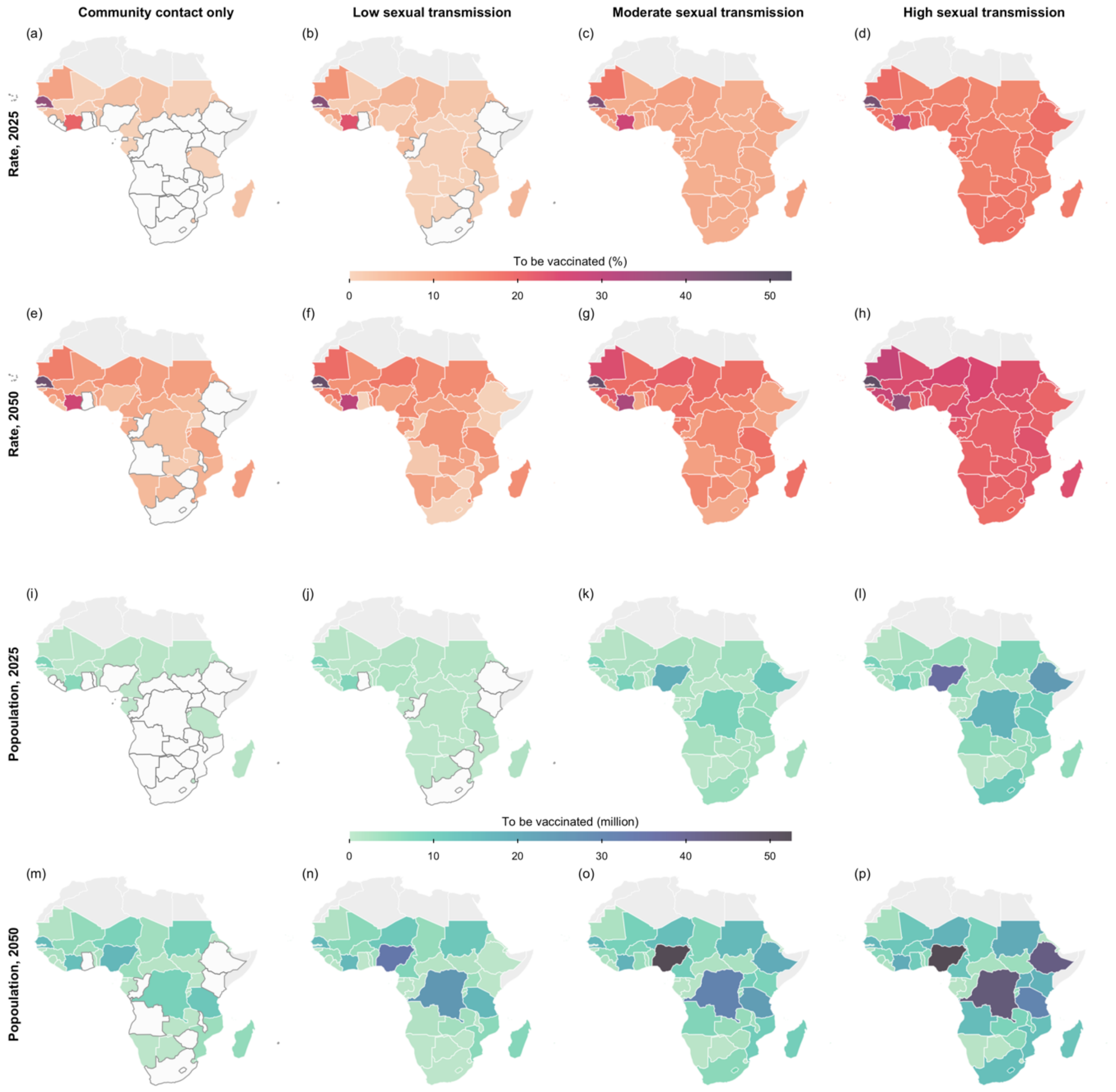

**Fig S34.** Projected minimal vaccine demand, assuming a vaccine effectiveness of 85%. Shown are the vaccine coverage rates (Row 1 and 2) and the number of individuals requiring vaccination (Row 3 and 4) to prevent secondary infections in each sub-Saharan African country modelled for 2025 (Row 1 and 3) and 2050 (Row 2 and 4), assuming mass vaccination. The four columns correspond to the four hypothetical scenarios with varying levels of sexual transmission. Countries which do not require vaccination are coloured in white, with borders outlined in dark grey. The base map layer (boundaries of African countries) is sourced from Natural Earth (https://www.naturalearthdata.com), available under the Public Domain license (https://www.naturalearthdata.com/about/terms-of-use/).

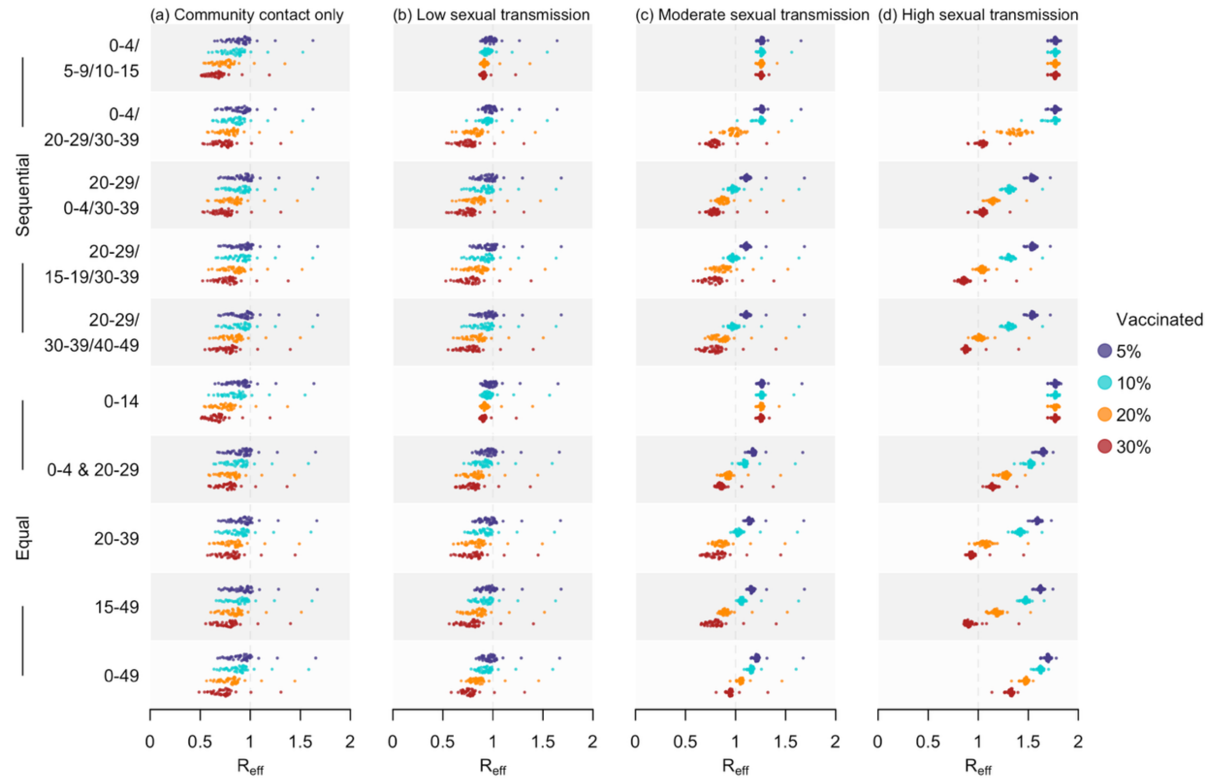

**Fig S35**. Estimated $R_{eff}$ in 2025 under diverse vaccination strategies and a vaccine effectiveness of 75% for all age groups. Doses were allocated using two allocation methods, including sequential (first five rows) and equal (last five rows). Four coverage rates, including 5%, 10%, 20%, and 30%, were assessed, with outcomes summarised as shaded distributions in purple, blue, orange, and red, respectively. The four columns correspond to the four hypothetical scenarios with varying levels of sexual transmission.

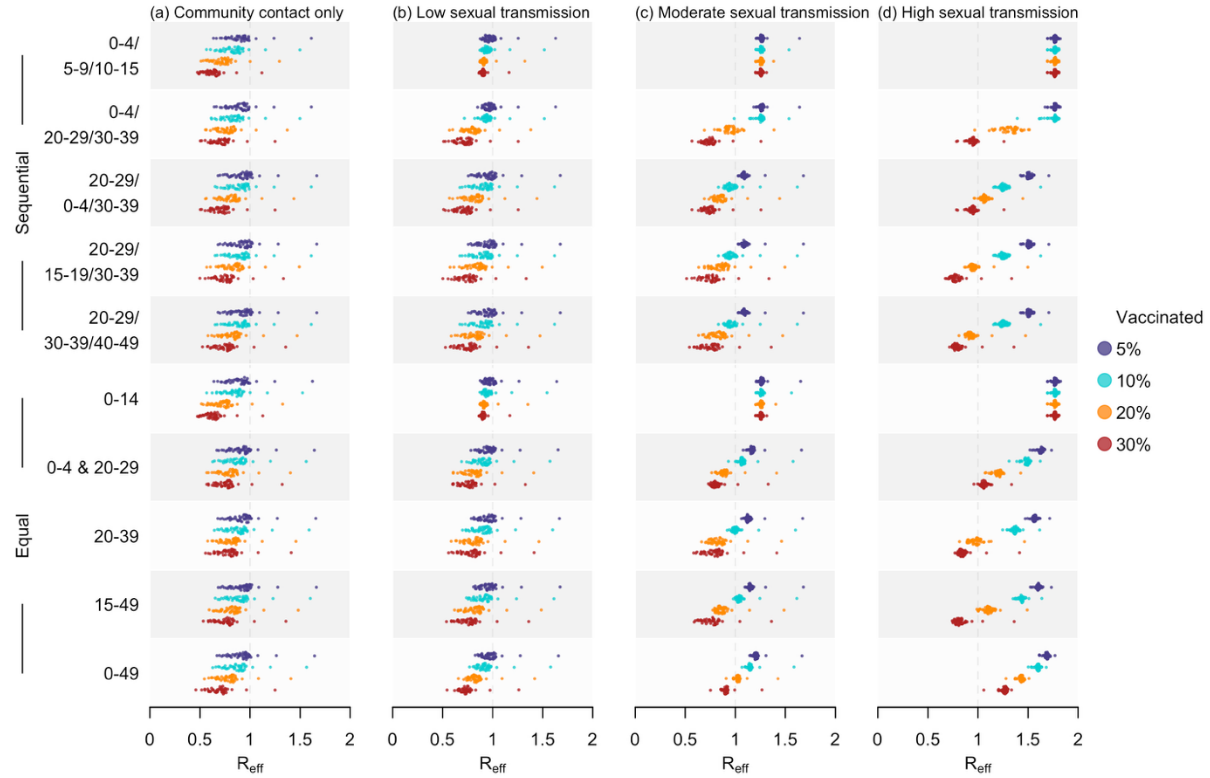

**Fig S36**. Estimated $R_{eff}$ in 2025 under diverse vaccination strategies and a vaccine effectiveness of 85% for all age groups. Doses were allocated using two allocation methods, including sequential (first five rows) and equal (last five rows). Four coverage rates—5%, 10%, 20%, and 30%—were assessed, with outcomes summarised as shaded distributions in purple, blue, orange, and red, respectively. The four columns correspond to the four hypothetical scenarios with varying levels of sexual transmission.

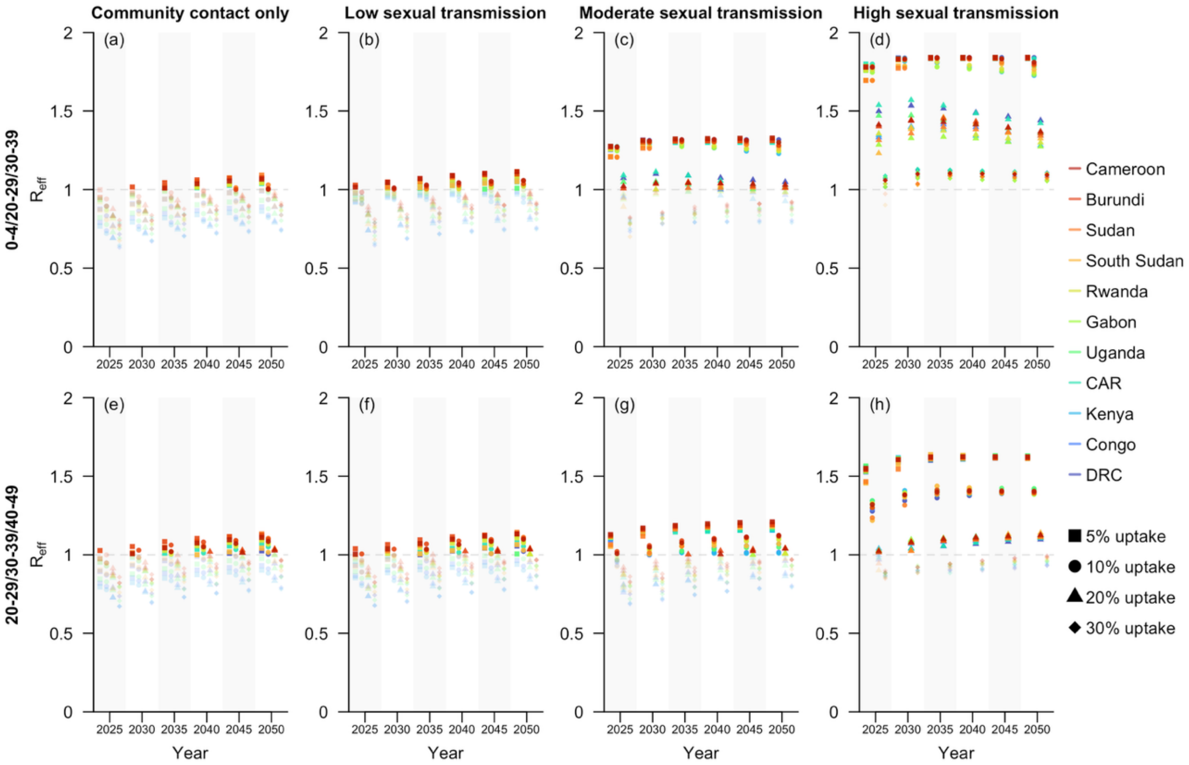

**Fig S37**. Projected $R_{eff}$ at five-year intervals from 2025 to 2050 under selected vaccination strategies for sub-Saharan African countries with documented local transmission of Clade I MPXV as of December 2024 [18], assuming a vaccine effectiveness of 75%. These countries were ordered in descending estimated $R_{eff}$ for 2050, based on the scenario with high sexual transmission and the vaccination strategy prioritizing individuals aged 20–29, 30–39, and 0–4, sequentially (Row 2, Column 4). Four coverage rates, including 5%, 10%, 20%, and 30%, were assessed, with outcomes displayed as scattered dots across four columns within one subfigure. The rows of subfigures represent the two selected vaccination strategies, while the columns correspond to the four hypothetical scenarios with varying levels of sexual transmission.

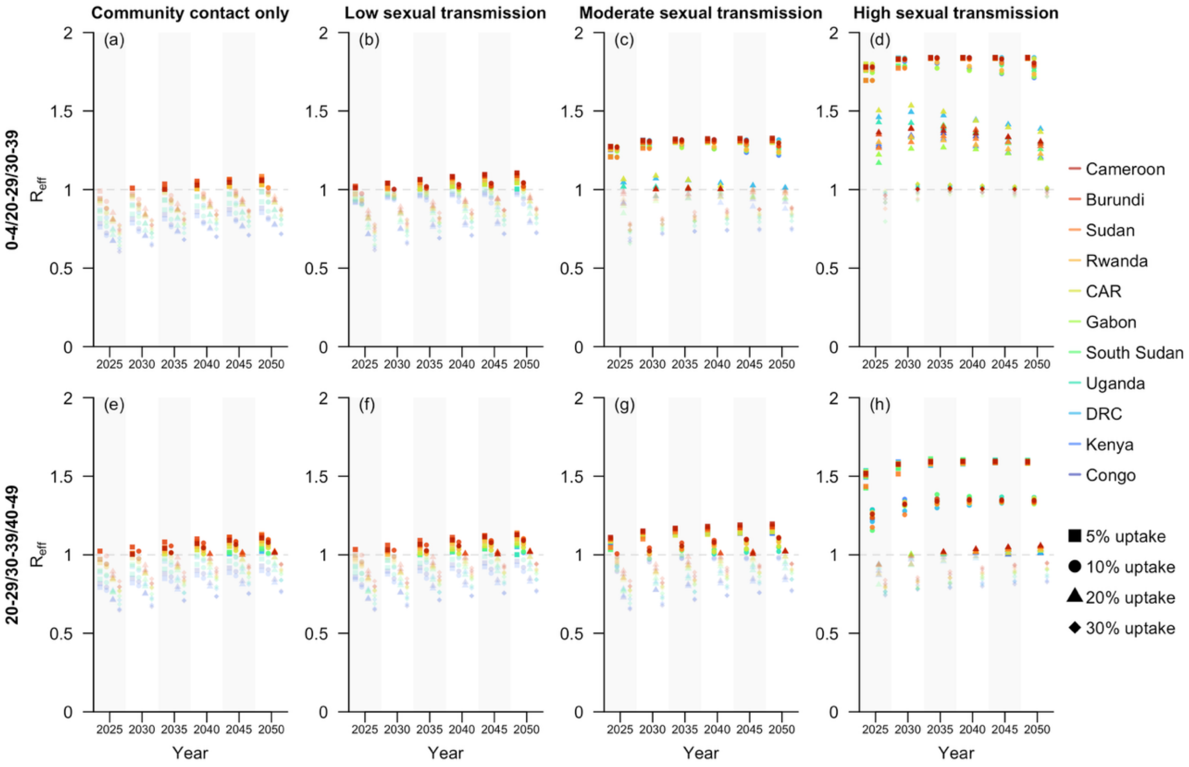

**Fig S38**. Projected $R_{eff}$ at five-year intervals from 2025 to 2050 under selected vaccination strategies for sub-Saharan African countries with documented local transmission of Clade I MPXV as of December 2024 [18], assuming a vaccine effectiveness of 85%. These countries were ordered in descending estimated $R_{eff}$ for 2050, based on the scenario with high sexual transmission and the vaccination strategy prioritizing individuals aged 20–29, 30–39, and 0–4, sequentially (Row 2, Column 4). Four coverage rates, including 5%, 10%, 20%, and 30%, were assessed, with outcomes displayed as scattered dots across four columns within one subfigure. The rows of subfigures represent the two selected vaccination strategies, while the columns correspond to the four hypothetical scenarios with varying levels of sexual transmission.
